# Quantifying the impact of delaying the second COVID-19 vaccine dose in England: a mathematical modelling study

**DOI:** 10.1101/2022.08.08.22278528

**Authors:** Natsuko Imai, Thomas Rawson, Edward S Knock, Raphael Sonabend, Yasin Elmaci, Pablo N Perez-Guzman, Lilith K Whittles, Divya Thekke Kanapram, Katy AM Gaythorpe, Wes Hinsley, Bimandra A Djaafara, Haowei Wang, Keith Fraser, Richard G FitzJohn, Alexandra B Hogan, Patrick Doohan, Azra C Ghani, Neil M Ferguson, Marc Baguelin, Anne Cori

**Author notes:** contributed equally (Natsuko Imai, Thomas Rawson).

## Abstract

**Background:** The UK was the first country to start national COVID-19 vaccination programmes, initially administering doses 3-weeks apart. However, early evidence of high vaccine effectiveness after the first dose and the emergence of the Alpha variant prompted the UK to extend the interval between doses to 12-weeks. In this study, we quantify the impact of delaying the second vaccine dose on the epidemic in England.

**Methods:** We used a previously described model of SARS-CoV-2 transmission and calibrated the model to English surveillance data including hospital admissions, hospital occupancy, seroprevalence data, and population-level PCR testing data using a Bayesian evidence synthesis framework. We modelled and compared the epidemic trajectory assuming that vaccine doses were administered 3-weeks apart against the real vaccine roll-out schedule. We estimated and compared the resulting number of daily infections, hospital admissions, and deaths. A range of scenarios spanning a range of vaccine effectiveness and waning assumptions were investigated.

**Findings:** We estimate that delaying the interval between the first and second COVID-19 vaccine doses from 3- to 12-weeks prevented an average 64,000 COVID-19 hospital admissions and 9,400 deaths between 8^th^ December 2020 and 13th September 2021. Similarly, we estimate that the 3-week strategy would have resulted in more infections and deaths compared to the 12-week strategy. Across all sensitivity analyses the 3-week strategy resulted in a greater number of hospital admissions.

**Interpretation:** England’s delayed second dose vaccination strategy was informed by early real-world vaccine effectiveness data and a careful assessment of the trade-offs in the context of limited vaccine supplies in a growing epidemic. Our study shows that rapidly providing partial vaccine-induced protection to a larger proportion of the population was successful in reducing the burden of COVID-19 hospitalisations and deaths. There is benefit in carefully considering and adapting guidelines in light of new emerging evidence and the population in question.

**Funding:** National Institute for Health Research, UK Medical Research Council, Jameel Institute, Wellcome Trust, and UK Foreign, Commonwealth and Development Office, National Health and Medical Research Council.

**Research in Context:** *Evidence before this study:* We searched PubMed up to 10^th^ June 2022, with no language restrictions using the following search terms: (COVID-19) AND (vaccin*) AND (dose OR dosing) AND (delay OR interval) AND (quant* OR assess* OR impact). We found 14 studies that explored the impact of different vaccine dosing intervals. However, the majority were prospective assessments of optimal vaccination strategies, exploring different trade-offs between vaccine mode of action, vaccine effectiveness, coverage, and availability. Only two studies retrospectively assessed the impact of different vaccination intervals. One assessed the optimal timing during the epidemic to switch to an extended dosing interval, and the other assessed the risk of all-cause mortality and hospitalisations between the two dosing groups.

*Added value of this study:* Our data synthesis approach combines real-world evidence from multiple data sources to retrospectively quantify the impact of extending the COVID-19 vaccine dosing interval from the manufacturer recommended 3-weeks to 12-weeks in England.

*Implications of all the available evidence:* Our study demonstrates that rapidly providing partial vaccine-induced protection to a larger proportion of the population was successful in reducing the COVID-19 hospitalisations and mortality. This was enabled by rapid and careful monitoring of vaccine effectiveness as nationwide vaccine programmes were initiated, and adaptation of guidelines in light of emerging evidence.

## Introduction

The Pfizer-BioNTech and Oxford–AstraZeneca COVID-19 vaccines were given regulatory approval on 2^nd^ and 30^th^ December 2020 respectively in the UK, making it the first country to start a nationwide COVID-19 vaccination campaign ^1,2^. Initially, the two scheduled doses of each vaccine product were administered similarly to the clinical trials at 3 to 4 weeks apart. However, the Joint Committee on Vaccination and Immunisation (JCVI) recommended delaying the second dose from 3- to 12-weeks after the first dose ^3^. This was based on evidence that one vaccine dose provided 70-90% protective effectiveness against symptomatic disease ^4^ and the higher peak antibody levels observed amongst individuals receiving a delayed second dose ^5^. This decision was also made in response to the emergence of the Alpha variant of concern (VOC).

The delayed second dose strategy prioritised delivery of first vaccine doses and thus high partial short-term protection to as many people as possible. This strategy received some criticism, citing the lower level of protection offered to the most vulnerable by delaying the second dose, the limited number of studies available to support this change from the trial protocols, and concerns of partial vaccination accelerating the emergence of new vaccine-evading VOCs ^6,7^. However, prompted by vaccine shortages and the emergence of Alpha, several countries including Canada ^8^, Denmark ^9^, Norway ^10^, India ^11^, and South Africa ^12^ also extended the average time between first and second doses. Previous studies have explored the potential impact and optimisation of mass vaccination schedules under different assumptions of vaccine effectiveness (VE), vaccine mode of action, waning immunity, and the degree of non-pharmaceutical interventions (NPIs) in place ^13,14^. They highlighted that more hospitalisations and deaths could be prevented by adopting a delayed second dose strategy, but that results were particularly sensitive to VE assumptions and vaccine mechanisms, underscoring the importance of continued NPIs ^15–17^.

In early 2021, informed by mathematical modelling, the UK Government published a “roadmap out of lockdown” for England, a policy setting out the conditions for and expected timeline of a stepwise lifting of NPIs. Between 8^th^ March and 19^th^ July, 2021, NPIs were incrementally lifted as vaccination coverage increased ^18^. During this time the recommended dose interval was initially shortened from 12 to 8 weeks only for priority groups 1-9 ^19^, then for all over 40-year-olds, and finally for all cohorts, in response to the emergence of the Delta variant and increasing vaccine supply ^20^ (supplementary material figure S9). Here we retrospectively assess the impact of delaying the second vaccine dose in England and examine what the epidemic trajectory in terms of SARS-CoV-2 transmission, infections, and COVID-19 hospital admissions and deaths would have looked like if a three-week dose interval had been maintained.

## Methods

### Study design

Ethics permission was sought for the study via Imperial College London’s (London, UK) standard ethical review processes and was approved by the College’s Research Governance and Integrity Team (ICREC reference 21IC6945).

### Epidemiological model and fitting

We used a previously described stochastic transmission model ^21^ which reliably captures the age-specific scale and timing of the SARS-CoV-2 pandemic in England in 2021 ^22^ to examine the impact of vaccination in the first 9 months of its roll-out. This was a period marked by the transition – in May and June 2021 – from the Alpha (B.1.1.7) variant to the Delta variant (B.1.617.2). Using a Bayesian inference framework, we fit a two-variant model to multiple data sources including daily hospital admissions and bed occupancy, deaths, population-level PCR prevalence, and serological data, for each English National Health Service (NHS) region (see supplementary materials for detail). To explicitly capture the emergence of the Delta variant, we additionally fitted the model to the UK Health Security Agency (UKHSA) variant and mutation dataset (VAM). Delta is seeded at a region-specific date determined by the model fit, no earlier than 8^th^ March 2021.

Our study period ranges from 8^th^ December 2020, when vaccination started, to 13^th^ September 2021, just before the introduction of booster doses in England which are not considered here. We modelled vaccine roll-out as reported in the NHS vaccine administration data including the distribution of doses by age and vaccine type. The dosing interval is informed by the real vaccination data; thus, the actual average dosing interval may differ slightly from reported guidelines. We refer to this as the “12-week strategy”.

VE against infection, symptomatic disease, hospitalisation, death, and onward transmission for each variant were informed by effectiveness studies in the UK or England ^4,23,32–36,24–31^. We assumed full effectiveness of the first and second dose is reached after three- and one-week post dose one and two respectively. For our baseline analysis, we assume the same VE under the 12- and 3-week strategies, and no waning post-dose one regardless of the dosing interval.

We allowed for waning infection- and vaccine-induced protection, imperfect cross-protection between Alpha and Delta, and a fitted increased severity of Delta infection relative to Alpha ^32^. We assumed that infection-induced immunity against the same SARS-CoV-2 variant wanes following an exponentially distributed duration with a mean of 6 years ^37^. To model the waning of vaccine-induced protection, we assumed individuals who had received a second dose moved to a waned compartment of reduced protection after, on average, 24-weeks. To ensure the population average VE in our compartmental model was consistent with the observations, we fitted VE in the second dose and in the reduced protection (i.e. waned) compartments, for all vaccines and disease outcomes, to the VE estimated by Andrews *et al*. ^26,38^ (figure 1) (see supplementary material “*Waning of vaccine-induced immunity*” and figure S4 for Alpha values).

**Figure 1:**
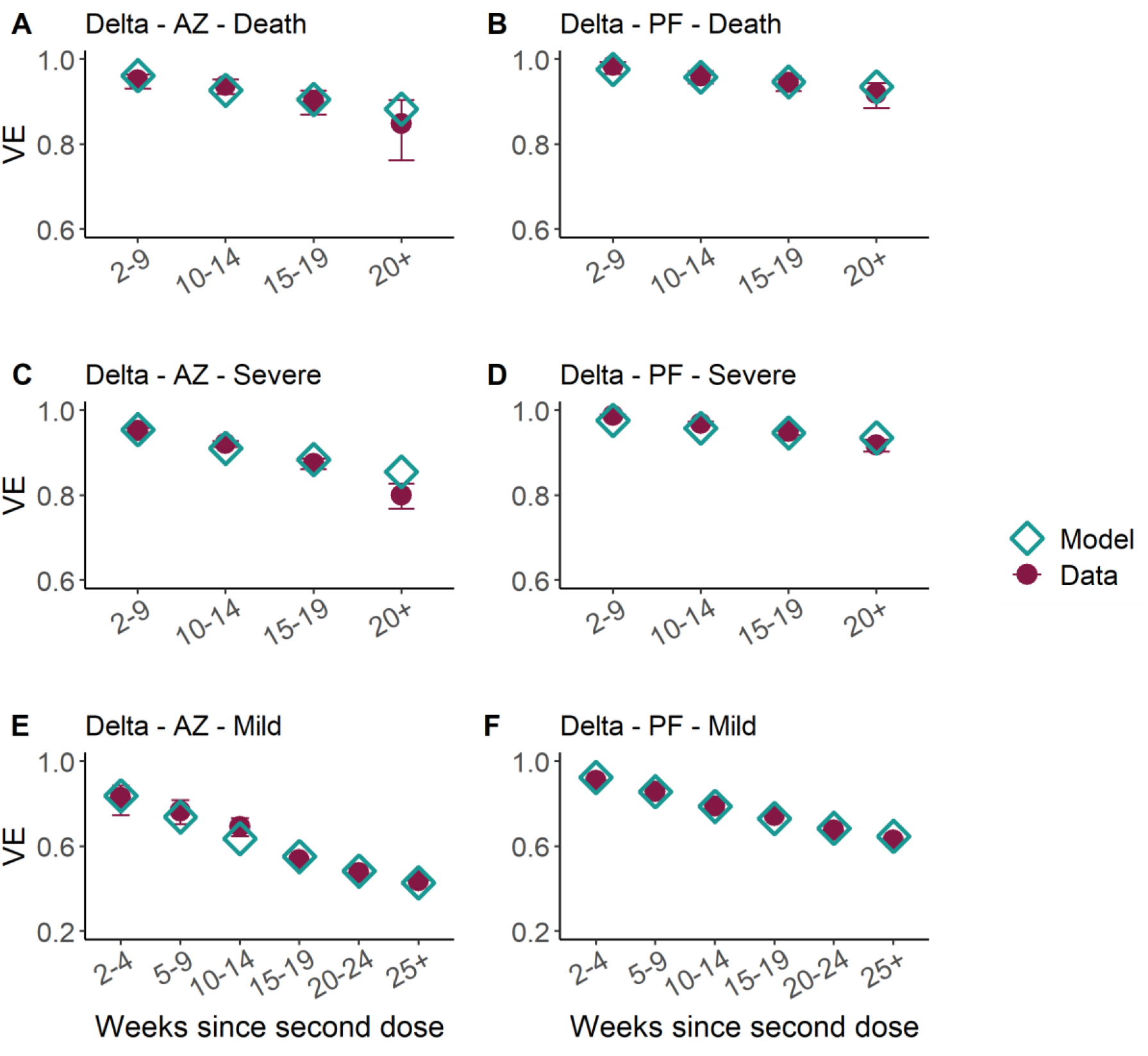
Vaccine effectiveness in weeks since second dose of AstraZeneca (AZ, left column) and Pfizer (PF, right column) vaccines against Delta (see supplementary materials for values against Alpha) for death (top), severe disease, (middle) and mild disease/infection (bottom). We assume the same protection against infection and mild disease. Purple points and uncertainty ranges show the VE estimates from Andrews et al, and the turquoise points our model assumptions. We assumed that the Moderna vaccine has the same VE as PF. Note the x-axis differs in the bottom row. See supplementary material for Alpha values.

### Assessing the potential impact of alternative vaccination schedules

We explored a counterfactual scenario with a 3-week interval (“3-week strategy”) between vaccine doses to assess the impact of delaying the second vaccine dose interval to 12-weeks (“12-week strategy”) on the epidemic trajectory. We assumed that the total daily number of vaccines administered matched that reported by the NHS. We compared the reported numbers and age distribution of hospital admissions between the fitted 12-week delay scenario and the counterfactual 3-week delay scenario. For our baseline VE assumption, we assumed the same VE for both 3-week and 12-week dosing intervals (supplementary material, table S3).

We used the transmission rate at each roadmap step as estimated by fitting the model to the full epidemic trajectory up to 13 September 2021 to capture the impact of changing restrictions and behaviours on transmission rates and use these estimated beta values to simulate the 3-week counterfactual strategy.

### Sensitivity Analyses

We ran a number of sensitivity analyses to assess the impact of assumptions on VE and waning of vaccine-induced protection on our results (see supplementary material *“Sensitivity Analyses”* and Table S9).

To understand the impact of the different VE assumptions by dosing interval on the epidemic trajectory, we explored two scenarios where we assumed a 10% absolute reduction in post dose 2 VE for the 3-week dosing interval: i) against infection or mild disease only, and ii) across all vaccine end points compared with the 12-week interval VE for both Alpha and Delta variants ^39^. We further explored uncertainty in all VE estimates by examining the impact of a 15% relative increase and decrease in VE for all end points and doses against Alpha and Delta for both the 12-week and 3-week strategies. We also explored the impact of slower or faster waning of VE over time and the impact of immediate vaccine-induced protection upon first dose vaccination (see supplementary material). Finally, to allow for a small level of waning of protection after the first dose but before the second, we allowed for an absolute 10% increase in the first dose VE across all end points for the 3-week strategy (where there would be less time for such waning).

### Role of the funding source

The funders of this study had no role in the study design, data collection, data analysis, data interpretation, or writing of the report.

## Results

Under the “12-week interval” between first and second doses adopted in the UK, first doses were distributed to a greater number of individuals. In comparison, with a shorter 3-week interval between first and second vaccine doses, second doses start being rolled out sooner as the most vulnerable individuals are prioritised for full vaccination (figure 2A). As previously shown ^21^, our model effectively captures national SARS-CoV-2 trends in daily hospital admissions by age between 1^st^ January and 13^th^ September 2021 (figure 2B, supplementary material figure S10). A 3-week interval between first and second vaccine doses led to higher peak daily hospital admissions during the summer Delta wave with 2,080 (95% credible interval (CrI): 1,760 – 2,440) hospitalisations on 22^nd^ July 2021 in our baseline scenario (figure 2B). Overall, we estimated 297,000 (95% CrI: 279,000-327,000) cumulative hospitalisations between 8^th^ December 2020 and 13^th^ September 2021 with the 3-week delay compared to the estimated 233,000 (95% CrI: 226,000-237,000) hospitalisations with the delayed second dose over the same time period (figure 2C). Similarly, the total number of infections and deaths were also higher in the 3-week, compared to the 12-week strategy (supplementary table 1).

**Figure 2:**
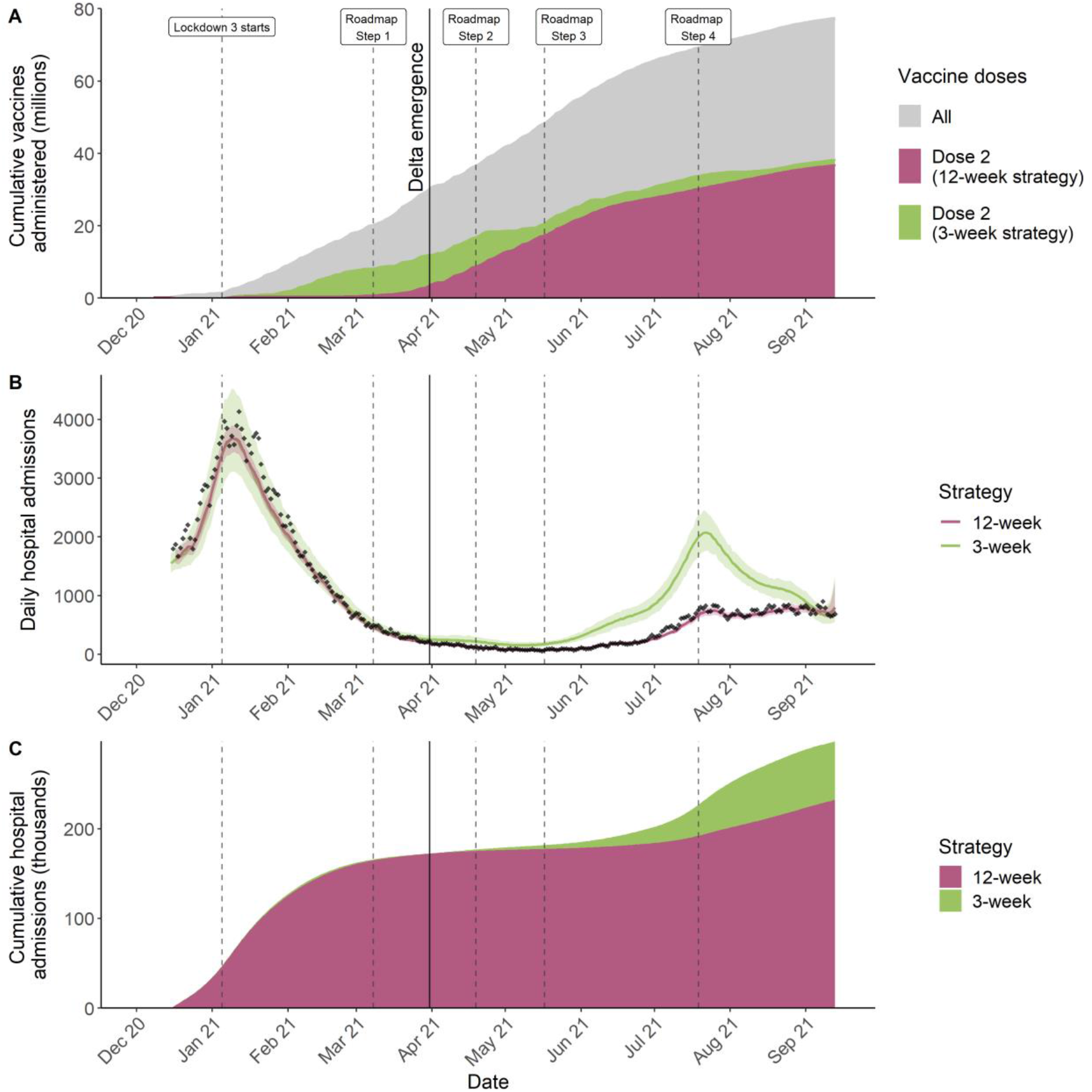
COVID-19 vaccines administered and epidemic trajectory in England. Results are shown for central vaccine effectiveness and waning assumptions (see supplementary material for sensitivity analyses results). A) Cumulative vaccine doses administered between 8th December 2020 and 13th September 2021. Grey ribbon shows all (first and second doses), purple and green show the second doses administered under the 12-week and 3-week strategy respectively over time. B) Daily hospital admissions. The black points show the data, the purple line and shaded area show the median model fit and 95% CrI. The green line and shaded area show the median and 95% CrI of the simulated daily hospital admissions under the counterfactual assuming a 3-week delay between vaccine doses. C) Cumulative hospital admissions over time under the 12-week (purple) and 3-week (green) strategy. The vertical dashed lines show the roadmap out of lockdown steps and the vertical solid line the time when the Delta variant emerged. In panel B, uncertainty is greater for the 3-week scenario, which was explored through unconstrained simulations, than in the 12-week fit, which was obtained by particle filtering and is thus more constrained by the data (see supplementary material for methods).

Cumulative COVID-19 hospitalisations were greater in the 3-week delay strategy, compared to the 12-week delay strategy (figure 2C). Whilst this difference was observed throughout, it became more apparent from late May 2021 onwards following the emergence of the Delta variant. This difference stabilised in late September, as under the 3-week strategy, the epidemic peaked and then declined at a much faster rate than the observed 12-week strategy due to the rapid depletion of susceptible individuals.

The higher hospitalisations estimated under the counterfactual 3-week strategy was driven by, overall, fewer individuals benefiting from full or partial vaccine-induced protection. For example, by the beginning of April 2021, 7.7 million fewer individuals had any level (one or two doses) of vaccine-induced protection under the 3-week, compared to the 12-week strategy. This trend continued in April and May (figure 3A), with a large difference in the total number of individuals with any vaccine protection between the two strategies. This gap then narrowed from June 2021 onwards.

**Figure 3:**
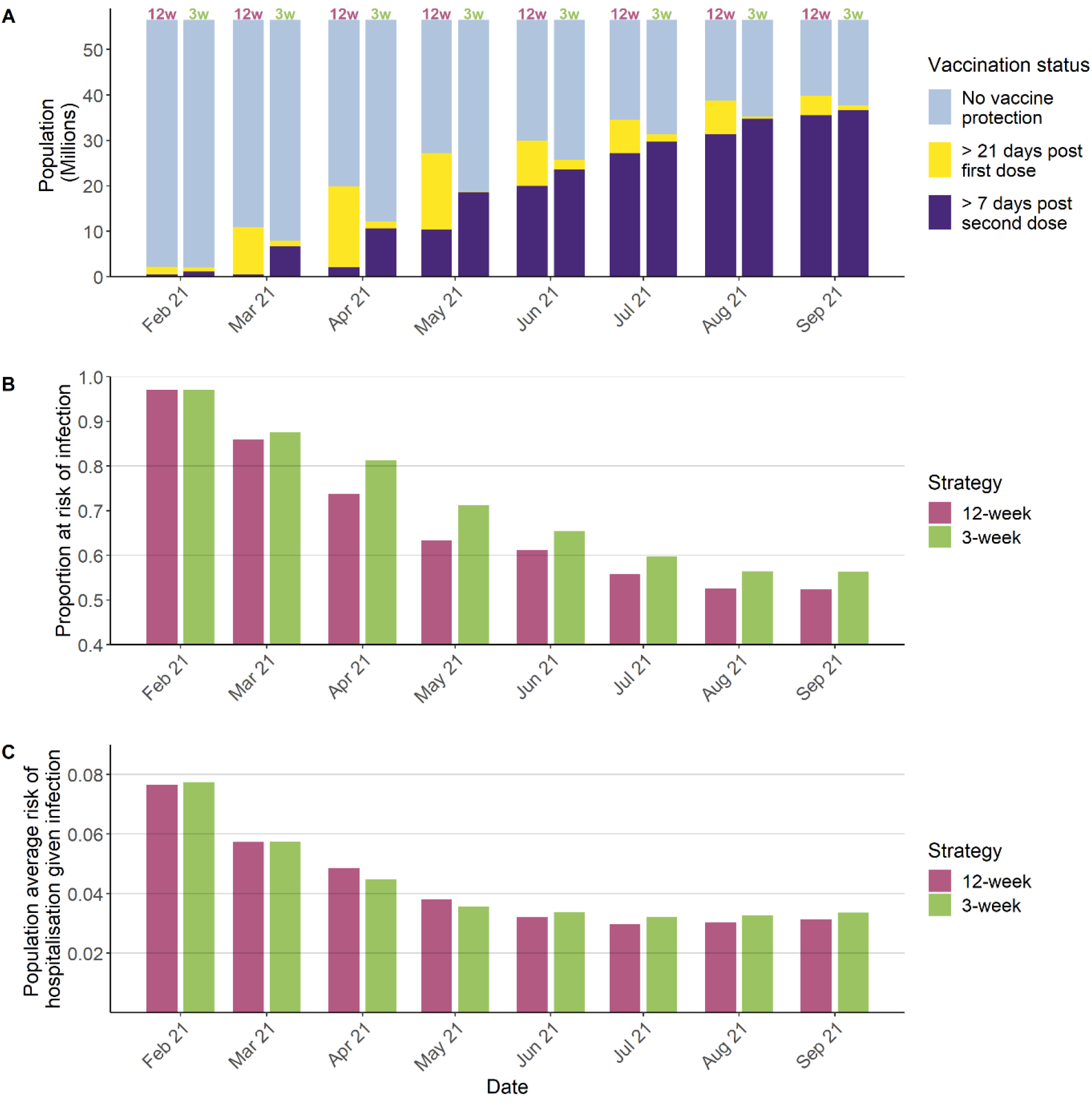
Vaccine-induced protection against COVID-19. A) Vaccination status within the population (millions of people) for each month for the 12-week strategy (model fit, “12w”) and 3-week strategy (counterfactual, “3w”). Colours show how the distribution of >21 days post first- and >7 days post second-dose and two-dose vaccine-induced protection, and population with no vaccine-induced protection changes over time. B) Average proportion of the population without vaccine protection against infection with Alpha or Delta. C) Average population risk of hospitalisation given infection, for first infections. This accounts for vaccine-induced protection, but not naturally acquired protection from prior infections. In B) and C), the 12-week strategy (model fit) is shown in purple and the 3-week strategy (counterfactual) in green.

The number of individuals with reduced protection after dose 2 was higher throughout in the 3-week strategy: vaccine protection under the 3-week interval had already started to wane in some individuals by March 2021 (supplementary figure S16). This increased over time with 10 million people 24-weeks or more post dose two, compared to 7 million people under the 3-week and 12-week strategies respectively, by the beginning of July 2021 when hospitalisations peaked (supplementary figure S16). This was due to prioritising fully vaccinating, with second doses, each eligible age group before the next group became eligible for vaccination. Thus, the number of individuals with reduced protection was always much greater in the 3-week than the 12-week strategy. Throughout the epidemic, we find a lower average risk of infection (figure 3B) and a lower risk of hospitalisation if infected through vaccination in the 12-week compared to the 3-week vaccination strategy, and a lower risk of hospitalisation if infected (figure 3C). In the 3-week strategy, although the most vulnerable oldest age groups received greater protection earlier, the population level severity was still higher than in the 12-week strategy (figure 3C).

We estimate the distribution of hospitalisations by age remains broadly similar under both vaccination strategies (supplementary material, figure S17), with most hospitalisations occurring in the older age groups (figure 4).

**Figure 4:**
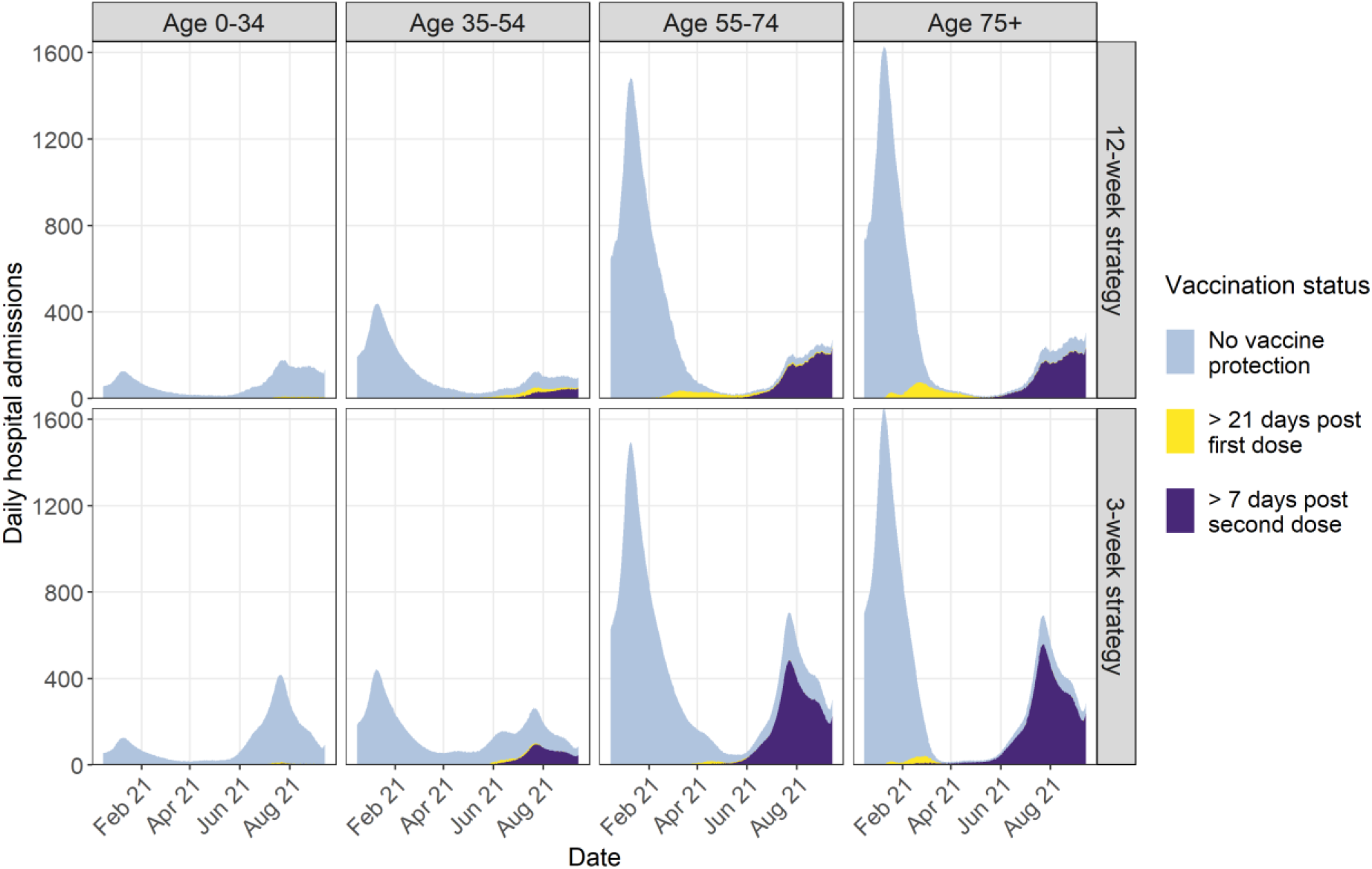
Modelled daily COVID-19 hospital admissions over time by age group (columns) for the 12-week (top row) and 3-week (bottom row) strategy. Colours show the status of vaccine-induced protection of hospitalised individuals.

The 12-week strategy meant a greater number of individuals remained partially protected for longer before their second dose was effective. This is reflected in a small proportion of older individuals in this category (yellow in top row, figure 4) being hospitalised during the study period. Conversely, under the 3-week strategy, there were almost no hospitalisations amongst individuals with partial protection from a single vaccine dose compared to the 12-week strategy. However, this short-term benefit of the 3-week strategy was rapidly lost, with many more hospitalisations occurring in the summer of 2021 compared to the 12-week strategy (figure 4).

These trends were observed regardless of age group. Across both strategies, the most vulnerable individuals (80+ years) were vaccinated first. The 3-week strategy, however, gave higher protection quickly for the most vulnerable individuals. Thus, by the peak of the Delta wave, vaccine-induced protection in these individuals who received their last vaccine dose at least 6 months ago would have decreased, making them vulnerable once more to severe outcomes (supplementary figure S17). For example, by 13^th^ September, we estimated 2.8 million and 3.3 million individuals aged 75+ years had their vaccine 24 or more weeks ago under the 12-week and 3-week strategies respectively. Prioritising fully vaccinating the most vulnerable population as quickly as possible under the 3-week strategy would have reduced the short-term morbidity and mortality in the older age groups, but increased the total long-term burden due to waning vaccine-induced protection and higher levels of virus circulating in younger age groups who drive transmission.

Across all sensitivity analyses, we found that the 3-week strategy led to a greater number of hospitalisations and deaths compared to the 12-week strategy (supplementary material, figures S18-S25).

## Discussion

Our study explored the impact of the COVID-19 vaccination strategy of delaying the interval between vaccine doses to 12 weeks (rather than the 3–4 weeks assessed in clinical trials) in England, comparing the observed number of hospitalisations and deaths to a counterfactual scenario where the vaccine dose interval remained at the recommended 3 weeks. We estimate in our baseline scenario, delaying the interval between the first and second COVID-19 doses from 3 to 12 weeks averted on average 64,000 hospitalisations in England by 13^th^ September 2021 and averted between 39,000 and 211,000 total hospitalisations across all sensitivity analyses.

The 12-week strategy led to a larger number of partially protected individuals which temporarily led to more hospitalisations and deaths in older age groups in spring 2021 (figure 3, supplementary table). However, the strategy provided partial but high protection to more age groups, including younger groups that may sustain transmission ^40^, which eventually indirectly protected the vulnerable. Conversely, prioritising fully vaccinating the most vulnerable with two vaccine doses under the 3-week strategy led to a large proportion of the most at-risk population having a lower level of vaccine-induced immunity during the Delta wave in summer 2021 due to waning immunity which resulted in higher peak hospitalisations. The age-prioritisation of vaccine roll-out and the choice of dosing interval was effectively a trade-off between protecting those who were most at risk of severe disease and death and limiting infectivity in younger age groups which would also indirectly protect vulnerable groups. The magnitude of the impact of the 12-week strategy may also be sensitive to the timing of the Delta wave. Had Delta emerged 3 months later, the 12-week strategy would likely have still been beneficial than the 3-week strategy, but to a smaller relative extent.

Retrospective assessments of the impact of different vaccination strategies in other countries are not yet available. However, our findings are consistent with previous prospective simulation studies that explored optimal vaccination strategies under different assumptions of VE, stringency of NPIs, vaccine mode of action, and waning immunity.

These studies also found that prioritising partial protection of a larger proportion of the population by increasing the dosing interval could reduce the number of hospitalisations and deaths ^13–17^. Barmpounakis *et al*. found that the optimal strategy was to prioritise fully vaccinating the most elderly, before switching to a delayed second dose strategy for those under 75 years ^41^. This is effectively the strategy England pursued, with around 25% of 80+ years old vaccinated with a first dose before JCVI changed their guidance. This combined strategy essentially targets a combination of susceptibility in the highest risk groups and infectivity in the younger age groups. Mathematical models are valuable tools to quantitatively evaluate vaccination programmes, improve their design, and monitor new vaccine initiatives ^21,42^. By using a Bayesian evidence-synthesis approach, we were able to integrate multiple sources of data including cases, hospitalisations, deaths, infection prevalence, and sero-surveys to ensure we captured the epidemic in England accurately. Our model is also able to explicitly account for the introduction of the Delta variant, different vaccine end points, and the waning of infection- and vaccine-induced protection. Thus, counterfactual scenarios simulated using our model calibrated to these data allow us to robustly explore the impact of alternative vaccination strategies and a wide range of sensitivity analyses. Additionally, by using the exact reported number of vaccinations distributed each week in both model fitting and simulations, we can mimic the actual capacity of the vaccination program.

Our study has some limitations. First, there is substantial uncertainty in VE by dosing interval in the UK. Studies are limited due to the switch in strategy early in the vaccine roll-out ^39^. VE studies in countries where a 3-week interval between doses was maintained suggest that VE is slightly lower compared to a 12-week interval ^43–46^. Although it is not possible to make direct comparisons between the VE using the two intervals between different countries, we did explore this hypothesis in a sensitivity analysis, and obtained similar findings. Second, we assume a simple model of waning immunity with only two stages. However, sensitivity analyses where we varied assumptions regarding waning of vaccine-induced protection produced results consistent with our main analysis. Third, we do not directly account for waning of vaccine-induced immunity after the first dose. Studies have estimated that protection starts to wane from approximately 4-5 weeks after first dose vaccination in some age groups but with large uncertainty ^27,38,47^. We performed a sensitivity analysis demonstrating that such waning after first dose did not alter our results. Fourth, although we capture changes in the overall level of mixing over time, we assumed that under the 3-week strategy mixing patterns remained identical to that estimated under the 12-week strategy. In reality, population-based behaviours and risk perception changes as case numbers or hospitalisations increase ^48^. It is possible that, under a 3-week strategy, the last step of the roadmap would have been delayed further as the projected hospitalisations would have been much higher (figure 2B); it is also possible that behaviour would have been different under that strategy. Therefore, our counterfactual analysis can only represent a scenario assuming only the vaccination interval changed. Additionally, we did not consider the potentially reduced risk of VOC emergence in the 3-week, compared to the 12-week strategy. However, with SARS-CoV-2 circulating globally, and most VOCs rapidly spreading worldwide, it is unlikely that a change in the risk of emergence in England alone would have affected the broader patterns of emergence and therefore the circulation of variants.

Finally, although we have quantified the relative success of the 12-versus 3-week strategy in terms of COVID-19 infections, hospitalisations, and deaths, we have not quantified specifically the long-term burden of disease, for example due to long COVID, although this is likely to be proportional to the burden of SARS-CoV-2 infections.

In line with previous studies, we find that a key aspect of England’s successful vaccination strategy was to rapidly provide partial vaccine-induced protection to a larger proportion of younger age groups who may be less vulnerable to severe disease but may contribute more to transmission, thus indirectly protecting the most vulnerable groups ^41,49^. This successful switch to a 12-week interval between COVID-19 vaccine doses was informed by early real-world VE estimates in the UK, careful assessment of the trade-offs in the context of limited vaccine supply in a growing epidemic, and limited 12-week efficacy data from the AstraZeneca vaccine trial. Our study shows that early and continuous assessment of real-world vaccine effectiveness is crucial especially in emergency situations. There is benefit in carefully considering and adapting guidelines in light of new emerging evidence and the population in question.

It is difficult to extrapolate findings from England, where vaccine roll-out started whilst strict NPIs were also maintained, to other settings due to differences in demography, behaviours, implementation of NPIs, availability of vaccines, and VOCs in circulation. However, across all sensitivity analyses explored, we estimated that in England, the decision to switch to a delayed vaccine interval was beneficial. Importantly, this beneficial effect is observed across all age groups, including the most vulnerable ones, who had higher indirect protection under the 12-week strategy.

It was fortunate that both vaccines licensed in England were highly effective, combined with a very high uptake of both first and second vaccine doses despite the longer interval between doses. In other settings where vaccine supply is limited, or vaccines with a lower effectiveness are widely used, a switch to a 12-week strategy may not be advantageous. In this context, it is crucial to consider the likely acceptance of new vaccines and/or further booster doses in the population, and potential barriers to uptake especially during a period of sustained high transmission as there is a risk of vaccine-resistant variants emerging. Models have shown that the long-term trajectory of COVID-19 is strongly shaped by vaccination policies with wide-ranging epidemiological and evolutionary outcomes ^50^. Therefore, as booster and fourth vaccines doses are rolled out, it will be important to continue evaluating VE against any new variants and waning vaccine-induced protection.

Beyond the immediate priority of COVID-19, understanding the impact that changing COVID-19 vaccination guidance has had or may have on hesitancy towards other vaccines as well as overall trust in public health guidance is crucial.

## Data Availability

All data files and source code required to reproduce this analysis are publicly available at https://github.com/mrc-ide/sarscov2-roadmap-england

## Declaration of interests

AC has received payment from Pfizer for teaching of mathematical modelling of infectious diseases. KAMG has received honoraria from Wellcome Genome Campus for lectures. LKW has received consultancy payments from the Wellcome Trust. ABH has received consultancy payments from the World Health Organization for COVID-19 related work. ABH provides COVID-19 modelling advice to the New South Wales Ministry of Health, Australia. ABH was previously engaged by Pfizer Inc to advise on modelling RSV vaccination strategies for which she received no financial compensation. RS is currently employed by the Wellcome Trust, however the Wellcome Trust had no role in the study design, data collection, data analysis, data interpretation, or writing of the report.All other authors declare no competing interests.

## Funding

This work was supported jointly by the Wellcome Trust and the Department for International Development (DFID; 221350). We acknowledge joint Centre funding from the UK MRC and DFID (MR/R015600/1). This work was also supported by the NIHR Health Protection Research Unit in Modelling Methodology, a partnership between UKHSA, Imperial College London, and London School of Hygiene & Tropical Medicine (NIHR200908), the Abdul Latif Jameel Foundation, and the EDCTP2 programme supported by the European Union (EU). LKW acknowledges support from a Wellcome Trust Fellowship (grant number 218669/Z/19/Z). ABH acknowledges support from an NHMRC Investigator Grant.

For the purpose of open access, the author has applied a ‘Creative Commons Attribution’ (CC BY) licence to any Author Accepted Manuscript version arising from this submission.

## Acknowledgments

We thank all colleagues at the UK Health Security Agency (UKHSA, formerly Public Health England) and front-line health professionals who have not only driven and continue to drive the daily response to the COVID-19 epidemic in England but also provided the necessary data to inform this study. This work would not have been possible without the dedication and expertise of said colleagues and professionals. The use of pillar-2 PCR testing data, vaccination data, and the variant and mutation data was made possible thanks to UKHSA colleagues, and we extend our thanks to N Gent and A Charlett for facilitation and insights into these data. The use of serological data was made possible by colleagues at UKHSA Porton Down, Colindale, and the NHS Blood Transfusion Service. We are particularly grateful to G Amirthalingam and N Andrews for helpful discussions around these data. We thank all the REal-time Assessment of Community Transmission (REACT) Study investigators for sharing PCR prevalence data. We also thank the entire Imperial College London COVID-19 response team for support and feedback throughout. The views expressed are those of the authors and not necessarily those of the UK Department of Health and Social Care, the NHS, the National Institute for Health Research (NIHR), UKHSA, UK Medical Research Council (MRC), UK Research and Innovation, or the EU.

## Notes

### Summary of Updates

Corrected author affiliation for Raphael Sonabend.

