## Supplementary Material for "Quantifying the impact of delaying the second COVID-19 vaccine dose in England: a mathematical modelling study"

Symbol Glossary 71

List of Figures 73

List of Tables 74

### 1 Model Description and Definitions

In this section we provide a brief description to our model along with key definitions. Full details about the fitting procedure, parameter assumptions, and model equations are provided in Section 4.

#### 1.1 Overview of the model

We used a discrete-time stochastic compartmental model of SARS-CoV-2 transmission, illustrated in Figure S1, which has previously been described in detail in Knock et al. [1] and Sonabend et al. [2]. The model is an extended SEIR-type model, stratified into 16 five-year age groups (0-4, 5-9, ..., 75-79), 80+, a group of care home residents (CHR) and a group of care home workers (CHW). Mixing between these groups is informed by survey data [3]. Upon infection with SARS-CoV-2, individuals enter an exposed compartment, before becoming infectious. A proportion of infectious individuals are assumed to develop symptoms, while the rest remain asymptomatic. All asymptomatic cases and a fraction of symptomatic cases recover naturally, while the rest of the symptomatic cases develop severe disease requiring hospitalisation. Of these, a proportion die at home, while the remainder are admitted to hospital. Hospital pathways are described in detail, with patients being either triaged before intensive care unit (ICU) admission, then admitted to ICU, before being transferred into general wards for stepdown recovery, or remaining in general beds throughout. Hospitalised cases are either confirmed as SARS-CoV-2 cases upon admission or may be tested and confirmed later during their stay.

Each compartment in the model is further stratified to account for vaccination status. We used four vaccination strata Figure S1) and (Table S2. These describe the recommended two-dose vaccination regimen (common to the three double-dose vaccines licensed and available for use in England during the study period: Oxford-AstraZeneca ChAdOx1 nCoV-19 (AZD1222) [4], Pfizer-BioNTech COVID-19 Vaccine BNT162b2 [5], and Moderna mRNA-1273 [6], henceforth referred to as AZ, PF, and Mod respectively), capturing a delay between receiving a dose and the onset of dose-specific effectiveness, as well as waning of vaccine effectiveness post second dose. Polymerase chain reaction (PCR) tests and serology status are modelled with parallel flows.

The model was extended to include the spread of variants of concern (VOC). In the context of this paper, we consider Alpha (B.1.1.7) coexisting with the Delta variant (B.1.617.2). All references to 'Alpha' here refer to the Alpha variant and all other previously circulating variants. We fit a two-variant model, with Delta seeded at a region-specific date determined by the model fit, no earlier than 8th March 2021.

The study period considered is 8th December 2020 to 13th September 2021, capturing the window between when the UK vaccination campaign began, up until the start of the third dose ("booster") campaign. This time period captures the emergence and establishment of the Delta variant, but ends before the introduction of the Omicron variant (B.1.1.529) which was first reported in the UK on 27th November 2021 [7]. Thus, we do not consider the Omicron variant.

We account for waning of infection-induced immunity in the model (for waning of vaccine-induced immunity see Section 3.2). Individuals who have recovered from SARS-CoV-2 infection are protected against reinfection with the same variant for an exponentially distributed duration with mean 6 years [8], after which they move back to the susceptible compartment. Further, we model asymmetrical cross-immunity between SARS-CoV-2 variants (Section 2.5). We will use the term 'susceptible' only to refer to individuals in compartment '$S$', whereas 'uninfected' will refer to those in compartment '$S$' or '$R$' (recovered).

Similarly to Knock et al. [1] and Sonabend et al. [2], the model is used in two stages: an initial model fitting stage to build posterior estimates of model parameters fitting to multiple epidemiological data streams, followed by a simulation stage, whereby the posterior estimates inform counterfactual "what if" scenarios.

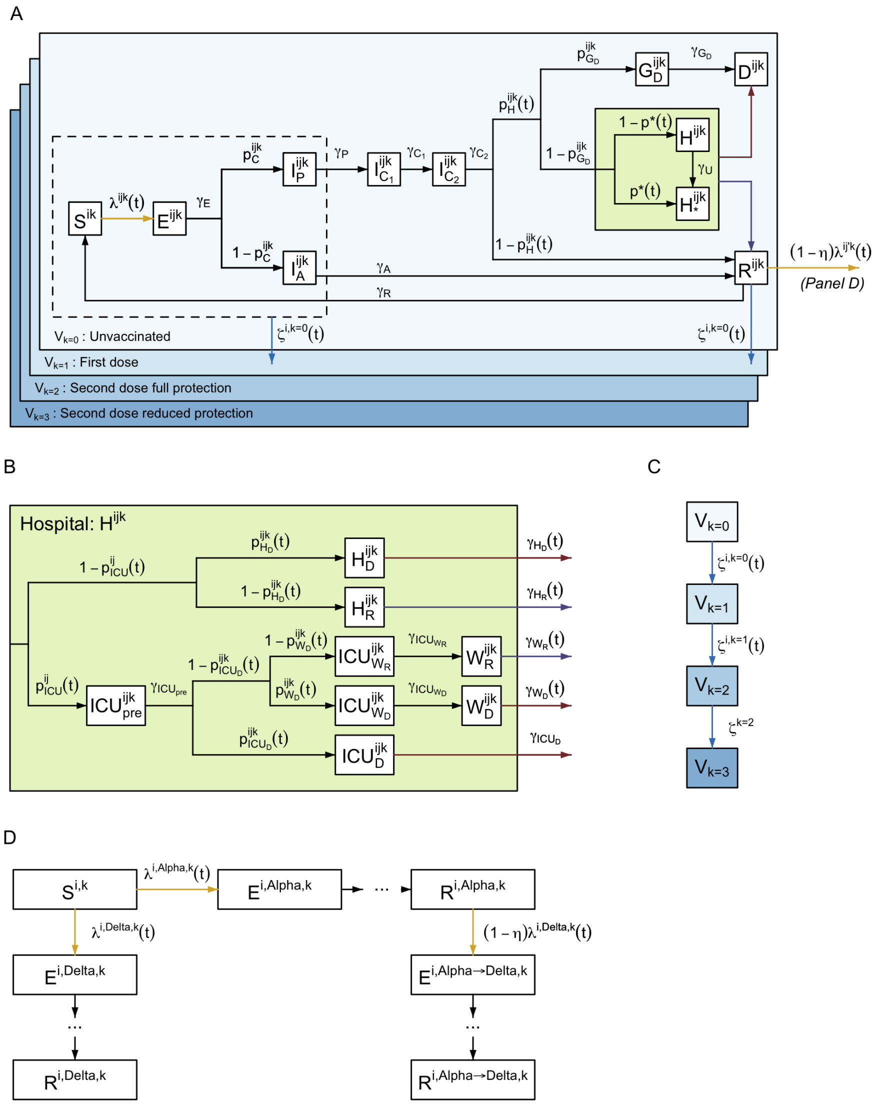

**Figure S1:** Model structure flow diagram with rates of transition between states. (A) Extended SEIR transmission model flow diagram overview. (B) Hospital flow diagram. (C) Vaccination flow diagram. (D) Multi-variant flow diagram showing possible infection with *Alpha*, *Delta*, or both in turn (*Alpha*→*Delta*). All variables and parameters defined in tables S5, S6, and S7. Superscripts refer to the age or care home group ($i \in [0-4], [5-9], \ldots, [75-79], [80+], CHW, CHR$), variant ($j = Alpha, Delta$, or $Alpha$→$Delta$), and vaccination stratum ($k = 0, \ldots, 3$) see table S2.

#### 1.2 Reproduction number

We use two definitions of the reproduction number throughout. We denote $R^j(t)$ as the reproduction number for variant $j$ ($j = Alpha, Delta$) in the absence of immunity at time $t$. This is defined as the average number of secondary infections that an individual infected at time $t$ with variant $j$ would generate in an entirely susceptible and unvaccinated population. In contrast, the effective reproduction number, $R_e^j(t)$, for variant $j$ at time $t$ is the number of secondary infections in the actual population, accounting for immunity (natural and vaccine-induced) present at that time in the population. Hence, by definition, $R_e^j(t) \leq R^j(t)$.

#### 1.3 Fitting to data

The model is fitted to multiple data streams from each National Health Service (NHS) region in England, as described in Sonabend et al. [2]; this is summarised in Table S1.

| Data | Description | Source | Reference |
|---|---|---|---|
| Hospital deaths | Daily number of COVID-19 hospital deaths reported by NHS England within 28 days of a COVID-19 positive result | Public Health England (PHE) | These data underlie the Gov.uk dashboard data [9] |
| Care home deaths | Daily number of deaths with COVID-19 mentioned as a cause on the death certificate and "care home", "hospice" or "other institution" as the place of death | ONS | These data underlie the Gov.uk dashboard data [9] |
| Community deaths | Daily number of deaths with COVID-19 mentioned as a cause on the death certificate and any place of death that is not one of "hospital", "care home", "hospice" or "other institution" | ONS | These data underlie the Gov.uk dashboard data [9] |
| ICU occupancy | Daily number of confirmed COVID-19 patients in ICU | Gov.uk Dashboard | [9] |
| General bed occupancy | Daily number of confirmed COVID-19 patients in non-ICU beds | Gov.uk Dashboard | [9] |
| Admissions | Daily number of confirmed COVID-19 patients admitted to hospital | Gov.uk Dashboard | [9] |
| Pillar 2 testing | Daily number of positive (cases) and negative PCR test results for individuals aged 25 or over | PHE | These data underlie the Gov.uk dashboard data [9] |
| REACT-1 testing | Real-time Assessment of Community Transmission (REACT) daily number of positive and negative PCR test results | REACT | [10] |
| Serology | Serology survey conducted on blood donors aged 15-65. Results from the EuroImmun and Roche N assays are used, fitting to each assay separately. EuroImmun results are only used up to (and including) 14th January 2020 | PHE | These data are collected as part of [11]. |
| Vaccinations by age | Daily number of first and second-vaccine doses - reported in 5-year age groups | PHE | [9] |
| Variant and Mutation | Daily number of variant tests by NHS region identified as Delta or Alpha (or non-Delta variants) | VAM | These data underlie the Gov.uk genomic surveillance reports [12] |

**Table S1:** Data sources and definitions. All data are reported by NHS region or processed to match these regions

### 2 The Delta Variant

For model fitting, we switch from a one-variant to a two-variant model on 8th March 2021 to capture the emergence and spread of Delta in all NHS regions of England. For the counterfactual analysis which starts before the introduction of the Delta variant, we use a two-variant model throughout, but only seed the Delta variant from the 8th March 2021 where region specific seeding dates are estimated. Key differences between the modelled variants are summarised in the next subsections, these are:

1. Dates of introduction.
2. Differences in transmissibility between variants
3. Differences in vaccine effectiveness against Alpha and Delta for various endpoints
4. Differences in severity between variants
5. Asymmetric cross immunity between the variants.

#### 2.1 Seeding the epidemic and the Delta variant

We seed each variant $j$ at a daily rate $\phi_j$, over a period of $\nu_j$ days from time $t_j$. All seeding infections are from the S to E compartment in the 15-19 year old group and unvaccinated class.

For the Alpha variant we seed at a rate of 10 per million of the total regional population per day, over a 1-day period, so $\phi_{Alpha} = \frac{1}{100,000}\sum_i N^i$ and $\nu_{Alpha} = 1$.

For the Delta variant we seed at a rate of 2 per million of the total regional population per day, over a 7-day period, so $\phi_{Delta} = \frac{1}{500,000}\sum_i N^i$ and $\nu_{Delta} = 7$.

The seeding dates $t_{Alpha}$ (which corresponds to the start date of the regional epidemic) and $t_{Delta}$ are fitted parameters (see Table S8).

The seeding rate in group $i$ ($i$ referring to age or care home group) and vaccine stratum $k$ of variant $j$ is then given by

$$\delta^{i,j,k}(t) = \begin{cases} \phi_j & \text{if } i = [15,20),\ j \in \{Alpha,\ Delta\},\ k = 0 \text{ and } t_j \leq t < t_j + \nu_j \\ 0 & \text{otherwise.} \end{cases} \tag{1}$$

Where$\delta^{i,j,k}(t)$ is the daily seeding rate of variant $j$ (stratified by age and vaccination strata).

#### 2.2 Transmissibility of Delta compared to Alpha

We fit a region-specific transmission advantage, $\sigma$, of the Delta variant compared to Alpha. We use a uniform prior between 0 and 3 (Table S8), and an initial value of 1 for the pMCMC chains. In the absence of strong evidence for differences in the generation interval of Alpha and Delta [13, 14], we assume the same generation interval for all variants in the model.

#### 2.3 Vaccine effectiveness against Delta

In addition to the intrinsic transmission advantage considered for Delta, we assume a reduction in vaccine effectiveness against Delta compared to Alpha. Both the transmission advantage, $\sigma$ (which is estimated), and variant-specific vaccine effectiveness (which are fixed) are captured in the force of infection (see Section 4.3). For full details on vaccine effectiveness against Delta, see Section 3 and in particular Table S3.

#### 2.4 Increased severity of Delta compared to Alpha

To account for the potentially increased severity of Delta relative to Alpha and previously circulating variants [15], we allow for fitted multipliers for probability of hospitalisation ($\pi_H^{Delta}$), admission to ICU ($\pi_{ICU}^{Delta}$) and death ($\pi_D^{Delta}$) upon infection with the Delta variant relative to the Alpha variant (see section 4.3.2). After application of multipliers, probabilities are capped at 1.

#### 2.5 Protection from previous infection

The level of cross-protection against Delta from prior infection with non-Delta variants is difficult to quantify, but in-vitro neutralisation studies found Delta was neutralised to a lesser extent by antibodies from previous infections [16]. We model asymmetric cross immunity between the two variants and assume that infection with Delta confers perfect immunity to infection with Alpha, whilst infection with Alpha is only partially protective against infection with Delta (see Table S7). In addition, for individuals infected by Delta following an infection with Alpha ($E^{i,Alpha \to Delta,k}$), we assume that, if the second infection is symptomatic, the probability of hospitalisation is reduced compared to individuals with no prior infection history. We consider this relative reduction equivalent to the conditional effectiveness against severe disease conditional on being symptomatic afforded by one dose of PF against Delta ($e_{SD|sympt}$ in Table S4 below) and as estimated by Kim et al [17]. Probability of infection or hospitalisation by either variant "resets" to baseline assumptions once an individual's infection-induced protection wanes and they re-enter the susceptible compartment, $S^{i,k}$.

### 3 Vaccination

The Medicines and Healthcare Products Regulatory Agency issued temporary authorisation grants for both the AZ and PF vaccines in December 2020 [4, 5], and approved Mod vaccine shortly after in January 2021 [6]. All three vaccines require two doses to be administered, with increasing levels of vaccine effectiveness (VE) seen after each dose. Waning in VE is observed following doses being administered. The study period considered is before the rollout of a third, "booster", dose, and as such third doses are not considered in this study. Thus, our model considers four distinct vaccination strata ($V_k$, for $k \in \{0,1,2,3\}$) representing the four stages of VE detailed in Table S2 and illustrated in Figure S2.

| Vaccination stratum name | Number of doses | Vaccine effectiveness for that group | Description | References |
|---|---|---|---|---|
| $V_0$ | 0 | None | Non-vaccinated individuals or those less than 3 weeks since first vaccination. | Section 3.4 |
| $V_1$ | 1 | First dose effectiveness | Individuals are partially protected 3 weeks from date of first vaccination. | [18, 19] |
| $V_2$ | 2 | Full second dose effectiveness | Individuals are fully protected 1 week from date of second vaccination. In counterfactual simulations we assume date of second vaccination is 3 weeks after date of first vaccination. | [20, 18, 19] |
| $V_3$ | 2 | Reduced second dose effectiveness | Individuals with reduced second dose vaccine protection; transition from $V_2$ is randomly drawn from an exponential distribution with mean waning time of 24 weeks. | Section 3.2 |

**Table S2:** Vaccination strata considered for individuals, corresponding schedule, and vaccine effectiveness at each stage.

Individuals in our model move out of an unvaccinated ($V_0$) stratum at a rate determined by vaccine roll-out data and the prioritisation strategy adopted by the UK government (Section 3.4). We only allow vaccination of individuals who are not symptomatic and not hospitalised, i.e. only individuals in the following compartments can be vaccinated: susceptible ($S$), exposed ($E$), infected asymptomatic ($I_A$), infected pre-symptomatic ($I_P$) or recovered ($R$). Other compartments are also stratified by our four vaccination strata but for those there is no movement between vaccine strata ($V_k$).

Phase 2 trials for the AZ and PF vaccines suggested substantial increases in immunogenicity started approximately two to three weeks after receiving the first dose [18, 19]. We therefore assumed a 21-day delay between first dose injection and onset of dose-specific effectiveness. In our model, after receiving their first dose, individuals remain in $V_0$ for 21 days on average, during which the vaccine offers no protection. They then move on to the $V_1$ strata, where they are protected with one dose VE and stay until the protection from the second dose kicks in. After receiving their second dose, individuals remain in $V_1$ for 7 days on average [18], until the second dose reaches maximum effectiveness, at which point individuals move on to the $V_2$ stratum where they are assumed to achieve maximal protection offered by the two doses. This is illustrated in Figure S2.

Our study compares two scenarios investigating England's vaccination strategy decision to delay the second dose to, on average, 12 weeks after the first dose [20] compared to the initial 3 week interval strategy. In our fits, we use actual data on the number of first and second doses given each day in each age group and region, corresponding to an average delay of 12 weeks between doses. In our counterfactual analyses however, we assume a three week interval between doses. We assume that the total daily number of doses by region per day remains the same as the data; only the daily split between first and second doses is modified, to ensure second doses are delivered in priority, three weeks after the first dose (see section 3.4).

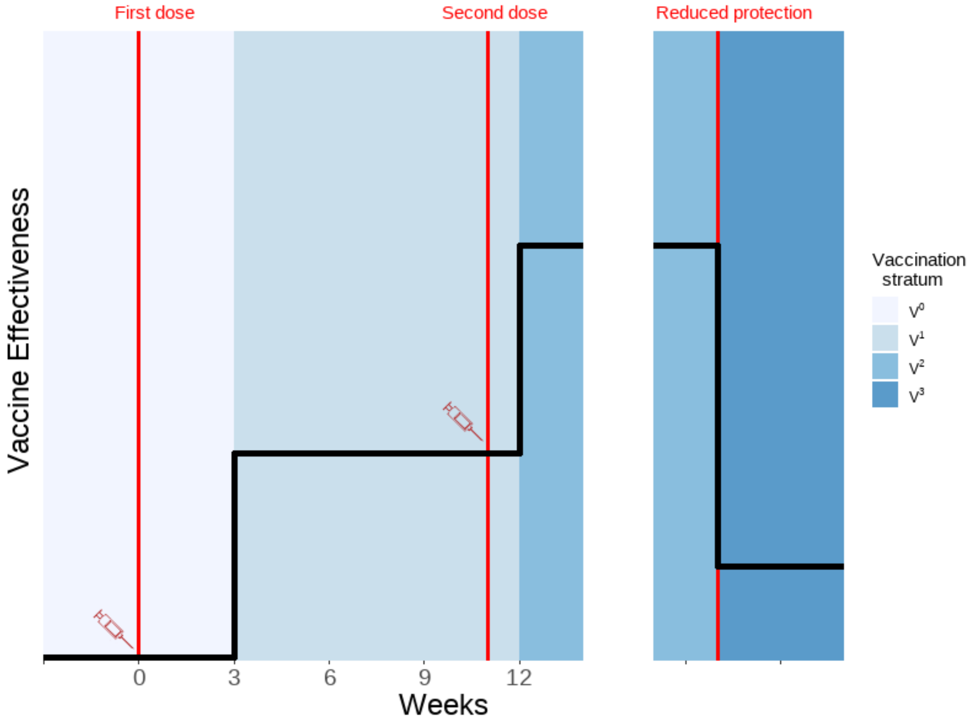

**Figure S2:** Vaccination strata duration and associated illustrative vaccine effectiveness. Red lines depict points at which a vaccine dose is administered. We assume an average 24-weeks to waning. Y-axis illustrates changing vaccine effectiveness. Vaccination strata are defined in Table S2.

#### 3.1 Vaccine effectiveness

The assumed values for vaccine effectiveness (VE) are derived from both vaccine efficacy measured in clinical trials and vaccine effectiveness studies. Where possible, data from the UK have been used and represent effectiveness of dosing schedules with an 12 week gap between doses. We assumed that there are no significant differences in vaccine effectiveness by age, sex, or underlying health conditions [21, 22]. Table S3 summarises our vaccine effectiveness assumptions for the PF, AZ, and Mod vaccines. We assume that vaccine protection against symptomatic disease, as determined from the original trials and real-world data, also provides a similar level of protection against infection. We further assume that, in those individuals who do become infected after vaccination, onward transmission is also reduced [23]. Finally, we incorporate a higher overall level of vaccine effectiveness against severe disease and against death. We enforce that all Delta VE estimates are capped by the associated Alpha VE estimate.

For our main analysis, we assume VE remains the same regardless of the dosing interval, summarised in Table S3. We then run a number of sensitivity analyses to explore:

1. uncertainty in VE when a 3-week rather than 12-week interval is used following Amirthalingam et al. [24]
2. uncertainty in VE against the Delta variant

3. uncertainty in the level of waning vaccine-induced immunity
4. uncertainty in the timing of vaccine-induced waning immunity
5. uncertainty in waning of immunity following the first vaccine dose.

See Section Section 5 for details. Table S9 summarises the 9 analyses (including our main assumptions) explored, and Table S10 and table S11 summarises the VE assumptions used for each analysis against the Alpha and Delta variant respectively.

| | | Alpha | | Delta | | Informed by |
|---|---|---|---|---|---|---|
| End point | Dose | AZ | PF/Mod | AZ | PF/Mod | |
| Death | 1 | 88% | 89% | 87% | 89% | [25, 26] |
| | 2 (Full protection) | 99% | 99% | 99% | 99% | [25, 26] |
| | 2 (Reduced protection) | 83% | 90% | 82% | 90% | [27, 28, 29] |
| Severe disease | 1 | 81% | 89% | 81% | 89% | [30, 31, 32] |
| | 2 (Full protection) | 99% | 99% | 99% | 99% | [33, 34] |
| | 2 (Reduced protection) | 77% | 90% | 77% | 90% | [27, 28, 29] |
| Mild disease or infection | 1 | 64% | 79% | 51% | 51% | [15, 31, 33, 34, 21, 35] |
| | 2 (Full protection) | 92% | 99% | 87% | 95% | [15, 34, 21, 35, 36, 37] |
| | 2 (Reduced protection) | 29% | 77% | 19% | 49% | [27, 28, 29] |
| Transmission | 1 | 45% | 45% | 33% | 33% | [25] |
| | 2 (Full protection) | 45% | 45% | 40% | 40% | [25] |
| | 2 (Reduced protection) | 40% | 40% | 19% | 19% | [27, 28, 29] |

**Table S3:** Vaccine effectiveness assumptions for AstraZeneca (AZ), Pfizer (PF), and Moderna (Mod) by vaccine dose. For our baseline analysis, we assume that VE for the 12-week and 3-week dosing intervals remain the same. See Table S10 and table S11 for full VE assumptions for each sensitivity analysis

We model cases that require hospitalisation and are hospitalised, as well as cases that require hospitalisation but are not hospitalised; for this reason we refer to vaccine effectiveness against *severe disease* and not *hospitalisation*. Vaccine effectiveness against severe disease, conditional on symptoms, acts on transition to both this compartment of individuals and those admitted to hospital.

We do not model individual vaccines separately, instead vaccine compartments are type-agnostic, and for vaccine effectiveness we compute an age-dependent weighted mean of each vaccine's effectiveness (where weights for a given age group are the proportion of each vaccine type administered to that age group as of 13th September 2021). Whilst we assume vaccine effectiveness does not vary by age, the proportion of each vaccine (PF, AZ or Mod) administered to each age group varied substantially (Figure S3) and vaccine effectiveness varies between vaccines (Table S3), therefore our weighted vaccine-effectiveness varies by age.

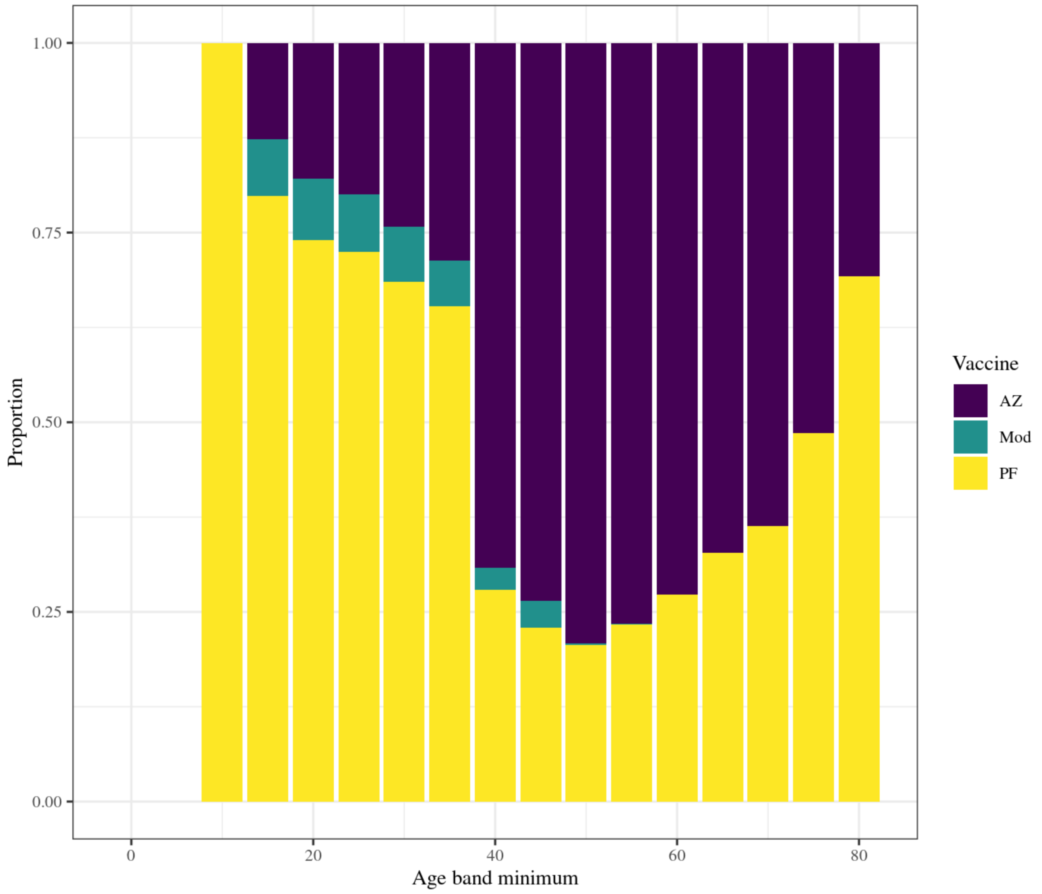

**Figure S3:** Proportion of each vaccine type: (Oxford-AstraZeneca (AZ), Pfizer-BioNTech (PF), Moderna (Mod)) dispensed to each five-year age band as of 13th September 2021. Data taken from UK Health Security Agency Immunisations database for vaccine delivery and ONS population estimates for each age group.

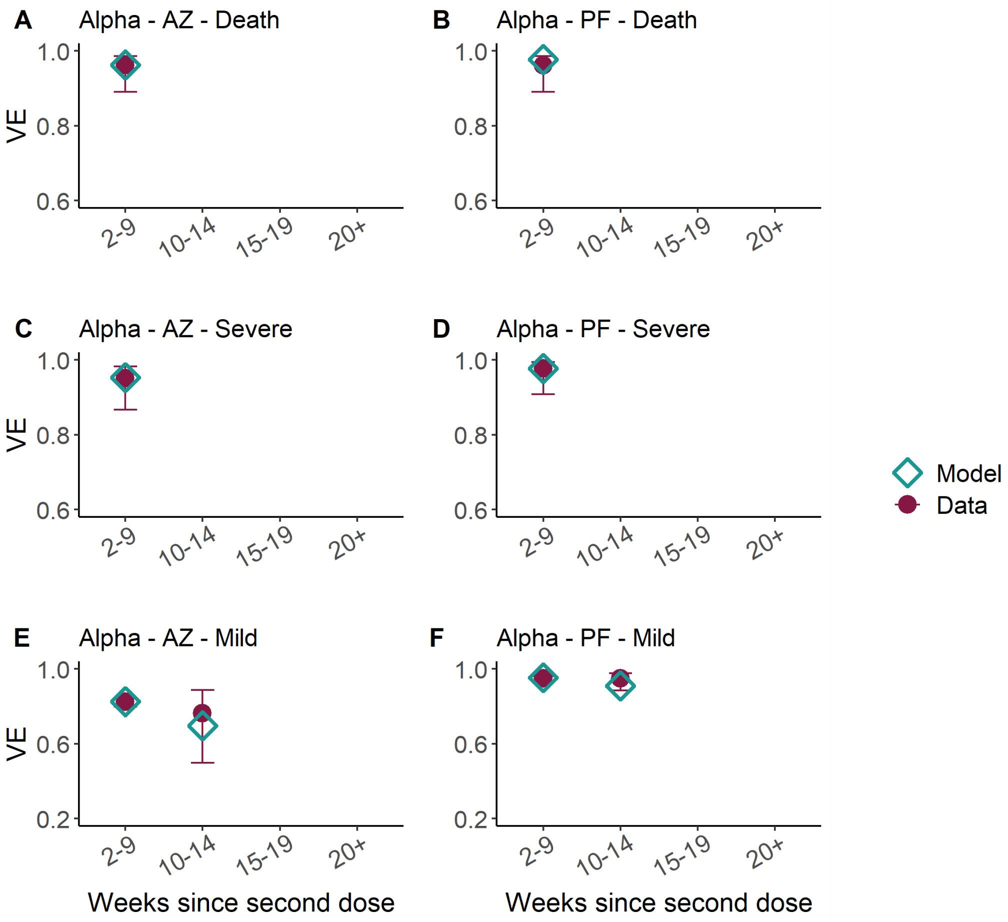

**Figure S4:** Vaccine effectiveness in weeks since second dose of AstraZeneca (AZ, left column) and Pfizer (PF, right column) vaccines against Alpha for death (top), severe disease, (middle) and mild disease/infection (bottom). We assume the same protection against infection and mild disease. Turquoise points show the VE estimates from Andrews et al, and the purple points our model assumptions. We assumed that the Moderna vaccine has the same VE as PF.

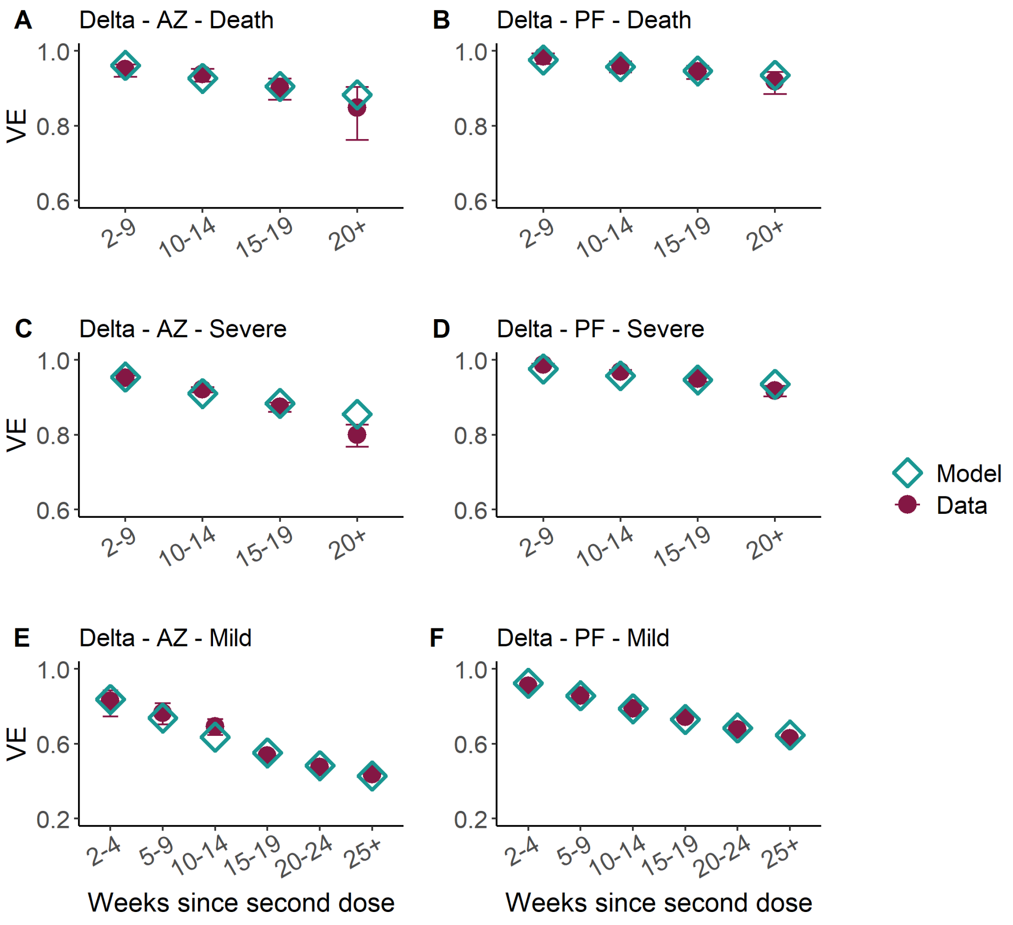

**Figure S5:** Vaccine effectiveness in weeks since second dose of AstraZeneca (AZ, left column) and Pfizer (PF, right column) vaccines against Delta for death (top), severe disease, (middle) and mild disease/infection (bottom). We assume the same protection against infection and mild disease. Turquoise points show the VE estimates from Andrews et al, and the purple points our model assumptions. We assumed that the Moderna vaccine has the same VE as PF.

#### 3.2 Waning of vaccine-induced immunity

Population-level vaccine effectiveness is observed to gradually wane after second doses are administered [29]. Our compartmental model assumes that, upon receiving their second dose, individuals firstly progress to the $V_2$ vaccination strata, granting full second dose vaccine protection. Eventually, individuals will then progress to a "reduced protection" strata, $V_3$, providing a lower degree of vaccine effectiveness, as illustrated in Figure S2. The duration an individual spends in the $V_2$ strata is stochastic, with an individual's rate of progression drawn from an exponential distribution with a mean $V_2$ duration of 24 weeks. Note that this is a simplification of biological reality necessitated by the compartmental structure of our model. In actuality, the waning of VE for an individual occurs gradually (and continuously) over time [27] and at different waning rates.

Heuristically, if 1,000 individuals received their AZ second dose on day 0, they would all start in the $V_2$ strata, and hence all with VE 0.99 (Table S3). After 20 weeks, following the exponential distribution, 57% ($P(X \geq 20 \text{ weeks}), X \sim exp(\frac{1}{24 \text{ weeks}})$) of those 1,000 individuals will have waned to the $V_3$ strata. Therefore, 570 individuals would have VE 0.82 protection against death, while 430 would remain in the $V_2$ strata with VE 0.99 protection against death - resulting in a population mean VE protection against death of 0.89. Andrews et al. (2021) [29] report population average vaccine effectiveness for 20+ weeks as 84.8 (76.2–90.3) 95% CI.

To ensure our waning mechanism closely mirrored the average level of vaccine waning actually observed in the population, we fit second dose full protection ($V_2$), and reduced protection compartment ($V_3$) VE values for all vaccines and health outcomes such that the mean VE for an individual at any time best replicates real world data. As such, we fit to the time-varying VE data presented in Andrews et al. (2021) [29], for all vaccines and health outcomes, save for VE against mild disease for the Delta variant, where we fit to the expanded data provided in Andrews et al. (2022) [38].

Due to the limited data available for waning effectiveness against the Alpha variant, we choose to fix the log-odds difference between full second dose VE and reduced second dose VE to be the same for each variant, assuming the same proportion of protection drop-off from their initial full second dose VE for each variant. Mathematically, if $V(x)$ represents the VE of vaccine/health outcome $x$, then we define the log odds as $L(x) = \log\left(\frac{V(x)}{1-V(x)}\right)$. Thus, we fix that $L$(Reduced second dose VE vs. Delta) $-$ $L$(Full second dose VE vs. Delta) $=$ $L$(Reduced second dose VE vs. Alpha) - $L$(Full second dose VE vs. Alpha) for each health outcome. Andrews et al. provide VE estimates for timeframes of multiple weeks, against which we compared the mean population average VE from our continuous model output for the timeframe indicated. For the 20+ weeks data we compared against our model average between 20 and 29 weeks, and for the 25+ data we compared against the average between 25 and 30 weeks.These averages were fit to the data via weighted least squares, using 1/width of the associated 95% CIs reported with the data as the weights.

For first dose VE estimates we assumed the 16+ age group Delta VE values as presented in supplementary table S6 of Andrews et al. (2021) [29], however due to prioritised groups being vaccinated during the Alpha wave, the reported VEs are unlikely to be generalisable to the public at large, and as such we assumed first dose VE estimates for Alpha such that $L$(First dose VE vs. Alpha) $-$ $L$(Full second dose VE vs. Alpha) $=$ $L$(First dose VE vs. Delta) - $L$(Full second dose VE vs. Delta).

#### 3.3 Conditional dependencies of vaccine-immunity

We present unconditional VE in Table S3 however our model is framed as a compartmental cascade of symptom severity, hence we convert unconditional effectiveness to conditional as detailed in Table S4.

| VE vs. | Symbol / Calculation |
|---|---|
| Infection | $e_{inf}$ |
| Symptoms | $e_{sympt}$ |
| Severe disease | $e_{SD}$ |
| Death | $e_{death}$ |
| Symptoms given infection | $e_{sympt\|inf} = \frac{e_{sympt} - e_{inf}}{1 - e_{inf}}$ |
| Severe disease given symptoms | $e_{SD\|sympt} = \frac{e_{SD} - e_{sympt}}{(1 - e_{inf})(1 - e_{sympt\|inf})}$ |
| Death given severe disease | $e_{death\|SD} = \frac{e_{death} - e_{SD}}{(1 - e_{inf})(1 - e_{sympt\|inf})(1 - e_{SD\|sympt})}$ |

**Table S4:** Conditional vaccine effectiveness values that we model.

#### 3.4 Vaccine roll-out

The Joint Committee on Vaccination and Immunisation (JCVI) established an ordered list of individuals prioritised for vaccination in the UK, first prioritising care home residents and care home workers, and then other adults by decreasing age and clinical vulnerability [39, 40].

We assume first and second doses were delivered in England between 8th December 2020 and 13th September 2021 as reported in age-stratified data received from UK Health Security Agency (UKHSA) and the Department of Health and Social Care (DHSC) via the Scientific Pandemic Influenza Group on Modelling (SPI-M) (Figure S6) as reported on the COVID-19 dashboard [9]. It is not possible to identify individuals in our CHR and CHW groups from the data. However, as these are the top two priority groups, we assume that all individuals vaccinated under 65 years were carehome workers and all individuals 65 years or over were carehome residents, up until an uptake of $95\% and 85\%$ is reached in CHR and CHW respectively [41].

In our counterfactual simulation we assume that the same number of vaccine doses per day and region as reported in the data was administered. However, these were split between first and second doses so that individuals who have already received one dose receive their second dose approximately 3 weeks after their first.

The UK government initially advised a 12-week prime-boost interval as supported by vaccine trials [36]. This was changed to a 8-week delay in mid-May 2021 to help combat the spread of Delta [42]. Initially altered only for those over the age of 50, this 8-week interval was expanded to the whole population in June 2021 (see Figure S6).

### 4 Model Parameterisation and Fitting

#### 4.1 Model compartments and parameters

In the following, $i$ denotes the age or care home group of individuals ($i = [0,5),[5,10),\ldots,[75-80),[80+),$ $CHW,CHR$), and $j$ denotes their variant status ($j = Alpha$ for infection with Alpha or previous variants, $Delta$ for infection with the Delta variant, or $Alpha \rightarrow Delta$ if an individual is infected with Delta immediately following infection from Alpha). Finally, $k$ denotes the index of the vaccination stratum of individuals (with $k$ corresponding to $V_k$ as defined in Table S2).

$\zeta^{i,k}(t)$ is the rate of movement from vaccination stratum $k$ to vaccination stratum $k+1$ at time $t$, for individuals in group $i$. For $k = 0$ and $k = 1$, this was set to match the observed number of daily first and second doses aimed to be given to each group at time step $t$. For $k = 2$, this was set so that the average time to waning of the second dose vaccine effectiveness was 24 weeks (see section 3.2). Note that there is no movement out of vaccination stratum 3, so by definition $\zeta^{i,3}(t) = 0$. Additionally we have no movement into vaccination stratum 0, so for ease of notation and equation simplicity we let $k = -1$ be a dummy vaccination stratum with empty compartments and let $\zeta^{i,-1}(t) = 0$.

We define all model compartments and parameters in Table S5 below, and illustrate the model structure and flows between compartments in Figure S1 (this figure is copied again below in Figure S7 for easy reference). The model assumes discrete time and four time steps are taken per day.

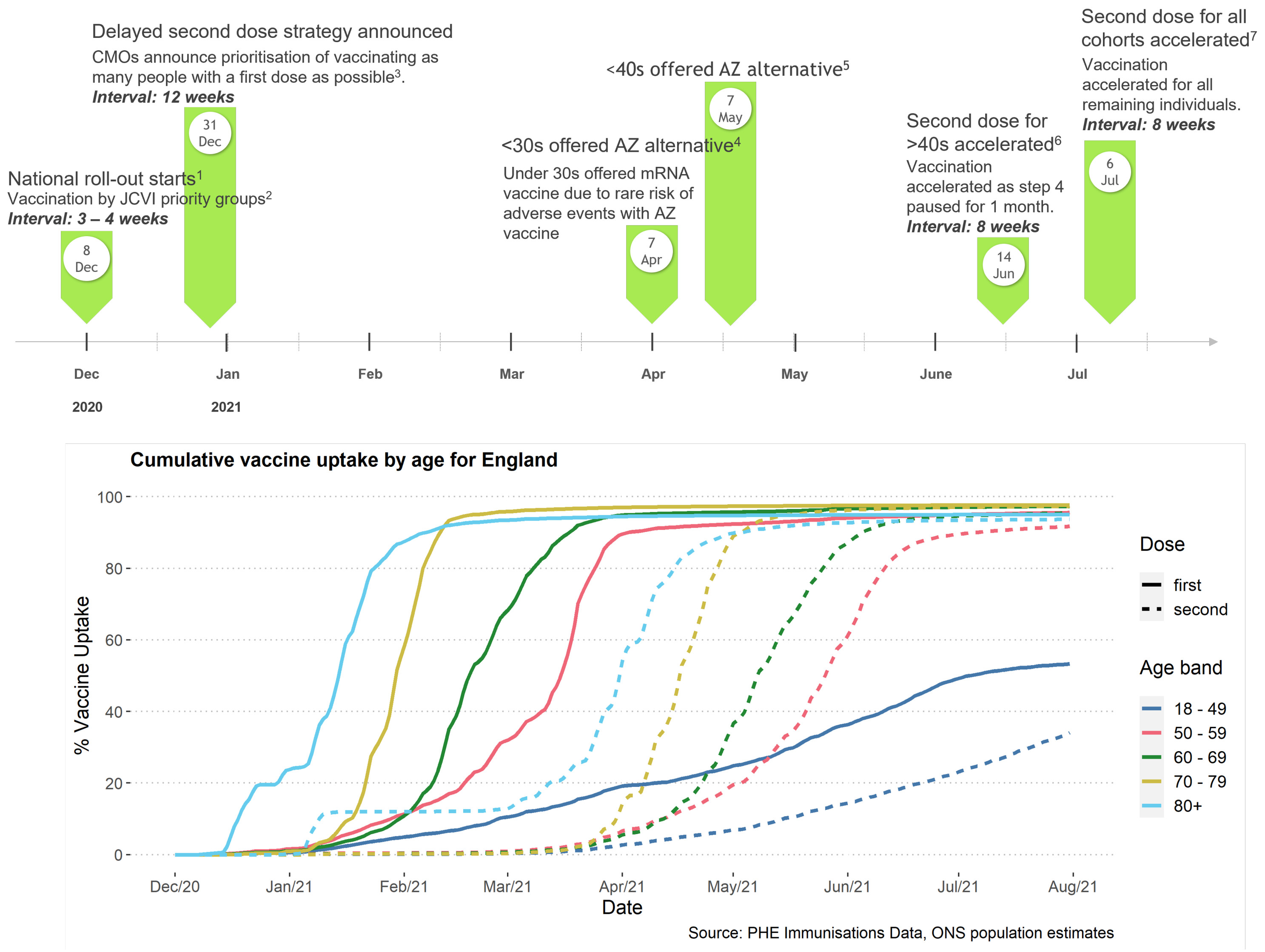

**Figure S6:** Timeline of vaccine roll-out and changing dosing schedule in England. Bottom panel shows the cumulative vaccine uptake by age for first (solid) or second (dashed lines) doses, up to August 2021 as reported in the Public Health England (now UK HSA) Immunisations Data. Shown as the proportion of the population age groups of England as reported by the Office for National Statistics.

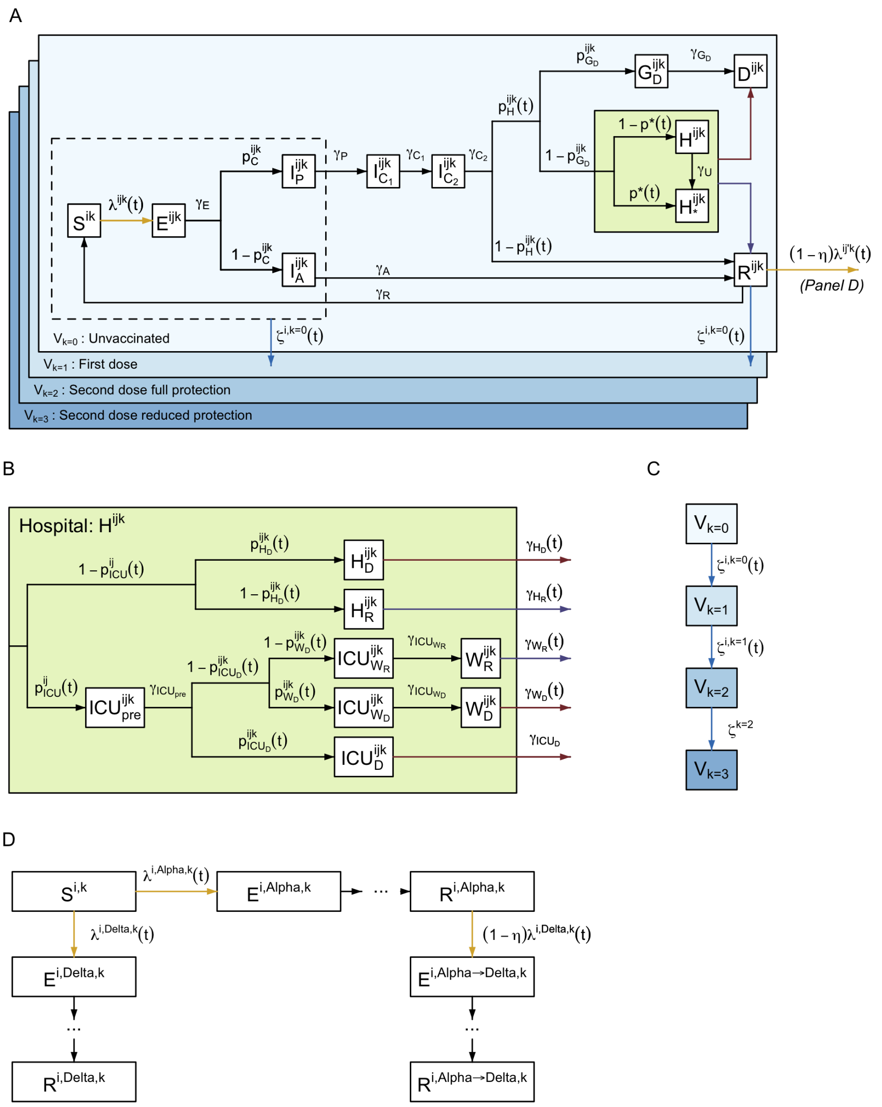

**Figure S7:** Model structure flow diagram with rates of transition between states. (A) Extended SEIR transmission model flow diagram overview. (B) Hospital flow diagram. (C) Vaccination flow diagram. (D) Multi-variant flow diagram showing possible infection with $Alpha$, $Delta$, or both in turn ($Alpha \rightarrow Delta$). All variables and parameters defined in tables S5, S6, and S7. Superscripts refer to the age or care home group $(i \in [0-4],[5-9],\ldots,[75-79],[80+],CHW,CHR)$, variant $(j = Alpha, Delta,$ or $Alpha \rightarrow Delta)$, and vaccination stratum $(k = 0,\ldots,3)$, see table S2. Identical to Figure S1, copied here for easy reference.

| Compartment | Definition |
|---|---|
| $S^{i,k}(t)$ | Susceptible. |
| $E^{i,j,k}(t)$ | Exposed (latent infection). |
| $I_A^{i,j,k}(t)$ | Asymptomatic infected. |
| $I_P^{i,j,k}(t)$ | Presymptomatic infected (infectious). |
| $I_{C_1}^{i,j,k}(t)$ | Symptomatic infected (infectious). |
| $I_{C_2}^{i,j,k}(t)$ | Symptomatic infected (not infectious). |
| $G_D^{i,j,k}(t)$ | Severely diseased, leading to death (at home). |
| $D^{i,j,k}(t)$ | Deceased (as a result of COVID-19). |
| $H_D^{i,j,k}(t)$ | Hospitalised on general ward leading to death. |
| $H_R^{i,j,k}(t)$ | Hospitalised on general ward leading to recovery. |
| $ICU_{pre}^{i,j,k}(t)$ | Awaiting admission to ICU. |
| $ICU_D^{i,j,k}(t)$ | Hospitalised in ICU, leading to death. |
| $ICU_{W_R}^{i,j,k}(t)$ | Hospitalised in ICU, leading to recovery. |
| $ICU_{W_D}^{i,j,k}(t)$ | Hospitalised in ICU, leading to death following step-down from ICU. |
| $W_R^{i,j,k}(t)$ | Step-down recovery period. |
| $W_D^{i,j,k}(t)$ | Step-down post-ICU period, leading to death. |
| $R^{i,j,k}(t)$ | Recovered. |

**Table S5:** Definitions of model compartments shown in Figure S1. $i$ defines age group ( $i \in \{[0,5),[5,10),\ldots,[75-80),[80+),CHW,CHR\}$ ), $j$ denotes the variant ($j \in \{$ *Alpha*, *Delta*, *Alpha*→*Delta* $\}$ as defined in Section 4.1), $k$ denotes vaccination strata ($k \in \{V_0,V_1,V_2,V_3\}$ as defined in Table S2). See Knock et al. [1] and Sonabend et al. [2] for further details.

| Parameter | Definition |
|---|---|
| $\lambda^{i,j,k}(t)$ | Force of infection. |
| $\gamma_x$ | Rate of progression from compartment $x$. |
| $\gamma_U$ | Rate at which unconfirmed hospital patients are confirmed as infected. |
| $p_C^{i,j,k}$ | Probability of being symptomatic if infected. |
| $p_H^{i,j,k}(t)$ | Probability of admission to hospital, conditional on symptomatic infection. |
| $p_{G_D}^{i,j,k}$ | Probability of death for severe symptomatic cases outside of hospital. |
| $p^*(t)$ | Probability of COVID-19 diagnosis confirmed prior to admission to hospital. |
| $p_{ICU}^{i,j}(t)$ | Probability of admission to ICU, conditional on hospitalisation. |
| $p_{H_D}^{i,j,k}(t)$ | Probability of death for hospitalised cases not in ICU. |
| $p_{ICU_D}^{i,j,k}(t)$ | Probability of death for cases in ICU. |
| $p_{W_D}^{i,j,k}(t)$ | Probability of death for cases after discharge from ICU. |
| $\zeta^{i,k}(t)$ | Rate of movement from vaccine strata $k$ to $k+1$. |
| $\eta$ | Probability of being protected against infection with $Delta$ for those recovered from $Alpha$ ($R^{i,Alpha,k}$), relative to those in the susceptible class ($S^{i,k}$). |

**Table S6:** Definitions of model parameters shown in Figure S1. These parameters define the routes of transmission through model compartments defined in Table S5. $i$ defines age group ( $i \in \{[0,5),[5,10),\ldots,[75-80),[80+),CHW,CHR\}$ ), $j$ denotes variant status $j$ denotes the variant ($j \in \{$ $Alpha$, $Delta$, $Alpha$→$Delta$ $\}$ as defined in Section 4.1), $k$ denotes vaccination strata ($k \in \{V_0,V_1,V_2,V_3\}$ as defined in Table S2). Refer to Knock et al. and Sonabend et al. [1, 2] for further details of parameter fitting.

#### 4.2 Parallel flows

In addition to compartments involved in the transmission dynamics and clinical progression, there are three parallel flows which we use for fitting to testing data from surveys: (i) one for PCR testing and (ii) two for serology testing (Figure S8), with separate flows used for testing with the EuroImmun and Roche N assays.

The PCR flow is used for fitting to data from the REACT-1 study. Upon infection, an individual enters the PCR flow in a pre-positivity compartment ($T_{PCR_{pre}}$) before moving into the PCR positivity compartment ($T_{PCR_{pos}}$) and then ultimately into the PCR negativity compartment ($T_{PCR_{neg}}$).

We have two serology flows to allow us to assume different distributions for the time to seroreversion when fitting to samples tested with two different assays: EuroImmun and Roche N. EuroImmun was used for testing NHS Blood and Transplant (NHSBT) samples from the first wave onwards, while Roche N only started being used in November 2020. Roche N tests only for seropositivity resulting from infection, whereas EuroImmun does not distinguish between seropositivity resulting from infection or from vaccination. Since our serology flows are only designed to capture seroconversion resulting from infection, we do not fit to samples using the EuroImmun assay from 15th January 2021 onwards as we can expect the vaccination to impact beyond this. After a seroconversion period ($T_{sero^1_{pre}}$ for EuroImmun, $T_{sero^2_{pre}}$ for Roche N), individuals can seroconvert ($T_{sero^1_{pos}}$ for EuroImmun, $T_{sero^2_{pos}}$ for Roche N) or not

($T_{sero^1_{neg}}$ for EuroImmun, $T_{sero^2_{neg}}$ for Roche N) ; if they do seroconvert, they eventually serorevert to $T_{sero^1_{neg}}$ or $T_{sero^2_{neg}}$ accordingly.

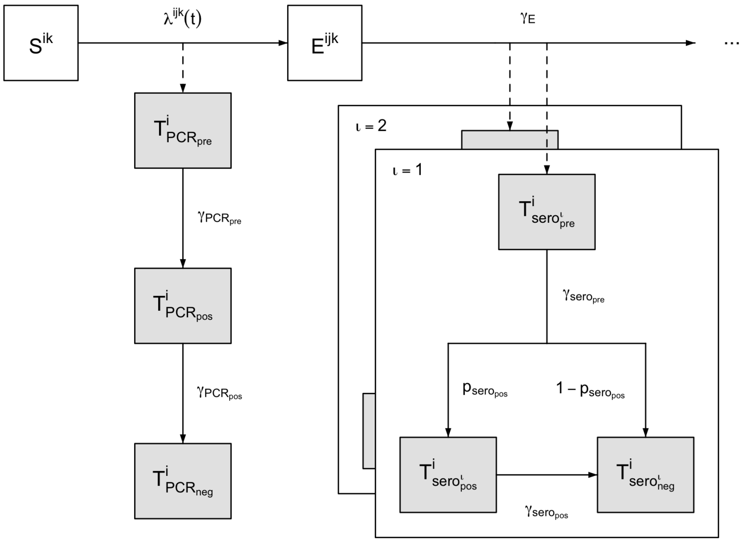

**Figure S8:** PCR positivity and seropositivity model structure flow diagram. Upon infection, an individual enters the pre-positive PCR compartment ($T_{PCR_{pre}}$) before moving into the PCR positivity compartment ($T_{PCR_{pos}}$) and then into the PCR negativity compartment ($T_{PCR_{neg}}$). After a seroreversion period ($T_{sero_{pre}}$), individuals can seroconvert ($T_{sero_{pos}}$) or not ($T_{sero_{neg}}$); if they do seroconvert, they eventually serorevert to $T_{sero_{pos}}$.

### 4.3 Equations

#### 4.3.1 Force of infection

We let $\chi^{i,j,k}$ be the susceptibility to variant $j$ of a susceptible individual in group $i$ and vaccine stratum $k$, relative to a non vaccinated individual (so that $\chi^{i,j,0} = 1$ for all $i$ and $j$), given by

$$\chi^{i,j,k} = (1 - e_{inf}^{i,j,k}), \tag{2}$$

where $e_{inf}^{i,j,k}$ is the vaccine effectiveness against infection of variant $j$ in vaccine strata $k$ (Table S4), scaled across vaccine types according to the distribution presented in Figure S3.

We let $\xi^{i,j,k}$ be the infectivity of an individual in group $i$ and vaccine stratum $k$ infected with variant $j$ relative to a non vaccinated individual infected with the Alpha variant (so that $\xi^{i,Alpha,0} = 1$). This infectivity captures both the vaccine effectiveness against infectiousness as presented in Table S3 and also the increased transmissibility of Delta compared to Alpha. As such $\xi^{i,j,k}$ is equal to

$$\xi^{i,j,k} = \begin{cases} (1 - e_{ins}^{i,j,k}), & \text{if } j = Alpha, \\ \\ (1 - e_{ins}^{i,j,k})\sigma, & \text{if } j \in \{Delta, Alpha \rightarrow Delta\}, \end{cases} \tag{3}$$

where $e_{ins}^{i,j,k}$ is the vaccine effectiveness against infectiousness of variant $j$ in vaccine strata $k$ as defined in Table S3, scaled across vaccine types according to the distribution presented in Figure S3, while $\sigma$ is the region-specific transmission advantage of Delta as described in Section 2.2.

We let $\Theta_{i,j,k}(t)$ be the number of infectious individuals with variant $j$ in group $i$ and vaccination stratum $k$, weighted by infectivity, given by:

$$\Theta_{i,j,k}(t) = \xi^{i,j,k}\left(\theta_{I_A} I_A^{i,j,k}(t) + I_P^{i,j,k}(t) + I_{C_1}^{i,j,k}(t)\right). \tag{4}$$

where $\theta_{I_A}$ is the infectivity of an asymptomatic infected individual, relative to a symptomatic individual infected with the same variant, and in the same vaccination strata.

The force of infection, $\lambda^{i,j,k}(t)$, of variant $j \in \{Alpha, Delta\}$ on a susceptible individual in group $i \in \{[0,5), \ldots, [75,80), [80+), CHW, CHR\}$ and vaccination stratum $k = 0,1,\ldots,3$ is then given by

$$\lambda^{i,Alpha,k}(t) = \chi^{i,Alpha,k} \sum_{i'} m_{i,i'}(t) \sum_{k'} \Theta_{i',Alpha,k'}(t) \tag{5}$$

$$\lambda^{i,Delta,k}(t) = \chi^{i,Delta,k} \sum_{i'} m_{i,i'}(t) \sum_{k'} \left(\Theta_{i',Delta,k'}(t) + \Theta_{i',Alpha\to Delta,k'}(t)\right) \tag{6}$$

where $m_{i,i'}(t)$ is the (symmetric) time-varying person-to-person transmission rate from group $i'$ to group $i$.

We let $\Lambda^{i,k}(t)$ be the total force of infection on a susceptible individual in group $i$ and vaccination stratum $k$, i.e.

$$\Lambda^{i,k}(t) = \lambda^{i,Alpha,k}(t) + \lambda^{i,Delta,k}(t). \tag{7}$$

Note that there is zero force of infection of $Alpha$ on all recovered individuals. The force of infection of $Delta$ on an individual recovered from $Alpha$ in group $i$ and vaccine stratum $k$ is $(1-\eta)\lambda^{i,Delta,k}(t)$, where $\eta$ is the cross immunity parameter Table S6. There is zero force of infection of $Delta$ on all individuals recovered from $Delta$ (or in the $Alpha\to Delta$ class).

Transmission between different age groups $(i,i') \in \{[0,5),\ldots,[75,80),[80+)\}^2$ was parameterised as follows:

$$m_{i,i'}(t) = \beta(t) c_{i,i'}, \tag{8}$$

where $c_{i,i'}$ is the (symmetric) person-to-person contact rate between age group $i$ and $i'$, derived from pre-pandemic data from the POLYMOD survey [3] for the United Kingdom. For each region, the `socialmixr` package [43] was used to derive the contact matrix between different age groups $(i,i') \in \{[0,5),\ldots,[75,80),[80+)\}^2$, which was then scaled by the regional population demography to yield the required person-to-person daily contact rate matrix, $c_{i,i'}$.

$\beta(t)$ is the time-varying transmission rate which encompasses both changes over time in transmission efficiency (e.g. due to temperature) and temporal changes in the overall level of contacts in the population (due to changes in policy and behaviours).

We assumed $\beta(t)$ to be piecewise linear:

$$\beta(t) = \begin{cases} \beta_i, & \text{if } t \le t_i,\ i = 1 \\ \dfrac{t_i - t}{t_i - t_{i-1}}\beta_{i-1} + \dfrac{t - t_{i-1}}{t_i - t_{i-1}}\beta_i, & \text{if } t_{i-1} < t \le t_i,\ i \in \{2, \dots, 27\} \\ \beta_i & \text{if } t > t_i,\ i = 27 \end{cases} \tag{9}$$

with 27 change points $t_i$ corresponding to major announcements and changes in COVID-19 related policy (Table S8).

We defined parameters representing transmission rates within care homes (between and among workers and residents), which were assumed to be constant over time. Parameter $m_{CHW}$ represents the person-to-person transmission rate among care home workers and between care home workers and residents; $m_{CHR}$ represents the person-to-person transmission rate among care home residents; these are defined as:

$$m_{CHW,CHW}(t) = m_{CHW,CHR}(t) = m_{CHW} \tag{10}$$

$$m_{CHR,CHR}(t) = m_{CHR} \tag{11}$$

Transmission between the general population and care home workers was assumed to be similar to that within the general population, accounting for the average age of care home workers, with, for $i \in \{[0,5), \dots, [75,80), [80+)\}$,

$$m_{i,CHW}(t) = \beta(t) c_{i,CHW}, \tag{12}$$

where $c_{i,CHW}$ is the mean of $c_{i,[25,30)}, c_{i,[30,35)}, \dots, c_{i,[60,65)}$ (i.e. of the age groups that the care home workers are drawn from).

Transmission between the general population and care home residents was assumed to be similar to that between the general population and the 80+ age group, adjusted by a reduction factor $\varepsilon$ (which was inferred, see Table S8), such that, for $i \in \{[0,5), \dots, [75,80), [80+)\}$,

$$m_{i,CHR} = \varepsilon \beta(t) c_{i,80+}. \tag{13}$$

These represent contact between visitors from the general community and care home residents. This might involve a slightly different age profile than the age profile of the contact made by people in the 80+ age group.

##### 4.3.2 Pathway probabilities and rates

The movement between model compartments is primarily dictated by the parameters $p_x^{i,j,k}$, defining the probability of progressing to compartment $x$ (Table S6), as well as rate parameters $\gamma$. These parameters vary between age groups ($i$), variant of infection ($j$), and vaccine strata ($k$). This section outlines how these differences are formally defined and calculated.

The probability that an individual will have a symptomatic infection given that they have been infected is given by

$$p_C^{i,j,k} = p_C^i \left(1 - e_{sympt|inf}^{i,j,k}\right), \tag{14}$$

where $p_C^i$ is given in [1].

The probability that an individual has severe disease requiring hospitalisation given that they have symptoms is given by

$$p_H^{i,j,k}(t) = \min\left\{h_H(t)\,\psi_H^i\left(1-e_{SD|sympt}^{i,j,k}\right)\pi_H^j,\ 1\right\}, \tag{15}$$

where $\psi_H^i$ is given in [1, 2] (with a slight amendment that now $\psi_H^{CHR}=1$) and $h_H(t)$ has a piecewise linear form with changepoints defined as follows:

$$h_H = (t)\begin{cases} p_{H,1}^{max} & \text{on (and before) 01/20/20,} \\ p_{H,2}^{max} & \text{on 15/12/20,} \\ p_{H,3}^{max} & \text{on (and after) 02/02/21,} \end{cases} \tag{16}$$

and $\pi_H^j$ is a multiplier accounting for the increased severity of hospitalisation for $Delta$ (see Section 2.4):

$$\pi_H^j = \begin{cases} 1 & \text{if } j = Alpha, \\ \pi_H^{Delta} & \text{if } j = Delta \\ \pi_H^{Delta}\left(1-e_{SD|sympt}^{PF2}\right) & \text{if } j = Alpha+Delta. \end{cases} \tag{17}$$

where $e^{PF2}$ denotes full VE for Pfizer after 2 doses.

Within the main manuscript, Figure 3C presents $(p_C^{i,j,k}(t)\cdot p_H^{i,j,k}(t))$ on the first date of each month, averaged over age, region, and vaccine strata $(i,j,k)$.

The probability that an individual dies in the community (or a care home if $i = CHR$) given they have severe disease is

$$p_{G_D}^{i,j,k}(t) = \begin{cases} \min\left\{p_{G_D}e_{death|SD}^{i,j,k}\pi_D^j,\ 1\right\} & \text{if } i \neq CHR, \\ \min\left\{p_{G_D}^{CHR}e_{death|SD}^{i,j,k}\pi_D^j,\ 1\right\} & \text{if } i = CHR, \end{cases} \tag{18}$$

and $\pi_D^j$ is a multiplier accounting for the increased severity of $Delta$ (see Section 2.4):

$$\pi_D^j = \begin{cases} 1 & \text{if } j = Alpha, \\ \pi_D^{Delta} & \text{if } j = Delta,\ Alpha+Delta. \end{cases} \tag{19}$$

The probability that an individual will be admitted to ICU given that they have been hospitalised is

$$p_{ICU}^{i,j}(t) = \min\left\{h_{ICU}(t)\,\psi_{ICU}^i\pi_{ICU}^j,\ 1\right\}, \tag{20}$$

where $\psi_{ICU}^i$ is given in [1, 2] and $h_{ICU}(t)$ has a piecewise linear form with changepoints defined as follows:

$$h_{ICU} = (t)\begin{cases} p_{ICU,1}^{max} & \text{on (and before) 01/04/20,} \\ p_{ICU,2}^{max} & \text{on 01/06/20,} \end{cases} \tag{21}$$

and $\pi_{ICU}^j$ is a multiplier accounting for the increased severity of $Delta$ (see Section 2.4):

$$\pi_{ICU}^j = \begin{cases} 1 & \text{if } j = Alpha, \\ \pi_{ICU}^{Delta} & \text{if } j = Delta,\ Alpha+Delta. \end{cases} \tag{22}$$

The probability that an individual will die in general beds given that they are not admitted to ICU is

$$p_{H_D}^{i,j,k}(t) = \min\left\{p_{H_D}^{max} h_D(t)\psi_{H_D}^{i} e_{death|SD}^{i,j,k} \pi_D^j, 1\right\}, \tag{23}$$

where $\psi_{H_D}^i$ is given in [1], $h_D(t)$ has a piecewise linear form with changepoints defined as follows:

$$h_D = (t)\begin{cases} 1 & \text{on (and before) 01/04/20,} \\ \mu_{D,1}^{max} & \text{on 01/06/20,} \\ \mu_{D,1}^{max} & \text{on 01/10/20,} \\ \mu_{D,2}^{max} & \text{on 15/12/20,} \\ \mu_{D,2}^{max} & \text{on 15/01/21,} \\ \mu_{D,1}^{max} & \text{on (and after) 01/02/21.} \end{cases} \tag{24}$$

The probability that an individual dies in ICU given that they are not admitted to ICU is

$$p_{ICU_D}^{i,j,k}(t) = \min\left\{p_{ICU_D}^{max} h_D(t)\psi_{ICU_D}^{i} e_{death|SD}^{i,j,k} \pi_D^j,\ 1\right\} \tag{25}$$

where $\psi_{ICU_D}^i$ is given in [1].

The probability that an individual who has been in ICU dies in stepdown beds given that they have not died in ICU is

$$p_{W_D}^{i,j,k}(t) = \min\left\{p_{W_D}^{max} h_D(t)\psi_{W_D}^{i} e_{death|SD}^{i,j,k} \pi_D^j,\ 1\right\}, \tag{26}$$

where $\psi_{W_D}^i$ is given in [1].

Finally, the probability of individuals having had a COVID-19 diagnosis confirmed prior to admission to hospital, $p^*(t)$ has a piecewise linear form with changepoints defined as follows:

$$p^*(t) = \begin{cases} 0.1 & \text{on (and before) 15/03/20,} \\ 0.42 & \text{on 01/07/20,} \\ 0.2 & \text{on 09/09/20,} \\ 0.45 & \text{on (and after) 27/06/21.} \end{cases} \tag{27}$$

These were informed by data on COVID-19 admissions and inpatient diagnoses for England from NHS England [44].

In addition, the duration rates for some hospital compartments are time-varying to account for changes in length of stay over time. We let

$$\gamma_{H_R}(t) = h_\gamma(t)\gamma_{H_R} \tag{28}$$
$$\gamma_{H_D}(t) = h_\gamma(t)\gamma_{H_D} \tag{29}$$
$$\gamma_{W_R}(t) = h_\gamma(t)\gamma_{W_R} \tag{30}$$
$$\gamma_{W_D}(t) = h_\gamma(t)\gamma_{W_D} \tag{31}$$
$$\tag{32}$$

where $h_\gamma(t)$ has a piecewise linear form with changepoints given by

$$h_\gamma(t) = \begin{cases} 1 & \text{on (and before) 01/12/20,} \\ 1/\mu_{\gamma_H,1} & \text{on 01/01/21,} \\ 1/\mu_{\gamma_H,2} & \text{on 01/03/21,} \\ 1/\mu_{\gamma_H,3} & \text{on 01/06/21,} \\ 1/\mu_{\gamma_H,4} & \text{on (and after) 01/09/21.} \end{cases} \tag{33}$$

#### 4.3.3 Compartmental model equations

To clearly illustrate the model dynamics, we describe a deterministic version of the model in differential equations (35)-(75), followed by the stochastic implementation used in the analysis. Full definitions of compartments and model parameters are set out in Tables S5 and S6. Unless otherwise specified, $\sum_j$ refers to the sum across all combinations of variants (i.e. $j \in \{Alpha, Delta, Alpha \rightarrow Delta\}$), and $\sum_k$ refers to the sum across all vaccination strata ($k \in \{0,1,2,3\}$).

In the following model equations we use $\mathbb{1}_A$ as an indicator function, such that

$$\mathbb{1}_A(j) := \begin{cases} 1 & \text{if } j \in A, \\ 0 & \text{if } j \notin A. \end{cases} \tag{34}$$

Further note that we split some compartments in two distinct compartments. For example, the exposed class, $E^{i,j,k}$, is modelled via two separate compartments, $E^{i,j,k,1}$ and $E^{i,j,k,2}$ (equations (36) and (37)). This is to be able to capture a non exponentially distributed duration of stay in certain compartments; the split allows us to model the duration of stay as an Erlang distribution instead (sum of independent exponential distributions) [45].

$$\frac{dS^{i,k}(t)}{dt} = \zeta^{i,k-1}(t)S^{i,k-1}(t) - \left(\zeta^{i,k}(t) + \Lambda^{i,k}(t)\right)S^{i,k}(t) - \sum_j \delta^{i,j,k}(t) + \gamma_R \sum_j R^{i,j,k}(t) \tag{35}$$

$$\begin{aligned} \frac{dE^{i,j,k,1}(t)}{dt} &= \mathbb{1}_{\{Alpha,Delta\}}(j)\lambda^{i,j,k}(t)S^{i,k}(t) + \mathbb{1}_{\{Alpha \rightarrow Delta\}}(j)\,(1-\eta)\,\lambda^{i,Delta,k}(t)R^{i,Alpha,k}(t) \\ &\quad + \zeta^{i,k-1}(t)E^{i,j,k-1,1}(t) + \delta^{i,j,k}(t) - \left(\gamma_E + \zeta^{i,k}(t)\right)E^{i,j,k,1}(t) \end{aligned} \tag{36}$$

$$\frac{dE^{i,j,k,2}(t)}{dt} = \gamma_E E^{i,j,k,1}(t) + \zeta^{i,k-1}(t)E^{i,j,k-1,2}(t) - \left(\gamma_E + \zeta^{i,k}(t)\right)E^{i,j,k,2}(t) \tag{37}$$

$$\frac{dI_A^{i,j,k}(t)}{dt} = \left(1 - p_C^{i,j,k}\right)\gamma_E E^{i,j,k,2}(t) + \zeta^{i,k-1}(t)I_A^{i,j,k-1}(t) - \left(\gamma_A + \zeta^{i,k}(t)\right)I_A^{i,j,k}(t) \tag{38}$$

$$\frac{dI_P^{i,j,k}(t)}{dt} = p_C^{i,j,k}\gamma_E E^{i,j,k,2}(t) + \zeta^{i,k-1}(t)I_P^{i,j,k-1}(t) - \left(\gamma_P + \zeta^{i,k}(t)\right)I_P^{i,j,k}(t) \tag{39}$$

$$\frac{dI_{C_1}^{i,j,k}(t)}{dt} = \gamma_P I_P^{i,j,k}(t) - \gamma_{C_1} I_{C_1}^{i,j,k}(t) \tag{40}$$

$$\frac{dI_{C_2}^{i,j,k}(t)}{dt} = \gamma_{C_1} I_{C_1}^{i,j,k}(t) - \gamma_{C_2} I_{C_2}^{i,j,k}(t) \tag{41}$$

$$\frac{dG_D^{i,j,k,1}(t)}{dt} = p_H^{i,j,k}(t)p_{G_D}^{i,j,k}\gamma_{C_2} I_{C_2}^{i,j,k}(t) - \gamma_{G_D} G_D^{i,j,k,1}(t) \tag{42}$$

$$\frac{dG_D^{i,j,k,2}(t)}{dt} = \gamma_{G_D} G_D^{i,j,k,1}(t) - \gamma_{G_D} G_D^{i,j,k,2}(t) \tag{43}$$

$$\begin{aligned} \frac{dICU_{pre}^{i,j,k}(t)}{dt} &= p_H^{i,j,k}(t)\left(1 - p_{G_D}^{i,j,k}\right)(1 - p^*(t))\,p_{ICU}^{i,j}(t)\gamma_{C_2} I_{C_2}^{i,j,k}(t) \\ &\quad - \left(\gamma_{ICU_{pre}} + \gamma_U\right) ICU_{pre}^{i,j,k}(t) \end{aligned} \tag{44}$$

$$
\begin{aligned}
\frac{dICU_{pre^*}^{i,j,k}(t)}{dt} &= p_H^{i,j,k}(t)\left(1-p_{G_D}^{i,j,k}\right)p^*(t)p_{ICU}^{i,j}(t)\gamma_{C_2}I_{C_2}^{i,j,k}(t)-\gamma_{ICU_{pre}}ICU_{pre^*}^{i,j,k}(t) \\
&\quad+\gamma_U ICU_{pre}^{i,j,k}(t)
\end{aligned} \tag{45}
$$

$$
\begin{aligned}
\frac{dICU_{W_R}^{i,j,k}(t)}{dt} &= \left(1-p_{ICU_D}^{i,j,k}(t)\right)\left(1-p_{W_D}^{i,j,k}(t)\right)\gamma_{ICU_{pre}}ICU_{pre}^{i,j,k}(t) \\
&\quad-\left(\gamma_{ICU_{W_R}}+\gamma_U\right)ICU_{W_R}^{i,j,k}(t)
\end{aligned} \tag{46}
$$

$$
\begin{aligned}
\frac{dICU_{W_{R^*}}^{i,j,k}(t)}{dt} &= \left(1-p_{ICU_D}^{i,j,k}(t)\right)\left(1-p_{W_D}^{i,j,k}(t)\right)\gamma_{ICU_{pre}}ICU_{pre^*}^{i,j,k}(t)-\gamma_{ICU_{W_R}}ICU_{W_{R^*}}^{i,j,k}(t) \\
&\quad+\gamma_U ICU_{W_R}^{i,j,k}(t)
\end{aligned} \tag{47}
$$

$$
\frac{dICU_{W_D}^{i,j,k}(t)}{dt} = \left(1-p_{ICU_D}^{i,j,k}(t)\right)p_{W_D}^{i,j,k}(t)\gamma_{ICU_{pre}}ICU_{pre}^{i,j,k}(t)-\left(\gamma_{ICU_{W_D}}+\gamma_U\right)ICU_{W_D}^{i,j,k}(t) \tag{48}
$$

$$
\begin{aligned}
\frac{dICU_{W_{D^*}}^{i,j,k}(t)}{dt} &= \left(1-p_{ICU_D}^{i,j,k}(t)\right)p_{W_D}^{i,j,k}(t)\gamma_{ICU_{pre}}ICU_{pre^*}^{i,j,k}(t)-\gamma_{ICU_{W_D}}ICU_{W_{D^*}}^{i,j,k}(t) \\
&\quad+\gamma_U ICU_{W_D}^{i,j,k}(t)
\end{aligned} \tag{49}
$$

$$
\frac{dICU_D^{i,j,k,1}(t)}{dt} = p_{ICU_D}^{i,j,k}(t)\gamma_{ICU_{pre}}ICU_{pre}^{i,j,k}(t)-\left(\gamma_{ICU_D}+\gamma_U\right)ICU_D^{i,j,k,1}(t) \tag{50}
$$

$$
\frac{dICU_D^{i,j,k,2}(t)}{dt} = \gamma_{ICU_D}ICU_D^{i,j,k,1}(t)-\left(\gamma_{ICU_D}+\gamma_U\right)ICU_D^{i,j,k,2}(t) \tag{51}
$$

$$
\frac{dICU_{D^*}^{i,j,k,1}(t)}{dt} = p_{ICU_D}^{i,j,k}(t)\gamma_{ICU_{pre}}ICU_{pre^*}^{i,j,k}(t)-\gamma_{ICU_D}ICU_{D^*}^{i,j,k,1}(t)+\gamma_U ICU_D^{i,j,k,1}(t) \tag{52}
$$

$$
\frac{dICU_{D^*}^{i,j,k,2}(t)}{dt} = \gamma_{ICU_D}ICU_{D^*}^{i,j,k,1}(t)-\gamma_{ICU_D}ICU_{D^*}^{i,j,k,2}(t)+\gamma_U ICU_D^{i,j,k,2}(t) \tag{53}
$$

$$
\frac{dW_R^{i,j,k,1}(t)}{dt} = \gamma_{ICU_{W_R}}ICU_{W_R}^{i,j,k}(t)-\left(\gamma_{W_R}(t)+\gamma_U\right)W_R^{i,j,k,1}(t) \tag{54}
$$

$$
\frac{dW_R^{i,j,k,2}(t)}{dt} = \gamma_{W_R}(t)W_R^{i,j,k,1}(t)-\left(\gamma_{W_R}(t)+\gamma_U\right)W_R^{i,j,k,2}(t) \tag{55}
$$

$$
\frac{dW_{R^*}^{i,j,k,1}(t)}{dt} = \gamma_{ICU_{W_R}}ICU_{W_{R^*}}^{i,j,k}(t)-\gamma_{W_R}(t)W_{R^*}^{i,j,k,1}(t)+\gamma_U W_R^{i,j,k,1}(t) \tag{56}
$$

$$
\frac{dW_{R^*}^{i,j,k,2}(t)}{dt} = \gamma_{W_R}(t)W_{R^*}^{i,j,k,1}(t)-\gamma_{W_R}(t)W_{R^*}^{i,j,k,2}(t)+\gamma_U W_R^{i,j,k,2}(t) \tag{57}
$$

$$
\frac{dW_D^{i,j,k}(t)}{dt} = \gamma_{ICU_{W_D}}ICU_{W_D}^{i,j,k}(t)-\left(\gamma_{W_D}(t)+\gamma_U\right)W_D^{i,j,k}(t) \tag{58}
$$

$$
\frac{dW_{D^*}^{i,j,k}(t)}{dt} = \gamma_{ICU_{W_D}}ICU_{W_{D^*}}^{i,j,k}(t)-\gamma_{W_D}(t)W_{D^*}^{i,j,k}(t)+\gamma_U W_D^{i,j,k}(t) \tag{59}
$$

$$
\begin{aligned}
\frac{dH_R^{i,j,k}(t)}{dt} &= p_H^{i,j,k}(t)\left(1-p_{G_D}^{i,j,k}\right)\left(1-p^*(t)\right)\left(1-p_{ICU}^{i,j}(t)\right)\left(1-p_{H_D}^{i,j,k}(t)\right)\gamma_{C_2}I_{C_2}^{i,j,k}(t) \\
&\quad-\left(\gamma_{H_R}(t)+\gamma_U\right)H_R^{i,j,k}(t)
\end{aligned} \tag{60}
$$

$$\frac{dH_{R^*}^{i,j,k}(t)}{dt} = p_H^{i,j,k}(t)\left(1-p_{G_D}^{i,j,k}\right)p^*(t)\left(1-p_{ICU}^{i,j}(t)\right)\left(1-p_{H_D}^{i,j,k}(t)\right)\gamma_{C_2}I_{C_2}^{i,j,k}(t) + \gamma_U H_R^{i,j,k}(t) - \gamma_{H_R}(t)H_{R^*}^{i,j,k}(t) \tag{61}$$

$$\frac{dH_D^{i,j,k,1}(t)}{dt} = p_H^{i,j,k}(t)\left(1-p_{G_D}^{i,j,k}\right)(1-p^*(t))\left(1-p_{ICU}^{i,j}(t)\right)p_{H_D}^{i,j,k}(t)\gamma_{C_2}I_{C_2}^{i,j,k}(t) - (\gamma_{H_D}(t)+\gamma_U)H_D^{i,j,k,1}(t) \tag{62}$$

$$\frac{dH_D^{i,j,k,2}(t)}{dt} = \gamma_{H_D}(t)H_D^{i,j,k,1}(t) - (\gamma_{H_D}(t)+\gamma_U)H_D^{i,j,k,2}(t) \tag{63}$$

$$\frac{dH_{D^*}^{i,j,k,1}(t)}{dt} = p_H^{i,j,k}(t)\left(1-p_{G_D}^{i,j,k}\right)p^*(t)\left(1-p_{ICU}^{i,j}(t)\right)\ p_{H_D}^{i,j,k}(t)\gamma_{C_2}I_{C_2}^{i,j,k}(t) + \gamma_U H_D^{i,j,k,1}(t) - \gamma_{H_D}(t)H_{D^*}^{i,j,k,1}(t) \tag{64}$$

$$\frac{dH_{D^*}^{i,j,k,2}(t)}{dt} = \gamma_{H_D}(t)H_{D^*}^{i,j,k,1}(t) - \gamma_{H_D}(t)H_{D^*}^{i,j,k,2}(t) + \gamma_U H_D^{i,j,k,2}(t) \tag{65}$$

$$\frac{dR^{i,j,k}(t)}{dt} = \gamma_A I_A^{i,j,k}(t) + \left(1-p_H^{i,j,k}(t)\right)\gamma_{C_2}I_{C_2}^{i,j,k}(t) + \gamma_{H_R}(t)\left(H_R^{i,j,k}(t)+H_{R^*}^{i,j,k}(t)\right) + \gamma_{W_R}(t)\left(W_R^{i,j,k,2}(t)+W_{R^*}^{i,j,k,2}(t)\right) + \zeta^{i,k-1}(t)R^{i,j,k-1}(t) - \left(\gamma_R+\zeta^{i,k}(t)\right)R^{i,j,k}(t) - \mathbb{1}_{\{Alpha\}}(j)(1-\eta)\lambda^{i,Delta,k}(t)R^{i,j,k}(t) \tag{66}$$

$$\frac{dT_{sero_{pre}^1}^i(t)}{dt} = -\gamma_{sero_{pre}}T_{sero_{pre}^1}^i(t) + \sum_j\sum_k \gamma_E E^{i,j,k,2}(t) \tag{67}$$

$$\frac{dT_{sero_{pos}^1}^i(t)}{dt} = p_{sero_{pos}}\gamma_{sero_{pre}}T_{sero_{pre}^1}^i(t) - \gamma_{sero_{pos}^1}T_{sero_{pos}^1}^i(t) \tag{68}$$

$$\frac{dT_{sero_{neg}^1}^i(t)}{dt} = \left(1-p_{sero_{pos}}\right)\gamma_{sero_{pre}}T_{sero_{pre}^1}^i(t) + \gamma_{sero_{pos}^1}T_{sero_{pos}^1}^i(t) \tag{69}$$

$$\frac{dT_{sero_{pre}^2}^i(t)}{dt} = -\gamma_{sero_{pre}}T_{sero_{pre}^2}^i(t) + \sum_j\sum_k \gamma_E E^{i,j,k,2}(t) \tag{70}$$

$$\frac{dT_{sero_{pos}^2}^i(t)}{dt} = p_{sero_{pos}}\gamma_{sero_{pre}}T_{sero_{pre}^2}^i(t) - \gamma_{sero_{pos}^2}T_{sero_{pos}^2}^i(t) \tag{71}$$

$$\frac{dT_{sero_{neg}^2}^i(t)}{dt} = \left(1-p_{sero_{pos}}\right)\gamma_{sero_{pre}}T_{sero_{pre}^2}^i(t) + \gamma_{sero_{pos}^2}T_{sero_{pos}^2}^i(t) \tag{72}$$

$$\frac{dT_{PCR_{pre}}^i(t)}{dt} = -\gamma_{PCR_{pre}}T_{PCR_{pre}}^i(t) + \sum_k\left(\lambda^{i,Alpha,k}(t)+\lambda^{i,Delta,k}(t)\right)S^{i,k}(t) \tag{73}$$

$$\frac{dT_{PCR_{pos}}^i(t)}{dt} = \gamma_{PCR_{pre}}T_{PCR_{pre}}^i(t) - \gamma_{PCR_{pos}}T_{PCR_{pos}}^i(t) \tag{74}$$

$$\frac{dT_{PCR_{neg}}^i(t)}{dt} = \gamma_{PCR_{pos}}T_{PCR_{pos}}^i(t). \tag{75}$$

We used the tau-leap method [46] to create a stochastic, time-discretised version of the model described in equations (78) - (211), taking four update steps per day ($dt = 0.25$ days).

For each time step, the model iterated through the procedure described below. In the following, we introduce a small abuse of notation: for transitions involving multiple onward compartments (e.g. transition from compartment $E$ to compartments $I_A$ or $I_P$ or to the next vaccination strata within $E$), for conciseness, we write

$$\left(d_{E,I_A}^{i,j,k},\ d_{E,I_P}^{i,j,k},\ d_{E,v}^{i,j,k}\right) \sim \text{Multinom}\left(E^{i,j,k,2}(t),\ q_{E,I_A}^{i,j,k},\ q_{E,I_P}^{i,j,k},\ q_{E,v}^{i,j,k}\right) \tag{76}$$

instead of

$$\left(d_{E,I_A}^{i,j,k},\ d_{E,I_P}^{i,j,k},\ d_{E,v}^{i,j,k},\ d_{nomove}^{i,j,k}\right) \sim \text{Multinom}\left(E^{i,j,k,2}(t),\ q_{E,I_A}^{i,j,k},\ q_{E,I_P}^{i,j,k},\ q_{E,v}^{i,j,k},\ 1-\sum_{x\in I_A,I_P,v} q_{E,x}^{i,j,k}\right) \tag{77}$$

where $d_{nomove}^{i,j,k}$ is a dummy variable counting the number of individuals remaining in compartment $E^{i,j,k,2}$. We also omit the time dependency i.e. we use $d_{E,I_A}^{i,j,k}$ or $q_{E,I_A}^{i,j,k}$ instead of $d_{E,I_A}^{i,j,k}(t)$ or $q_{E,I_A}^{i,j,k}(t)$.

Using this convention, transition variables are drawn from the following distributions, with probabilities defined below:

$$q_{S,E}^{i,Alpha,k} = \left(1-e^{-\Lambda^{i,k}(t)dt}\right)\frac{\lambda^{i,Alpha,k}(t)}{\Lambda^{i,k}(t)} \tag{78}$$

$$q_{S,E}^{i,Delta,k} = \left(1-e^{-\Lambda^{i,k}(t)dt}\right)\frac{\lambda^{i,Delta,k}(t)}{\Lambda^{i,k}(t)} \tag{79}$$

$$q_{S,v}^{i,k} = 1-e^{-\zeta^{i,k}(t)dt} \tag{80}$$

$$\left(d_{S,E}^{i,Alpha,k},\ d_{S,E}^{i,Delta,k}\right) \sim \text{Multinom}\left(S^{i,k}(t),\ q_{S,E}^{i,Alpha,k},\ q_{S,E}^{i,Delta,k}\right) \tag{81}$$

$$d_{seed}^{i,j,k} \sim \min\left(\text{Poisson}\left(\hat{\delta}^{i,j,k}(t)dt\right),\ S^{i,k}(t)-d_{S,E}^{i,Alpha,k}-d_{S,E}^{i,Delta,k}\right) \tag{82}$$

$$d_{S,v}^{i,k} \sim \text{Binom}\left(S^{i,k}(t)-d_{seed}^{i,j,k}-d_{S,E}^{i,Alpha,k}-d_{S,E}^{i,Delta,k},\ q_{S,v}^{i,k}\right) \tag{83}$$

$$\left(q_{E,E}^{i,j,k},\ q_{E,v}^{i,j,k,1}\right) = \left(1-e^{-\gamma_E dt},\ e^{-\gamma_E dt}\left(1-e^{-\zeta^{i,k}(t)dt}\right)\right) \tag{84}$$

$$\left(d_{E,E}^{i,j,k},\ d_{E,v}^{i,j,k,1}\right) \sim \text{Multinom}\left(E^{i,j,k,1}(t),\ q_{E,E}^{i,j,k},\ q_{E,v}^{i,j,k,1}\right) \tag{85}$$

$$q_{E,I_A}^{i,j,k} = \left(1-p_C^{i,j,k}\right)\left(1-e^{-\gamma_E dt}\right) \tag{86}$$

$$q_{E,I_P}^{i,j,k} = p_C^{i,j,k}\left(1-e^{-\gamma_E dt}\right) \tag{87}$$

$$q_{E,v}^{i,j,k,2} = e^{-\gamma_E dt}\left(1-e^{-\zeta^{i,k}(t)dt}\right) \tag{88}$$

$$\left(d_{E,I_A}^{i},\ d_{E,I_P}^{i},\ d_{E,v}^{i,j,k,2}\right) \sim \text{Multinom}\left(E^{i,j,k,2}(t),\ q_{E,I_A}^{i,j,k},\ q_{E,I_P}^{i,j,k},\ q_{E,v}^{i,j,k,2}\right) \tag{89}$$

$$\left(q_{I_A,R}^{i,j,k},\ q_{I_A,v}^{i,j,k}\right) = \left(1-e^{-\gamma_A dt},\ e^{-\gamma_A dt}\left(1-e^{-\zeta^{i,k}(t)dt}\right)\right) \tag{90}$$

$$\left(d_{I_A,R}^{i,j,k},\ d_{I_A,v}^{i,j,k}\right) \sim \text{Multinom}\left(I_A^{i}(t),\ q_{I_A,R}^{i,j,k},\ q_{I_A,v}^{i,j,k}\right) \tag{91}$$

$$\left(q_{I_P,I_{C_1}}^{i,j,k},\ q_{I_P,v}^{i,j,k}\right) = \left(1-e^{-\gamma_P dt},\ e^{-\gamma_P dt}\left(1-e^{-\zeta^{i,k}(t)dt}\right)\right) \tag{92}$$

$$\left(d_{I_P,I_{C_1}}^{i,j,k},\ d_{I_P,v}^{i,j,k}\right) \sim \text{Multinom}\left(I_P^{i}(t),\ q_{I_P,I_{C_1}}^{i,j,k},\ q_{I_P,v}^{i,j,k}\right) \tag{93}$$

$$d^{i,j,k}_{I_{C_1},I_{C_2}} \sim \text{Binom}\left(I^{i,j,k}_{C_1}(t),\ 1-e^{-\gamma_{C_1}dt}\right) \tag{94}$$

$$q^{i,j,k}_{I_{C_2},G_D} = p^{i,j,k}_{H}(t)p^{i,j,k}_{G_D}\left(1-e^{-\gamma_{C_2}dt}\right) \tag{95}$$

$$q^{i,j,k}_{I_{C_2},R} = \left(1-p^{i,j,k}_{H}(t)\right)\left(1-e^{-\gamma_{C_2}dt}\right) \tag{96}$$

$$q^{i,j,k}_{I_{C_2},ICU_{pre}} = p^{i,j,k}_{H}(t)\left(1-p^{i,j,k}_{G_D}\right)(1-p^*(t))\,p^{i,j}_{ICU}(t)\left(1-e^{-\gamma_{C_2}dt}\right) \tag{97}$$

$$q^{i,j,k}_{I_{C_2},ICU_{pre^*}} = p^{i,j,k}_{H}(t)\left(1-p^{i,j,k}_{G_D}\right)p^*(t)p^{i,j}_{ICU}(t)\left(1-e^{-\gamma_{C_2}dt}\right) \tag{98}$$

$$q^{i,j,k}_{I_{C_2},H_R} = p^{i,j,k}_{H}(t)\left(1-p^{i,j,k}_{G_D}\right)(1-p^*(t))\left(1-p^{i,j}_{ICU}(t)\right)\left(1-p^{i,j,k}_{H_D}(t)\right)\left(1-e^{-\gamma_{C_2}dt}\right) \tag{99}$$

$$q^{i,j,k}_{I_{C_2},H_{R^*}} = p^{i,j,k}_{H}(t)\left(1-p^{i,j,k}_{G_D}\right)\ p^*(t)\left(1-p^{i,j}_{ICU}(t)\right)\left(1-p^{i,j,k}_{H_D}(t)\right)\left(1-e^{-\gamma_{C_2}dt}\right) \tag{100}$$

$$q^{i,j,k}_{I_{C_2},H_D} = p^{i,j,k}_{H}(t)\left(1-p^{i,j,k}_{G_D}\right)(1-p^*(t))\left(1-p^{i,j}_{ICU}(t)\right)p^{i,j,k}_{H_D}(t)\left(1-e^{-\gamma_{C_2}dt}\right) \tag{101}$$

$$q^{i,j,k}_{I_{C_2},H_{D^*}} = p^{i,j,k}_{H}(t)\left(1-p^{i,j,k}_{G_D}\right)p^*(t)\left(1-p^{i,j}_{ICU}(t)\right)p^{i,j,k}_{H_D}(t)\left(1-e^{-\gamma_{C_2}dt}\right) \tag{102}$$

$$\left(d^{i,j,k}_{I_{C_2},G_D},\ldots,d^{i,j,k}_{I_{C_2},H_{D^*}}\right) \sim \text{Multinom}\left(I^{i,j,k}_{C_2}(t),\ q^{i,j,k}_{I_{C_2},G_D},\ldots,q^{i,j,k}_{I_{C_2},H_{D^*}}\right) \tag{103}$$

$$d^{i,j,k}_{G_D,G_D} \sim \text{Binom}\left(G^{i,j,k,1}_{D}(t),\ 1-e^{-\gamma_{G_D}dt}\right) \tag{104}$$

$$d^{i,j,k}_{G_D,D} \sim \text{Binom}\left(G^{i,j,k,2}_{D}(t),\ 1-e^{-\gamma_{G_D}dt}\right) \tag{105}$$

$$q^{i,j,k}_{ICU_{pre},ICU_{W_R}} = \left(1-p^{i,j,k}_{ICU_D}(t)\right)\left(1-p^{i,j,k}_{W_D}(t)\right)\left(1-e^{-\gamma_{ICU_{pre}}dt}\right)e^{-\gamma_U dt} \tag{106}$$

$$q^{i,j,k}_{ICU_{pre},ICU_{W_{R^*}}} = \left(1-p^{i,j,k}_{ICU_D}(t)\right)\left(1-p^{i,j,k}_{W_D}(t)\right)\left(1-e^{-\gamma_{ICU_{pre}}dt}\right)\left(1-e^{-\gamma_U dt}\right) \tag{107}$$

$$q^{i,j,k}_{ICU_{pre},ICU_{W_D}} = \left(1-p^{i,j,k}_{ICU_D}(t)\right)p^{i,j,k}_{W_D}(t)\left(1-e^{-\gamma_{ICU_{pre}}dt}\right)e^{-\gamma_U dt} \tag{108}$$

$$q^{i,j,k}_{ICU_{pre},ICU_{W_{D^*}}} = \left(1-p^{i,j,k}_{ICU_D}(t)\right)p^{i,j,k}_{W_D}(t)\left(1-e^{-\gamma_{ICU_{pre}}dt}\right)\left(1-e^{-\gamma_U dt}\right) \tag{109}$$

$$q^{i,j,k}_{ICU_{pre},ICU_D} = p^{i,j,k}_{ICU_D}(t)\left(1-e^{-\gamma_{ICU_{pre}}dt}\right)e^{-\gamma_U dt} \tag{110}$$

$$q^{i,j,k}_{ICU_{pre},ICU_{D^*}} = p^{i,j,k}_{ICU_D}(t)\left(1-e^{-\gamma_{ICU_{pre}}dt}\right)\left(1-e^{-\gamma_U dt}\right) \tag{111}$$

$$q^{i,j,k}_{ICU_{pre},ICU_{pre^*}} = e^{-\gamma_{ICU_{pre}}dt}\left(1-e^{-\gamma_U dt}\right) \tag{112}$$

$$\begin{aligned}\left(d^{i,j,k}_{ICU_{pre},ICU_{W_R}},\ \ldots, d^{i,j,k}_{ICU_{pre},ICU_{pre^*}}\right)&\\ \sim \text{Multinom}&\left(ICU^{i,j,k}_{pre}(t),\ q^{i,j,k}_{ICU_{pre},ICU_{W_R}},\ldots,q^{i,j,k}_{ICU_{pre},ICU_{pre^*}}\right)\end{aligned} \tag{113}$$

$$q^{i,j,k}_{ICU_{pre^*},ICU_{W_{R^*}}} = \left(1-p^{i,j,k}_{ICU_D}(t)\right)\left(1-p^{i,j,k}_{W_D}(t)\right)\left(1-e^{-\gamma_{ICU_{pre}}dt}\right) \tag{114}$$

$$q^{i,j,k}_{ICU_{pre^*},ICU_{W_{D^*}}} = \left(1-p^{i,j,k}_{ICU_D}(t)\right)p^{i,j,k}_{W_D}(t)\left(1-e^{-\gamma_{ICU_{pre}}dt}\right) \tag{115}$$

$$q^{i,j,k}_{ICU_{pre^*},ICU_{D^*}} = p^{i,j,k}_{ICU_D}(t)\left(1-e^{-\gamma_{ICU_{pre}}dt}\right) \tag{116}$$

$$\begin{aligned}\left(d^{i,j,k}_{ICU_{pre^*},ICU_{W_{R^*}}},\ \ldots,\ d^{i,j,k}_{ICU_{pre^*},ICU_{D^*}}\right)&\\ \sim \text{Multinom}&\left(ICU^{i,j,k}_{pre^*}(t),\ q^{i,j,k}_{ICU_{pre^*},ICU_{W_{R^*}}},\ldots,q^{i,j,k}_{ICU_{pre^*},ICU_{D^*}}\right)\end{aligned} \tag{117}$$

$$q^{i,j,k}_{H_D,H_D} = \left(1-e^{-\gamma_{H_D}(t)dt}\right) e^{-\gamma_U dt} \tag{118}$$

$$q^{i,j,k,1,1}_{H_D,H_{D^*}} = e^{-\gamma_{H_D}(t)dt}\left(1-e^{-\gamma_U dt}\right) \tag{119}$$

$$q^{i,j,k,1,2}_{H_D,H_{D^*}} = \left(1-e^{-\gamma_{H_D}(t)dt}\right)\left(1-e^{-\gamma_U dt}\right) \tag{120}$$

$$\begin{aligned}&\left(d^{i,j,k}_{H_D,H_D},\ d^{i,j,k,1,1}_{H_D,H_{D^*}},\ d^{i,j,k,1,2}_{H_D,H_{D^*}}\right)\\ &\qquad\sim \text{Multinom}\left(H^{i,j,k,1}_D(t),\ q^{i,j,k}_{H_D,H_D},\ q^{i,j,k,1,1}_{H_D,H_{D^*}},\ q^{i,j,k,1,2}_{H_D,H_{D^*}}\right)\end{aligned} \tag{121}$$

$$d^{i,j,k}_{H_{D^*},H_{D^*}} \sim \text{Binom}\left(H^{i,j,k,1}_{D^*}(t),\ 1-e^{-\gamma_{H_D}(t)dt}\right) \tag{122}$$

$$\left(d^{i,j,k}_{H_D,D},\ d^{i,j,k,2,2}_{H_D,H_{D^*}}\right) \sim \text{Multinom}\left(H^{i,j,k,2}_D(t),\ 1-e^{-\gamma_{H_D}(t)dt},\ e^{-\gamma_{H_D}(t)dt}\left(1-e^{-\gamma_U dt}\right)\right) \tag{123}$$

$$d^{i,j,k}_{H_{D^*},D} \sim \text{Binom}\left(H^{i,j,k,2}_{D^*}(t),\ 1-e^{-\gamma_{H_D}(t)dt}\right) \tag{124}$$

$$\left(d^{i,j,k}_{H_R,R},\ d^{i,j,k}_{H_R,H_{R^*}}\right) \sim \text{Multinom}\left(H^{i,j,k}_R(t),\ 1-e^{-\gamma_{H_R}(t)dt},\ e^{-\gamma_{H_R}(t)dt}\left(1-e^{-\gamma_U dt}\right)\right) \tag{125}$$

$$d^{i,j,k}_{H_{R^*},R} \sim \text{Binom}\left(H^{i,j,k}_{R^*}(t),\ 1-e^{-\gamma_{H_R}(t)dt}\right) \tag{126}$$

$$q^{i,j,k}_{ICU_{W_R},W_R} = \left(1-e^{-\gamma_{ICU_{W_R}}dt}\right) e^{-\gamma_U dt} \tag{127}$$

$$q^{i,j,k}_{ICU_{W_R},ICU_{W_{R^*}}} = e^{-\gamma_{ICU_{W_R}}dt}\left(1-e^{-\gamma_U dt}\right) \tag{128}$$

$$q^{i,j,k}_{ICU_{W_R},W_{R^*}} = \left(1-e^{-\gamma_{ICU_{W_R}}dt}\right)\left(1-e^{-\gamma_U dt}\right) \tag{129}$$

$$\begin{aligned}&\left(d^{i,j,k}_{ICU_{W_R},W_R},\ \ldots,d^{i,j,k}_{ICU_{W_R},W_{R^*}}\right)\\ &\qquad\sim \text{Multinom}\left(ICU^{i,j,k}_{W_R}(t),\ q^{i,j,k}_{ICU_{W_R},W_R},\ldots,q^{i,j,k}_{ICU_{W_R},W_{R^*}}\right)\end{aligned} \tag{130}$$

$$d^{i,j,k}_{ICU_{W_{R^*}},W_{R^*}} \sim \text{Binom}\left(ICU^{i,j,k}_{W_{R^*}}(t),\ 1-e^{-\gamma_{ICU_{W_R}}dt}\right) \tag{131}$$

$$q^{i,j,k}_{ICU_{W_D},W_D} = \left(1-e^{-\gamma_{ICU_{W_D}}dt}\right) e^{-\gamma_U dt} \tag{132}$$

$$q^{i,j,k}_{ICU_{W_D},ICU_{W_{D^*}}} = e^{-\gamma_{ICU_{W_D}}dt}\left(1-e^{-\gamma_U dt}\right) \tag{133}$$

$$q^{i,j,k}_{ICU_{W_D},W_{D^*}} = \left(1-e^{-\gamma_{ICU_{W_D}}dt}\right)\left(1-e^{-\gamma_U dt}\right) \tag{134}$$

$$\begin{aligned}&\left(d^{i,j,k}_{ICU_{W_D},W_D},\ \ldots,d^{i,j,k}_{ICU_{W_D},W_{D^*}}\right)\\ &\qquad\sim \text{Multinom}\left(ICU^{i,j,k}_{W_D}(t),\ q^{i,j,k}_{ICU_{W_D},W_D},\ldots,q^{i,j,k}_{ICU_{W_D},W_{D^*}}\right)\end{aligned} \tag{135}$$

$$d^{i,j,k}_{ICU_{W_{D^*}},W_{D^*}} \sim \text{Binom}\left(ICU^{i,j,k}_{W_{D^*}}(t),\ 1-e^{-\gamma_{ICU_{W_D}}dt}\right) \tag{136}$$

$$q^{i,j,k}_{ICU_D,ICU_D} = \left(1-e^{-\gamma_{ICU_D}dt}\right) e^{-\gamma_U dt} \tag{137}$$

$$q^{i,j,k,1,1}_{ICU_D,ICU_{D^*}} = e^{-\gamma_{ICU_D}dt}\left(1-e^{-\gamma_U dt}\right) \tag{138}$$

$$q^{i,j,k,1,2}_{ICU_D,ICU_{D^*}} = \left(1-e^{-\gamma_{ICU_D}dt}\right)\left(1-e^{-\gamma_U dt}\right) \tag{139}$$

$$\begin{aligned}\left(d^{i,j,k}_{ICU_D,ICU_D},\ d^{i,j,k,1,1}_{ICU_D,ICU_{D^*}},\ d^{i,j,k,1,2}_{ICU_D,ICU_{D^*}}\right)\\ \sim \text{Multinom}\left(ICU^{i,j,k,1}_D(t),\ q^{i,j,k}_{ICU_D,ICU_D},\ q^{i,j,k,1,1}_{ICU_D,ICU_{D^*}},\ q^{i,j,k,1,2}_{ICU_D,ICU_{D^*}}\right)\end{aligned} \tag{140}$$

$$d^{i,j,k}_{ICU_{D^*},ICU_{D^*}} \sim \text{Binom}\left(ICU^{i,j,k,1}_{D^*}(t),\ 1-e^{-\gamma_{ICU_D}dt}\right) \tag{141}$$

$$\left(q^{i,j,k}_{ICU_D,D},\ q^{i,j,k,2,2}_{ICU_D,ICU_{D^*}}\right) = \left(1-e^{-\gamma_{ICU_D}dt},\ e^{-\gamma_{ICU_D}dt}\left(1-e^{-\gamma_U dt}\right)\right) \tag{142}$$

$$\left(d^{i,j,k}_{ICU_D,D},\ d^{i,j,k,2,2}_{ICU_D,ICU_{D^*}}\right) \sim \text{Multinom}\left(ICU^{i,j,k,2}_D(t),\ q^{i,j,k}_{ICU_D,D},\ q^{i,j,k,2,2}_{ICU_D,ICU_{D^*}}\right) \tag{143}$$

$$d^{i,j,k}_{ICU_{D^*},D} \sim \text{Binom}\left(ICU^{i,j,k,2}_{D^*}(t),\ 1-e^{-\gamma_{ICU_D}dt}\right) \tag{144}$$

$$q^{i,j,k}_{W_R,W_R} = \left(1-e^{-\gamma_{W_R}(t)dt}\right)e^{-\gamma_U dt} \tag{145}$$

$$q^{i,j,k,1,1}_{W_R,W_{R^*}} = e^{-\gamma_{W_R}(t)dt}\left(1-e^{-\gamma_U dt}\right) \tag{146}$$

$$q^{i,j,k,1,2}_{W_R,W_{R^*}} = \left(1-e^{-\gamma_{W_R}(t)dt}\right)\left(1-e^{-\gamma_U dt}\right) \tag{147}$$

$$\begin{aligned}\left(d^{i,j,k}_{W_R,W_R},\ d^{i,j,k,1,1}_{W_R,W_{R^*}},\ d^{i,j,k,1,2}_{W_R,W_{R^*}}\right)\\ \sim \text{Multinom}\left(W^{i,j,k,1}_R(t),\ q^{i,j,k}_{W_R,W_R},\ q^{i,j,k,1,1}_{W_R,W_{R^*}},\ q^{i,j,k,1,2}_{W_R,W_{R^*}}\right)\end{aligned} \tag{148}$$

$$d^{i,j,k}_{W_{R^*},W_{R^*}} \sim \text{Binom}\left(W^{i,j,k,1}_{R^*}(t),\ 1-e^{-\gamma_{W_R}(t)dt}\right) \tag{149}$$

$$\left(q^{i,j,k}_{W_R,R},\ q^{i,j,k,2,2}_{W_R,W_{R^*}}\right) = \left(1-e^{-\gamma_{W_R}(t)dt},\ e^{-\gamma_{W_R}(t)dt}\left(1-e^{-\gamma_U\ dt}\right)\right) \tag{150}$$

$$\left(d^{i,j,k}_{W_R,R},\ d^{i,j,k,2,2}_{W_R,W_{R^*}}\right) \sim \text{Multinom}\left(W^{i,j,k,2}_R(t),\ q^{i,j,k}_{W_R,R},\ q^{i,j,k,2,2}_{W_R,W_{R^*}}\right) \tag{151}$$

$$d^{i,j,k}_{W_{R^*},R} \sim \text{Binom}\left(W^{i,j,k,2}_{R^*}(t),\ 1-e^{-\gamma_{W_R}(t)dt}\right) \tag{152}$$

$$\left(q^{i,j,k}_{W_D,D},\ q^{i,j,k}_{W_D,W_{D^*}}\right) = \left(1-e^{-\gamma_{W_D}(t)dt},\ e^{-\gamma_{W_D}(t)dt}\left(1-e^{-\gamma_U dt}\right)\right) \tag{153}$$

$$\left(d^{i,j,k}_{W_D,D},\ d^{i,j,k}_{W_D,W_{D^*}}\right) \sim \text{Multinom}\left(W^{i,j,k}_D(t),\ q^{i,j,k}_{W_D,D},\ q^{i,j,k}_{W_D,W_{D^*}}\right) \tag{154}$$

$$d^{i,j,k}_{W_{D^*},D} \sim \text{Binom}\left(W^{i,j,k}_{D^*}(t),\ 1-e^{-\gamma_{W_D}(t)dt}\right) \tag{155}$$

$$\gamma^{i,j,k}_{R,E} = \mathbb{1}_{\{Alpha\}}(j)(1-\eta)\lambda^{i,Delta,k}(t) \tag{156}$$

$$q^{i,j,k}_{R,S} = \left(1-e^{-\left(\gamma_R+\gamma^{i,j,k}_{R,E}\right)dt}\right)\frac{\gamma_R}{\gamma_R+\gamma^{i,j,k}_{R,E}} \tag{157}$$

$$q^{i,j,k}_{R,E} = \left(1-e^{-\left(\gamma_R+\gamma^{i,j,k}_{R,E}\right)dt}\right)\frac{\gamma^{i,j,k}_{R,E}}{\gamma_R+\gamma^{i,j,k}_{R,E}} \tag{158}$$

$$q^{i,j,k}_{R,v} = e^{-\left(\gamma_R+\gamma^{i,j,k}_{R,E}\right)dt}\left(1-e^{-\zeta^{i,k}(t)dt}\right) \tag{159}$$

$$\left(d^{i,j,k}_{R,S},\ d^{i,j,k}_{R,E},\ d^{i,j,k}_{R,v}\right) \sim \text{Multinom}\left(R^{i,j,k}(t),\ q^{i,j,k}_{R,S},\ q^{i,j,k}_{R,E},\ q^{i,j,k}_{R,v}\right) \tag{160}$$

$$q^{i}_{T_{sero^1_{pre}},T_{sero^1_{pos}}} = p_{sero_{pos}}\left(1-e^{-\gamma_{sero_{pre}}dt}\right) \tag{161}$$

$$q^{i}_{T_{sero^1_{pre}},T_{sero^1_{neg}}} = \left(1-p_{sero_{pos}}\right)\left(1-e^{-\gamma_{sero_{pre}}dt}\right) \tag{162}$$

$$\left(d^i_{T_{sero^1_{pre}},T_{sero^1_{pos}}},\ d^i_{T_{sero^1_{pre}},T_{sero^1_{neg}}}\right) \sim \text{Multinom}\left(T^i_{sero^1_{pre}}(t),\ q^i_{T_{sero^1_{pre}},T_{sero^1_{pos}}},\ q^i_{T_{sero^1_{pre}},T_{sero^1_{neg}}}\right) \tag{163}$$

$$d^i_{T_{sero^1_{pos}},T_{sero^1_{neg}}} \sim \text{Binom}\left(T^i_{sero^1_{pos}}(t),\ 1-e^{-\gamma_{sero^1_{pos}}dt}\right) \tag{164}$$

$$q^i_{T_{sero^2_{pre}},T_{sero^2_{pos}}} = p_{sero_{pos}}\left(1-e^{-\gamma_{sero_{pre}}dt}\right) \tag{165}$$

$$q^i_{T_{sero^2_{pre}},T_{sero^2_{neg}}} = \left(1-p_{sero_{pos}}\right)\left(1-e^{-\gamma_{sero_{pre}}dt}\right) \tag{166}$$

$$\left(d^i_{T_{sero^2_{pre}},T_{sero^2_{pos}}},\ d^i_{T_{sero^2_{pre}},T_{sero^2_{neg}}}\right) \sim \text{Multinom}\left(T^i_{sero^2_{pre}}(t),\ q^i_{T_{sero^2_{pre}},T_{sero^2_{pos}}},\ q^i_{T_{sero^2_{pre}},T_{sero^2_{neg}}}\right) \tag{167}$$

$$d^i_{T_{sero^2_{pos}},T_{sero^2_{neg}}} \sim \text{Binom}\left(T^i_{sero^2_{pos}}(t),\ 1-e^{-\gamma_{sero^2_{pos}}dt}\right) \tag{168}$$

$$d^i_{T_{PCR_{pre}},T_{PCR_{pos}}} \sim \text{Binom}\left(T^i_{PCR_{pre}}(t),\ 1-e^{-\gamma_{PCR_{pre}}dt}\right) \tag{169}$$

$$d^i_{T_{PCR_{pos}},T_{PCR_{neg}}} \sim \text{Binom}\left(T^i_{PCR_{pos}}(t),\ 1-e^{-\gamma_{PCR_{pos}}dt}\right) \tag{170}$$

Model compartments were then updated as follows (Note that $d^{i,Alpha \to Delta,k}_{S,E} = 0$):

$$S^{i,k}(t+dt) := S^{i,k}(t) - d^{i,Alpha,k}_{S,E} - d^{i,Delta,k}_{S,E} + d^{i,j,k}_{R,S} + d^{i,k-1}_{S,v} - d^{i,k}_{S,v} - \sum_j d^{i,j,k}_{seed} \tag{171}$$

$$E^{i,j,k,1}(t+dt) := E^{i,j,k,1}(t) + d^{i,j,k}_{S,E} + \mathbb{1}_{\{Alpha \to Delta\}}(j) d^{i,Alpha,k}_{R,E} - d^{i,j,k}_{E,E} + d^{i,j,k-1,1}_{E,v} - d^{i,j,k,1}_{E,v} + d^{i,j,k}_{seed} \tag{172}$$

$$E^{i,j,k,2}(t+dt) := E^{i,j,k,2}(t) + d^{i,j,k}_{E,E} - d^{i,j,k}_{E,I_A} - d^{i,j,k}_{E,I_P} + d^{i,j,k-1,2}_{E,v} - d^{i,j,k,2}_{E,v} \tag{173}$$

$$I^{i,j,k}_A(t+dt) := I^{i,j,k}_A(t) + d^{i,j,k}_{E,I_A} - d^{i,j,k}_{I_A,R} + d^{i,j,k-1}_{I_A,v} - d^{i,j,k}_{I_A,v} \tag{174}$$

$$I^{i,j,k}_P(t+dt) := I^{i,j,k}_P(t) + d^{i,j,k}_{E,I_P} - d^{i,j,k}_{I_P,I_{C_1}} + d^{i,j,k-1}_{I_P,v} - d^{i,j,k}_{I_P,v} \tag{175}$$

$$I^{i,j,k}_{C_1}(t+dt) := I^{i,j,k}_{C_1}(t) + d^{i,j,k}_{I_P,I_{C_1}} - d^{i,j,k}_{I_{C_1},I_{C_2}} \tag{176}$$

$$\begin{aligned} I^{i,j,k}_{C_2}(t+dt) := {} & I^{i,j,k}_{C_2}(t) + d^{i,j,k}_{I_{C_1},I_{C_2}} - d^{i,j,k}_{I_{C_2},G_D} - d^{i,j,k}_{I_{C_2},R} - d^{i,j,k}_{I_{C_2},ICU_{pre}} \\ & - d^{i,j,k}_{I_{C_2},ICU_{pre^*}} - d^{i,j,k}_{I_{C_2},H_R} - d^{i,j,k}_{I_{C_2},H_{R^*}} - d^{i,j,k}_{I_{C_2},H_D} - d^{i,j,k}_{I_{C_2},H_{D^*}} \end{aligned} \tag{177}$$

$$G^{i,j,k,1}_D(t+dt) := G^{i,j,k,1}_D(t) + d^{i,j,k}_{I_{C_2},G_D} - d^{i,j,k}_{G_D,G_D} \tag{178}$$

$$G^{i,j,k,2}_D(t+dt) := G^{i,j,k,2}_D(t) + d^{i,j,k}_{G_D,G_D} - d^{i,j,k}_{G_D,D} \tag{179}$$

$$\begin{aligned} ICU^{i,j,k}_{pre}(t+dt) := {} & ICU^{i,j,k}_{pre}(t) + d^{i,j,k}_{I_{C_2},ICU_{pre}} - d^{i,j,k}_{ICU_{pre},ICU_{W_R}} - d^{i,j,k}_{ICU_{pre},ICU_{W_D}} \\ & - d^{i,j,k}_{ICU_{pre},ICU_D} - d^{i,j,k}_{ICU_{pre},ICU_{pre^*}} - d^{i,j,k}_{ICU_{pre},ICU_{W_{R^*}}} - d^{i,j,k}_{ICU_{pre},ICU_{W_{D^*}}} \\ & - d^{i,j,k}_{ICU_{pre},ICU_{D^*}} \end{aligned} \tag{180}$$

$$ICU^{i,j,k}_{pre^*}(t+dt) := ICU^{i,j,k}_{pre^*}(t) + d^{i,j,k}_{IC_2,ICU_{pre^*}} - d^{i,j,k}_{ICU_{pre},ICU_{W_{D^*}}} - d^{i,j,k}_{ICU_{pre^*},ICU_{W_{R^*}}} - d^{i,j,k}_{ICU_{pre^*},ICU_{D^*}} \tag{181}$$

$$ICU^{i,j,k}_{W_R}(t+dt) := ICU^{i,j,k}_{W_R}(t) + d^{i,j,k}_{ICU_{pre},ICU_{W_R}} - d^{i,j,k}_{ICU_{W_R},W_R} - d^{i,j,k}_{ICU_{W_R},ICU_{W_{R^*}}} - d^{i,j,k}_{ICU_{W_R},W_{R^*}} \tag{182}$$

$$ICU^{i,j,k}_{W_{R^*}}(t+dt) := ICU^{i,j,k}_{W_{R^*}}(t) + d^{i,j,k}_{ICU_{pre^*},ICU_{W_{R^*}}} + d^{i,j,k}_{ICU_{W_R},ICU_{W_{R^*}}} + d^{i,j,k}_{ICU_{pre},ICU_{W_{R^*}}} - d^{i,j,k}_{ICU_{W_{R^*}},W_{R^*}} \tag{183}$$

$$ICU^{i,j,k}_{W_D}(t+dt) := ICU^{i,j,k}_{W_D}(t) + d^{i,j,k}_{ICU_{pre},ICU_{W_D}} - d^{i,j,k}_{ICU_{W_D},W_D} - d^{i,j,k}_{ICU_{W_D},ICU_{W_{D^*}}} - d^{i,j,k}_{ICU_{W_D},W_{D^*}} \tag{184}$$

$$ICU^{i,j,k}_{W_{D^*}}(t+dt) := ICU^{i,j,k}_{W_{D^*}}(t) + d^{i,j,k}_{ICU_{pre^*},ICU_{W_{D^*}}} + d^{i,j,k}_{ICU_{W_D},ICU_{W_{D^*}}} + d^{i,j,k}_{ICU_{pre},ICU_{W_{D^*}}} - d^{i,j,k}_{ICU_{W_{D^*}},W_{D^*}} \tag{185}$$

$$ICU^{i,j,k,1}_{D}(t+dt) := ICU^{i,j,k,1}_{D}(t) + d^{i,j,k}_{ICU_{pre},ICU_D} - d^{i,j,k}_{ICU_D,ICU_D} - d^{i,j,k,1,1}_{ICU_D,ICU_{D^*}} - d^{i,j,k,1,2}_{ICU_D,ICU_{D^*}} \tag{186}$$

$$ICU^{i,j,k,2}_{D}(t+dt) := ICU^{i,j,k,2}_{D}(t) + d^{i,j,k}_{ICU_D,ICU_D} - d^{i,j,k}_{ICU_D,D} - d^{i,j,k,2,2}_{ICU_D,ICU_{D^*}} \tag{187}$$

$$ICU^{i,j,k,1}_{D^*}(t+dt) := ICU^{i,j,k,1}_{D^*}(t) + d^{i,j,k}_{ICU_{pre^*},ICU_{D^*}} + d^{i,j,k,1,1}_{ICU_D,ICU_{D^*}} + d^{i,j,k}_{ICU_{pre},ICU_{D^*}} - d^{i,j,k}_{ICU_{D^*},ICU_{D^*}} \tag{188}$$

$$ICU^{i,j,k,2}_{D^*}(t+dt) := ICU^{i,j,k,2}_{D^*}(t) + d^{i,j,k}_{ICU_{D^*},ICU_{D^*}} + d^{i,j,k,1,2}_{ICU_D,ICU_{D^*}} + d^{i,j,k,2,2}_{ICU_D,ICU_{D^*}} - d^{i,j,k}_{ICU_{D^*},D} \tag{189}$$

$$W^{i,j,k,1}_R(t+dt) := W^{i,j,k,1}_R(t) + d^{i,j,k}_{ICU_{W_R},W_R} - d^{i,j,k}_{W_R,W_R} - d^{i,j,k,1,1}_{W_R,W_{R^*}} - d^{i,j,k,1,2}_{W_R,W_{R^*}} \tag{190}$$

$$W^{i,j,k,2}_R(t+dt) := W^{i,j,k,2}_R(t) + d^{i,j,k}_{W_R,W_R} - d^{i,j,k}_{W_R,R} - d^{i,j,k,2,2}_{W_R,W_{R^*}} \tag{191}$$

$$W^{i,j,k,1}_{R^*}(t+dt) := W^{i,j,k,1}_{R^*}(t) + d^{i,j,k}_{ICU_{W_{R^*}},W_{R^*}} + d^{i,j,k,1,1}_{W_R,W_{R^*}} + d^{i,j,k}_{ICU_{W_R},W_{R^*}} - d^{i,j,k}_{W_{R^*},W_{R^*}} \tag{192}$$

$$W^{i,j,k,2}_{R^*}(t+dt) := W^{i,j,k,2}_{R^*}(t) + d^{i,j,k}_{W_{R^*},W_{R^*}} + d^{i,j,k,2,2}_{W_R,W_{R^*}} + d^{i,j,k,1,2}_{W_R,W_{R^*}} - d^{i,j,k}_{W_{R^*},R} \tag{193}$$

$$W^{i,j,k}_D(t+dt) := W^{i,j,k}_D(t) + d^{i,j,k}_{ICU_{W_D},W_D} - d^{i,j,k}_{W_D,D} - d^{i,j,k}_{W_D,W_{D^*}} \tag{194}$$

$$W^{i,j,k}_{D^*}(t+dt) := W^{i,j,k}_{D^*}(t) + d^{i,j,k}_{ICU_{W_{D^*}},W_{D^*}} + d^{i,j,k}_{W_D,W_{D^*}} + d^{i,j,k}_{ICU_{W_D},W_{D^*}} - d^{i,j,k}_{W_{D^*},D} \tag{195}$$

$$H^{i,j,k,1}_D(t+dt) := H^{i,j,k,1}_D(t) + d^{i,j,k}_{IC_2,H_D} - d^{i,j,k}_{H_D,H_D} - d^{i,j,k,1,1}_{H_D,H_{D^*}} - d^{i,j,k,1,2}_{H_D,H_{D^*}} \tag{196}$$

$$H^{i,j,k,2}_D(t+dt) := H^{i,j,k,2}_D(t) + d^{i,j,k}_{H_D,H_D} - d^{i,j,k}_{H_D,D} - d^{i,j,k,2,2}_{H_D,H_{D^*}} \tag{197}$$

$$H^{i,j,k,1}_{D^*}(t+dt) := H^{i,j,k,1}_{D^*}(t) + d^{i,j,k}_{IC_2,H_{D^*}} + d^{i,j,k,1,1}_{H_D,H_{D^*}} - d^{i,j,k}_{H_{D^*},H_{D^*}} \tag{198}$$

$$H^{i,j,k,2}_{D^*}(t+dt) := H^{i,j,k,2}_{D^*}(t) + d^{i,j,k}_{H_{D^*},H_{D^*}} + d^{i,j,k,2,2}_{H_D,H_{D^*}} + d^{i,j,k,1,2}_{H_D,H_{D^*}} - d^{i,j,k}_{H_{D^*},D} \tag{199}$$

$$H^{i,j,k}_R(t+dt) := H^{i,j,k}_R(t) + d^{i,j,k}_{IC_2,H_R} - d^{i,j,k}_{H_R,R} - d^{i,j,k}_{H_R,H_{R^*}} \tag{200}$$

$$H^{i}_{R^*}(t+dt) := H^{i,j,k}_{R^*}(t) + d^{i,j,k}_{IC_2,H_{R^*}} + d^{i,j,k}_{H_R,H_{R^*}} - d^{i,j,k}_{H_{R^*},R} \tag{201}$$

$$R^{i,j,k}(t+dt) := R^{i,j,k}(t) + d^{i,j,k}_{I_A,R} + d^{i,j,k}_{I_{C_2},R} + d^{i,j,k}_{H_R,R} + d^{i,j,k}_{H_{R^*},R} + d^{i,j,k}_{W_R,R} + d^{i,j,k}_{W_{R^*},R} \\ - d^{i,j,k}_{R,S} - \mathbb{1}_{\{Alpha\}} d^{i,Alpha,k}_{R,E} + d^{i,j,k-1}_{R,v} - d^{i,j,k}_{R,v} \tag{202}$$

$$T^i_{sero^1_{pre}}(t+dt) := T^i_{sero^1_{pre}}(t) - d^i_{T_{sero^1_{pre}},T_{sero^1_{pos}}} - d^i_{T_{sero^1_{pre}},T_{sero^1_{neg}}} + \sum_j \sum_k d^{i,j,k}_{E,I_A} + d^{i,j,k}_{E,I_P} \tag{203}$$

$$T^i_{sero^1_{pos}}(t+dt) := T^i_{sero^1_{pos}}(t) + d^i_{T_{sero^1_{pre}},T_{sero^1_{pos}}} - d^i_{T_{sero^1_{pos}},T_{sero^1_{neg}}} \tag{204}$$

$$T^i_{sero^1_{neg}}(t+dt) := T^i_{sero^1_{neg}}(t) + d^i_{T_{sero^1_{pre}},T_{sero^1_{neg}}} + d^i_{T_{sero^1_{pos}},T_{sero^1_{neg}}} \tag{205}$$

$$T^i_{sero^2_{pre}}(t+dt) := T^i_{sero^2_{pre}}(t) - d^i_{T_{sero^2_{pre}},T_{sero^2_{pos}}} - d^i_{T_{sero^2_{pre}},T_{sero^2_{neg}}} + \sum_j \sum_k d^{i,j,k}_{E,I_A} + d^{i,j,k}_{E,I_P} \tag{206}$$

$$T^i_{sero^2_{pos}}(t+dt) := T^i_{sero^2_{pos}}(t) + d^i_{T_{sero^2_{pre}},T_{sero^2_{pos}}} - d^i_{T_{sero^2_{pos}},T_{sero^2_{neg}}} \tag{207}$$

$$T^i_{sero^2_{neg}}(t+dt) := T^i_{sero^2_{neg}}(t) + d^i_{T_{sero^2_{pre}},T_{sero^2_{neg}}} + d^i_{T_{sero^2_{pos}},T_{sero^2_{neg}}} \tag{208}$$

$$T^i_{PCR_{pre}}(t+dt) := T^i_{PCR_{pre}}(t) - d^i_{T_{PCR_{pre}},T_{PCR_{pos}}} + \sum_j \sum_k d^{i,j,k}_{S,E} \tag{209}$$

$$T^i_{PCR_{pos}}(t+dt) := T^i_{PCR_{pos}}(t) + d^i_{T_{PCR_{pre}},T_{PCR_{pos}}} - d^i_{T_{PCR_{pos}},T_{PCR_{neg}}} \tag{210}$$

$$T^i_{PCR_{neg}}(t+dt) := T^i_{PCR_{neg}}(t) + d^i_{T_{PCR_{pos}},T_{PCR_{neg}}}. \tag{211}$$

$$\tag{212}$$

Note that the fitted seeding dates of the epidemic ($t_{Alpha}$) and Delta ($t_{Delta}$) have continuous support, and the seeding process (see Section 2.1) is handled within the discretisation to four update steps per day such that:

$$\hat{\delta}^{i,j,k}(t) = \begin{cases} \phi_j f_j(t) & \text{if } i = [15,20),\ j \in \{Alpha,\ Delta\},\ k = 0 \\ 0 & \text{otherwise,} \end{cases} \tag{213}$$

where

$$f_j(t) = \begin{cases} \left(\left\lceil \frac{t_j}{dt} \right\rceil - \frac{t_j}{dt}\right) & \text{if } t = dt \left\lfloor \frac{t_j}{dt} \right\rfloor \\ 1 & \text{if } dt \left\lfloor \frac{t_j}{dt} \right\rfloor < t < dt \left\lfloor \frac{t_j}{dt} \right\rfloor + \nu_j \\ \left(\frac{t_j}{dt} - \left\lfloor \frac{t_j}{dt} \right\rfloor\right) & \text{if } t = dt \left\lfloor \frac{t_j}{dt} \right\rfloor + \nu_j \\ 0 & \text{otherwise.} \end{cases} \tag{214}$$

where $\lfloor . \rfloor$ and $\lceil . \rceil$ denote the floor and ceiling functions respectively.

#### 4.4 Observation process

To describe the epidemic in each NHS region, we fitted our model to time series data on hospital admissions, hospital ward occupancy (both in general beds and in ICU beds), deaths in hospitals, deaths in care homes, population serological surveys, PCR testing data and Variant and Mutation (VAM) data (see Table S1).

#### 4.4.1 Notation for distributions used in this section

If $X \sim \text{Binom}(n, p)$, then $X$ follows a binomial distribution with mean $np$ and variance $np(1-p)$, such that

$$P(X=x) = P_{\text{Binom}}(x|n,\ p) = \binom{n}{x} p^x (1-p)^{(n-x)}. \tag{215}$$

If $Y \sim \text{NegBinom}(m, \kappa)$, then $Y$ follows a negative binomial distribution with mean $m$ and shape $\kappa$, such that

$$P(Y=y) = P_{\text{NegBinom}}(y|m,\kappa) = \frac{\Gamma(\kappa+y)}{y!\Gamma(\kappa)} \left(\frac{\kappa}{\kappa+m}\right)^{\kappa} \left(\frac{m}{\kappa+m}\right)^{y}, \tag{216}$$

where $\Gamma(x)$ is the gamma function. The variance of $Y$ is $m + m^2/\kappa$.

If $Z \sim \text{BetaBinom}(n, \omega, \rho)$, then $Z$ follows a beta-binomial distribution with size $n$, mean probability $\omega$ and overdispersion parameter $\rho$, such that

$$P(Z=z) = P_{\text{BetaBinom}}(z|n,\omega,\rho) = \binom{n}{z} \frac{B(z+a, n-z+b)}{B(a,b)}, \tag{217}$$

where $a = \omega\left(\frac{1-\rho}{\rho}\right)$, $b = (1-\omega)\left(\frac{1-\rho}{\rho}\right)$ and $B(a,b)$ is the beta function. The mean of Z is $n\omega$ and the variance is $n\omega(1-\omega)[1+(n-1)\rho]$.

In the following, we use $t$ to represent a day with observations. Note that different data streams had different sets of days with observations.

#### 4.4.2 Hospital admissions and new diagnoses in hospital

We represented the daily number of confirmed COVID-19 hospital admissions and new diagnoses for existing hospitalised cases, $Y_{adm}(t)$, as the observed realisations of an underlying hidden Markov process, $X_{adm}(t)$, defined as:

$$\begin{aligned} X_{adm}\ (t) := \sum_i \sum_j \sum_k \Big( & d^{i,j,k}_{I_C,H_{R^*}} + d^{i,j,k}_{I_C,H_{D^*}} + d^{i,j,k}_{I_C,ICU_{pre^*}} + d^{i,j,k}_{H_R,H_{R^*}} + d^{i,j,k}_{ICU_{pre},ICU_{pre^*}} + d^{i,j,k}_{ICU_{W_R},ICU_{W_{R^*}}} \\ & + d^{i,j,k}_{ICU_{W_D},ICU_{W_D^*}} + d^{i,j,k}_{W_D,W_{D^*}} + d^{i,j,k,1,1}_{H_D H_{D^*}} + d^{i,j,k,1,2}_{H_D,H_{D^*}} + d^{i,j,k,2,2}_{H_D,H_{D^*}} + d^{i,j,k,1,1}_{ICU_D,ICU_{D^*}} \\ & + d^{i,j,k,1,2}_{ICU_D,ICU_{D^*}} + d^{i,j,k,2,2}_{ICU_D,ICU_{D^*}} + d^{i,j,k,1,1}_{W_R,W_{R^*}} + d^{i,j,k,1,2}_{W_R,W_{R^*}} + d^{i,j,k,2,2}_{W_R,W_{R^*}} + d^{i,j,k}_{ICU_{pre},ICU_{W_{R^*}}} \\ & + d^{i,j,k}_{ICU_{pre},ICU_{W_{D^*}}} + d^{i,j,k}_{ICU_{pre},ICU_{D^*}} + d^{i,j,k}_{ICU_{W_{D^*}},W_{D^*}} + d^{i,j,k}_{ICU_{W_{R^*}},W_{R^*}} \Big) \end{aligned} \tag{218}$$

which was related to the data via a reporting distribution:

$$Y_{adm}(t) \sim \text{NegBinom}\left(X_{adm}(t), \kappa_H\right) \tag{219}$$

We allow for overdispersion in the observation process to account for noise in the underlying data streams, for example due to day-of-week effects on data collection. We fit the overdispersion parameter $\alpha_H = \frac{1}{\kappa_H}$ , which we use for all hospital data streams (including general hospital bed occupancy, ICU bed occupancy, and hospital death data which contribute equal weight to the overall likelihood – all of which we describe below).

The contribution to the likelihood of the data on hospital admissions and new diagnoses in hospital was therefore:

$$\mathscr{L}_{adm} = \prod_t P_{\text{NegBinom}}\left(Y_{adm}(t)\,|X_{adm}\,(t),\ \kappa_H\right) \tag{220}$$

#### 4.4.3 Hospital bed occupancy by confirmed COVID-19 cases

The model predicted general hospital bed occupancy by confirmed COVID-19 cases, $X_{hosp}(t)$ as:

$$X_{hosp}(t) := \sum_i \sum_j \sum_k \left(H_{R^*}^{i,j,k}(t) + H_{D^*}^{i,j,k,1}(t) + H_{D^*}^{i,j,k,2}(t) + ICU_{pre^*}^{i,j,k}(t) + W_{D^*}^{i,j,k}(t) + W_{R^*}^{i,j,k,1}(t) + W_{R^*}^{i,j,k,2}(t)\right), \tag{221}$$

which was related to the observed daily general bed-occupancy via a reporting distribution:

$$Y_{hosp}\ (t) \sim \text{NegBinom}\left(X_{hosp}(t), \kappa_H\right). \tag{222}$$

Similarly, the model predicted ICU bed occupancy by confirmed COVID-19 cases, $X_{ICU}(t)$ as:

$$X_{ICU}(t) := \sum_i \sum_j \sum_k \left(ICU_{W_{R^*}}^{i,j,k}(t) + ICU_{W_{D^*}}^{i,j,k}(t) + ICU_{D^*}^{i,j,k,1}(t) + ICU_{D^*}^{i,j,k,2}(t)\right), \tag{223}$$

which was related to the observed daily ICU bed-occupancy via a reporting distribution:

$$Y_{ICU}(t) \sim \text{NegBinom}\left(X_{ICU}(t), \kappa_H\right). \tag{224}$$

The overall contribution to the likelihood of the data on general bed and ICU bed occupancy was:

$$\begin{aligned} \mathscr{L}_{beds} = \prod_t P_{\text{NegBinom}}\left(Y_{hosp}(t)\,|X_{hosp}(t), \kappa_H\right) \\ \times \prod_t P_{\text{NegBinom}}\left(Y_{ICU}(t)\,|X_{ICU}(t), \kappa_H\right). \end{aligned} \tag{225}$$

#### 4.4.4 Hospital, community and care homes COVID-19 deaths

The reported number of daily COVID-19 deaths in hospitals, $Y_{hosp_D}\ (t)$ was considered as the observed realisation of an underlying hidden Markov process, $X_{hosp_D}(t)$, defined as:

$$X_{hosp_D}(t) := \sum_i \sum_j \sum_k \left(d_{H_D,D}^{i,j,k} + d_{H_{D^*},D}^{i,j,k} + d_{ICU_D,D}^{i,j,k} + d_{ICU_{D^*},D}^{i,j,k} + d_{W_D,D}^{i,j,k} + d_{W_{D^*},D}^{i,j,k}\right), \tag{226}$$

which was related to the data via a reporting distribution:

$$Y_{hosp_D}(t) \sim \text{NegBinom}\left(X_{hosp_D}(t), \kappa_H\right). \tag{227}$$

Similarly, we represented the reported number of daily COVID-19 deaths in the community, $Y_{comm_D}(t)$, as the observed realisations of an underlying hidden Markov process, $X_{comm_D}(t)$, defined as:

$$X_{comm_D}(t) := \sum_{i \neq CHR} \sum_{j} \sum_{k} d^{i,j,k}_{G_D,D}, \tag{228}$$

which was related to the data via a reporting distribution:

$$Y_{comm_D}(t) \sim \text{NegBinom}\left(X_{comm_D}(t), \kappa_D\right). \tag{229}$$

Finally, we represented the reported number of daily COVID-19 deaths in care homes, $Y_{CH_D}(t)$, as the observed realisations of an underlying hidden Markov process, $X_{CH_D}(t)$, defined as:

$$X_{CH_D}(t) := d^{CHR}_{G_D,D}, \tag{230}$$

which was related to the data via a reporting distribution:

$$Y_{CH_D}(t) \sim \text{NegBinom}\left(X_{CH_D}(t), \kappa_D\right). \tag{231}$$

We fit the overdispersion parameter $\alpha_D = \frac{1}{\kappa_D}$ , for the community and care home death data streams (note that we use the same overdispersion for hospital deaths as we do for other hospital data streams).

The overall contribution to the likelihood of the data on COVID-19 deaths in hospital, in the community and in care homes was:

$$\begin{aligned} \mathscr{L}_{deaths} = & \prod_t P_{\text{NegBinom}}\left(Y_{hosp_D}(t) \,|X_{hosp_D}(t), \kappa_H\right) \\ & \times \prod_t P_{\text{NegBinom}}\left(Y_{comm_D}(t) \,|X_{comm_D}(t), \kappa_D\right) \\ & \times \prod_t P_{\text{NegBinom}}\left(Y_{CH_D}(t) \,|X_{CH_D}(t), \kappa_D\right). \end{aligned} \tag{232}$$

#### 4.4.5 Serosurveys

We model serological testing of all individuals aged 15-64 inclusive, and define the resulting number of seropositive and seronegative individuals (were all individuals aged 15-65 to be tested) from serology flow $j$ (where $j = 1$ corresponds to EuroImmun and $j = 2$ to Roche N), as:

$$X_{sero^j_{pos}}(t) := \sum_{i=[15,20)}^{[60,65)} T^i_{sero^j_{pos}}(t) \tag{233}$$

$$X_{sero^j_{neg}}(t) := \left(\sum_{i=[15,20)}^{[60,65)} N^i\right) - X_{sero^j_{pos}}(t). \tag{234}$$

We compared the observed number of seropositive results, $Y_{sero^j_{pos}}(t)$, with that predicted by our model, allowing for i) the sample size of each serological survey, $Y_{sero^j_{test}}(t)$ and ii) imperfect sensitivity ($p_{sero_{sens}}$) and specificity ($p_{sero_{spec}}$) of the serological assay:

$$Y_{sero^j_{pos}}(t) \sim \text{Binom}\left(Y_{sero^j_{test}}(t), \omega_{sero_{pos}}(t)\right) \tag{235}$$

where:

$$\omega_{sero^j_{pos}}(t) := \frac{p_{sero_{sens}} X_{sero^j_{pos}}(t) + \left(1 - p_{sero_{spec}}\right) X_{sero^j_{neg}}(t)}{X_{sero^j_{pos}}(t) + X_{sero^j_{neg}}(t)}. \tag{236}$$

The contribution to the likelihood of the serosurvey data was:

$$\mathscr{L}_{sero} = \prod_t \prod_{j=1,2} P_{\text{Binom}}\left(Y_{sero^j_{pos}}(t) \middle| X_{sero^j_{test}}(t), \omega_{sero^j_{pos}}(t)\right) \tag{237}$$

#### 4.4.6 PCR testing

As described in the data section (section 4), we fitted the model to PCR testing data from two separate sources:

- Pillar 2 testing: government community testing programme, which recommends that symptomatic individuals in the community with COVID-19 symptoms are tested [9].
- the REACT-1 study, which aims to quantify the prevalence of SARS-CoV-2 in a random sample of the England population on an ongoing basis [10].

We only use Pillar 2 PCR test results for individuals aged 25 and over (we assume this includes all care home workers and residents). We assume that individuals who get tested through Pillar 2 PCR testing are either newly symptomatic SARS-CoV-2 cases (who will test positive):

$$X_{P2_{pos}}(t) := \sum_{i=[25,30)}^{CHW} \sum_j \sum_k d^{i,j,k}_{I_P,I_{C_1}} \tag{238}$$

or non-SARS-CoV-2 cases who have symptoms consistent with COVID-19 (who will test negative):

$$X_{P2_{neg}}(t) := g(t) \left( \left( \sum_{i=[25,30)}^{CHW} N^i \right) - X_{P2_{pos}}(t) \right), \tag{239}$$

where

$$g(t) = \begin{cases} p_{NC} & \text{if } t \text{ is a weekday} \\ p^{weekend}_{NC} & \text{if } t \text{ is a weekend day} \end{cases} \tag{240}$$

is the probability of non SARS-CoV-2 cases having symptoms consistent with COVID-19 that might lead them to get a PCR test.

We compared the observed number of positive PCR tests, $Y_{P2_{pos}}(t)$ with that predicted by our model, accounting for the number of PCR tests conducted each day under pillar 2, $Y_{P2_{test}}(t)$, by calculating the probability of a positive PCR result (assuming perfect sensitivity and specificity of the PCR test):

$$\omega_{P2_{pos}}(t) := \frac{X_{P2_{pos}}(t)}{X_{P2_{pos}}(t) + X_{P2_{neg}}(t)} \tag{241}$$

People may seek PCR tests for many reasons and thus the pillar 2 data are subject to competing biases. We therefore allowed for an over-dispersion parameter $\rho_{P2_{test}}$, which we fitted separately for each region in the modelling framework:

$$Y_{P2_{pos}}(t) \sim \text{BetaBinom}\left(Y_{P2_{test}}(t), \omega_{P2_{pos}}(t), \rho_{P2_{test}}\right). \tag{242}$$

We incorporated the REACT-1 PCR testing data into the likelihood analogously to the serology data, by considering the model-predicted number of PCR-positives, $X_{R1_{pos}}(t)$, and PCR-negatives, $X_{R1_{neg}}(t)$, were all individuals aged over five and not resident in a care home to be tested:

$$X_{R1_{pos}}(t) := \sum_{i \neq [0,4), CHR} T^{i}_{PCR_{pos}}(t), \tag{243}$$

$$X_{R1_{neg}}(t) := \left(\sum_{i \neq [0,4), CHR} N^{i}\right) - X_{R1_{pos}}(t). \tag{244}$$

We compared the daily number of positive results observed in REACT-1, $Y_{R1_{pos}}(t)$, given the number of people tested on that day, $Y_{R1_{test}}(t)$, to our model predictions, by calculating the probability of a positive result, assuming perfect sensitivity and specificity of the REACT-1 assay:

$$\omega_{R1_{pos}}(t) := \frac{X_{R1_{pos}}(t)}{X_{R1_{pos}}(t) + X_{R1_{neg}}(t)} \tag{245}$$

so

$$Y_{R1_{pos}}(t) \sim \text{Binom}\left(Y_{R1_{test}}(t), \omega_{R1_{pos}}(t)\right). \tag{246}$$

The contribution to the likelihood of the PCR testing data was:

$$\begin{aligned} \mathscr{L}_{PCR} = &\prod_{t} P_{\text{BetaBinom}}\left(Y_{P2_{pos}}(t) \,\middle|\, Y_{P2_{test}}(t), \omega_{P2_{pos}}(t), \rho_{P2_{test}}\right) \\ &\times \prod_{t} P_{\text{Binom}}\left(Y_{R1_{pos}}(t) \,\middle|\, Y_{R1_{test}}(t), \omega_{R1_{pos}}(t)\right) \end{aligned} \tag{247}$$

#### 4.4.7 Variant and Mutation data

To inform the replacement of the Alpha variant by the Delta variant, we fitted to Variant and Mutation (VAM) data. We assume that samples tested for VAM are newly symptomatic cases (across all groups), with the number for $Alpha$ and for $Delta$ given by

$$X_{VAM_{Alpha}}(t) := \sum_{i}\sum_{k} d^{i,Alpha,k}_{I_P,I_{C_1}} \tag{248}$$

$$X_{VAM_{Delta}}(t) := \sum_{i}\sum_{k} d^{i,Delta,k}_{I_P,I_{C_1}} + d^{i,Alpha+Delta,k}_{I_P,I_{C_1}} \tag{249}$$

We compared the observed number of Delta VAM tests, $Y_{VAM_{Delta}}(t)$ with that predicted by our model, accounting for the total number of VAM tests conducted each day, $Y_{VAM_{test}}(t)$, by calculating the probability of a Delta VAM test result:

$$\omega_{VAM_{Delta}}(t) := \frac{X_{VAM_{Delta}}(t)}{X_{VAM_{Alpha}}(t) + X_{VAM_{Delta}}(t)} \tag{250}$$

so

$$Y_{VAM_{Delta}}(t) \sim \text{Binom}\left(Y_{VAM_{test}}(t), \omega_{VAM_{Delta}}(t)\right). \tag{251}$$

The contribution to the likelihood of the VAM data was:

$$\mathscr{L}_{VAM} = \prod_t P_{\text{Binom}}\left(Y_{VAM_{Delta}}(t) \left| Y_{VAM_{test}}(t), \omega_{VAM_{Delta}}(t)\right.\right) \tag{252}$$

#### 4.4.8 Full likelihood

The overall likelihood was then calculated as the product of the likelihoods of the individual observations, i.e.:

$$\mathscr{L} = \mathscr{L}_{adm} \times \mathscr{L}_{beds} \times \mathscr{L}_{deaths} \times \mathscr{L}_{sero} \times \mathscr{L}_{PCR} \times \mathscr{L}_{VAM}. \tag{253}$$

### 4.5 Reproduction number

Both $R^j(t)$ and $R_e^j(t)$ are calculated using next generation matrix (NGM) methods [47], and only consider mixing in the general population, i.e. we do not consider the $CHW$ and $CHR$ age categories for that calculation.

Note that in this calculation only, we make a simplifying assumption that individuals cannot change vaccine strata between initial infection and the end of their infectious period (or death).

To compute the next generation matrix, we calculated the mean duration of infectiousness weighted by infectivity (asymptomatic individuals are less infectious than symptomatic individuals by factor $\theta_{I_A}$) for an individual in group $i$ and vaccine stage $k$, $\Delta_I^{i,k}$:

$$\Delta_I^{i,k} = \theta_{I_A}\left(1 - p_C^{i,j,k}\right)\mathbb{E}\left[\tau_{I_A}\right] + p_C^{i,j,k}\left(\mathbb{E}\left[\tau_{I_P}\right] + \mathbb{E}\left[\tau_{I_{C_1}}\right]\right). \tag{254}$$

Note that $\Delta_I^{i,k}$ does not depend on $j$, as we assume the same duration spent in compartments and probability of being symptomatic between variants. The next generation matrices for the variants were calculated as,

$$NGM_{i,i'}^{Alpha}(t) = m_{i,i'}(t)\Delta_I^{i,0}N^{i'}, \tag{255}$$

$$NGM_{i,i'}^{Delta}(t) = m_{i,i'}(t)\xi^{i,Delta,0}\Delta_I^{i,0}N^{i'}, \tag{256}$$

where $\xi$ is the infectivity of an individual (fully defined in eq. (3)), $N^i$ is the total population of age group $i$, and with $R^j(t)$ taken as the dominant eigenvalue of the 17 by 17 matrix $NGM^j(t)$. The element $NGM_{i,i'}^j(t)$ is therefore defined as the average number of secondary cases that an individual in age group $i'$ infected with variant $j$ at time $t$ would generate among age group $i$.

The effective next generation matrices for the variants were calculated as

$$NGM_{D(i,k),D(i',k')}^{Alpha,e}(t) = m_{i,i'}(t)\chi^{i,Alpha,k}\xi^{i,Alpha,k'}\Delta_I^{i,k}S^{i',k'}(t), \tag{257}$$

$$NGM_{D(i,k),D(i',k')}^{Delta,e}(t) = m_{i,i'}(t)\chi^{i,Delta,k}\xi^{i,Delta,k'}\Delta_I^{i,k}\left(S^{i',k'}(t) + (1-\eta)R^{i',Alpha,k'}(t)\right), \tag{258}$$

where $D : \{[0-4],[5-9],\ldots,[75-79],80+\} \times \{0,1,2,3\} \rightarrow \{1,2,\ldots,68\}$ is a one-to-one mapping. Then $R_e^j(t)$ is taken as the dominant eigenvalue of the 68 by 68 matrix $NGM^{j,e}(t)$ .

We calculate the reproduction numbers weighted by the two variants as

$$R(t) = \frac{w_{Alpha}(t)R^{Alpha}(t) + w_{Delta}(t)R^{Delta}(t)}{w_{Alpha}(t) + w_{Delta}(t)} \tag{259}$$

$$R_e(t) = \frac{w_{Alpha}(t)R_e^{Alpha}(t) + w_{Delta}(t)R_e^{Delta}(t)}{w_{Alpha}(t) + w_{Delta}(t)}, \tag{260}$$

where the weightings $w_j(t)$ are weightings based on the infectious prevalence of each variant (accounting for the baseline relative infectivity of each compartment), such that for $j = Alpha, Delta$,

$$w_j(t) = \sum_i \sum_k \left( \theta_{I_A} I_A^{i,j,k}(t) + I_P^{i,j,k}(t) + I_{C_1}^{i,j,k}(t) \right). \tag{261}$$

### 4.6 Fixed parameters

We used parameter values calibrated to data from 13th September 2021. We assume that the performance of the tests (PCR and serological assays) are the same for all variants [2].

| Parameter | Definition | Value | Source |
|---|---|---|---|
| $1/\gamma_U$ | Mean time to confirmation of SARS-CoV-2 diagnosis within hospital. | 3 days | [48] |
| $1/\gamma_R$ | Mean duration of natural immunity following infection. | 6 years | [8] |
| $p_{sero_{pos}}$ | Probability of seroconversion following infection. | 0.85 | [49] |
| $1/\gamma_{sero_{pre}}$ | Mean time to seroconversion from onset of infectiousness. | 13 days | [50] |
| $1/\gamma_{sero^1_{pos}}$ | Mean duration of seropositivity (Euroimmun assay). | 200 days | [49, 51, 52] |
| $1/\gamma_{sero^2_{pos}}$ | Mean duration of seropositivity (Roche N). | 400 days | [49, 51, 52] |
| $p_{sero_{spec}}$ | Specificity of serology test. | 0.99 | [49] |
| $p_{sero_{sens}}$ | Sensitivity of serology test. | 1 | Assumed |
| $\eta$ | Probability of cross-immunity to $Delta$ following infection with $Alpha$. | 0.85 | [53, 17] |
| $\theta_{I_A}$ | Infectivity of an asymptomatic individual, relative to a symptomatic individual. | 0.223 | [1] |

**Table S7:** Fixed model parameter notations, values, and evidence-base.

#### 4.7 Prior distributions

Prior distributions are described in table S8. Informative prior distributions for the single strain model are the same as prior distributions in the model given in [1, 2]. In the absence of evidence from the literature (or because existing evidence has been derived from the same datasets we use in our study), uninformative or weakly informative prior distributions have been chosen for the two-strain model; the prior for $t_{Delta}$ covers a wide period of time spanning over more than two months and the assumption of $\sigma$, and the prior for the Delta transmission advantage is assumed uniform between [0,3]."

**Table S8:** Inferred model parameter notations and prior distributions. Region codes: NW = North West, NEY = North East and Yorkshire, MID = Midlands, EE = East of England, LON = London, SW = South West, SE = South East

| | Description | Region | Prior distribution |
|---|---|---|---|
| $t_{Alpha}$ | Start date of regional outbreak (dd/mm/20) | All | $U[01-01,03-15]$ |
| $t_{Delta}$ | Delta seeding date (dd/mm/21) | All | $U[03-10,05-31]$ |
| $\sigma$ | Delta transmission advantage | All | $U(0,3)$ |
| $\beta(t)$ | Transmission rate (pp) at $t = dd/mm/yy$ | | |
| $\beta_1$ | 16/03/20: PM advises WFH and essential travel only | All | $\Gamma(136, 0.0008)$ |
| $\beta_2$ | 23/03/20: PM announces lockdown 1 | All | $\Gamma(3.73, 0.0154)$ |
| $\beta_3$ | 25/03/20: Lockdown 1 into full effect | All | $\Gamma(4.25, 0.0120)$ |
| $\beta_4$ | 11/05/20: Initial easing of lockdown 1 | All | $\Gamma(4.25, 0.0120)$ |
| $\beta_5$ | 15/06/20: Non-essential shops re-open | All | $\Gamma(4.25, 0.0120)$ |
| $\beta_6$ | 04/07/20: Hospitality re-opens | All | $\Gamma(4.25, 0.0120)$ |
| $\beta_7$ | 01/08/20: "Eat out to help out" scheme starts | All | $\Gamma(4.25, 0.0120)$ |
| $\beta_8$ | 01/09/20: Schools and universities re-open | All | $\Gamma(4.25, 0.0120)$ |
| $\beta_9$ | 14/09/20: "Rule of six" introduced | All | $\Gamma(4.25, 0.0120)$ |
| $\beta_{10}$ | 14/10/20: Tiered system introduced | All | $\Gamma(4.25, 0.0120)$ |
| $\beta_{11}$ | 31/10/20: Lockdown 2 announced | All | $\Gamma(4.25, 0.0120)$ |
| $\beta_{12}$ | 05/11/20: Lockdown 2 starts | All | $\Gamma(4.25, 0.0120)$ |
| $\beta_{13}$ | 02/12/20: Lockdown 2 ends | All | $\Gamma(4.25, 0.0120)$ |
| $\beta_{14}$ | 18/12/20: School holidays start | All | $\Gamma(4.25, 0.0120)$ |
| $\beta_{15}$ | 25/12/20: Last day of holiday season relaxation | All | $\Gamma(4.25, 0.0120)$ |
| $\beta_{16}$ | 05/01/21: Lockdown 3 starts | All | $\Gamma(4.25, 0.0120)$ |
| $\beta_{17}$ | 08/03/21: Roadmap step one - schools reopen | All | $\Gamma(4.25, 0.0120)$ |
| $\beta_{18}$ | 01/04/21: School holidays | All | $\Gamma(4.25, 0.0120)$ |
| $\beta_{19}$ | 19/04/21: Roadmap step two - outdoor rule of 6 (12/04) and schools re-open (19/04) | All | $\Gamma(2.72, 0.0292)$ |
| $\beta_{20}$ | 17/05/21: Roadmap step three - Indoor hospitality opens | All | $\Gamma(2.72, 0.0292)$ |
| $\beta_{21}$ | 21/06/21: Wedding and care home restrictions eased | All | $\Gamma(2.72, 0.0292)$ |
| $\beta_{22}$ | 03/07/21: Euro 2020 quarter finals (cited as significant influence [54]) | All | $\Gamma(2.72, 0.0292)$ |
| $\beta_{23}$ | 11/07/21: End of Euros football tournament | All | $\Gamma(2.72, 0.0292)$ |
| $\beta_{24}$ | 19/07/21: Full lift of NPIs | All | $\Gamma(2.72, 0.0292)$ |

Continued on next page

| Table S8 – continued from previous page | | | |
| --- | --- | --- | --- |
| | Description | Region | Prior distribution |
| $\beta_{25}$ | 15/08/21: Summer festivals/holidays | All | $\Gamma(2.72, 0.0292)$ |
| $\beta_{26}$ | 01/09/21: Schools return | All | $\Gamma(2.72, 0.0292)$ |
| $\beta_{27}$ | 13/09/21: End of fits | All | $\Gamma(2.72, 0.0292)$ |
| $\varepsilon$ | Relative reduction in contacts between CHR and the general population | All | $B(1,1)$ |
| $m_{CHW}$ | Transmission rate between CHR and CHW | NW | $\Gamma(5, 4.3\times10^{-7})$ |
| | | NEY | $\Gamma(5, 3.7\times10^{-7})$ |
| | | MID | $\Gamma(5, 2.9\times10^{-7})$ |
| | | EE | $\Gamma(5, 5.2\times10^{-7})$ |
| | | LON | $\Gamma(5, 7.6\times10^{-7})$ |
| | | SW | $\Gamma(5, 4.9\times10^{-7})$ |
| | | SE | $\Gamma(5, 3.1\times10^{-7})$ |
| $m_{CHR}$ | Transmission rate among CHR | NW | $\Gamma(5, 4.3\times10^{-7})$ |
| | | NEY | $\Gamma(5, 3.7\times10^{-7})$ |
| | | MID | $\Gamma(5, 2.9\times10^{-7})$ |
| | | EE | $\Gamma(5, 5.2\times10^{-7})$ |
| | | LON | $\Gamma(5, 7.6\times10^{-7})$ |
| | | SW | $\Gamma(5, 4.9\times10^{-7})$ |
| | | SE | $\Gamma(5, 3.1\times10^{-7})$ |
| $p^{max}_{H,1}, p^{max}_{H,2}, p^{max}_{H,3}$ | The probability of symptomatic individuals developing serious disease requiring hospitalisation, for the group with the largest probability at different timepoints (see Section 4.3.2) | All | $B(15.8, 5.28)$ |
| $p^{max}_{G_D}$ | Probability of death in the community given disease severe enough for hospitalisation | All | $B(1,1)$ |
| $p^{CHR}_{G_D}$ | Probability of death in CHR given disease severe enough for hospitalisation | All | $B(1,1)$ |
| $p^{max}_{ICU,1}, p^{max}_{ICU,2}$ | Probability of triage to ICU for new hospital admissions, for the group with the largest probability at different timepoints (see Section 4.3.2) | All | $B(13.9, 43.9)$ |
| $p^{max}_{H_D}$ | Initial probability of death for general inpatients | All | $B(42.1, 50.1)$ |
| $p^{max}_{ICU_D}$ | Initial probability of death for ICU inpatients | All | $B(60.2, 29.3)$ |
| $p^{max}_{W_D}$ | Initial probability of death for stepdown inpatients | All | $B(28.7, 52.1)$ |
| $\mu_{D,1}, \mu_{D,2}$ | Hospital mortality multipliers due to changes in clinical care at different timepoints (see Section 4.3.2) | All | $B(1,1)$ |
| $\pi^{Delta}_{H}$ | Multiplier for the probability of disease severe enough for hospitalisation for Delta, relative to Alpha | All | $U(0,3)$ |
| $\pi^{Delta}_{ICU}$ | Multiplier for the probability of ICU admission for new hospital admissions for Delta, relative to Alpha | All | $U(0,3)$ |
| $\pi^{Delta}_{D}$ | Multiplier for mortality given disease severe enough for hospitalisation for Delta, relative to Alpha | All | $U(0,3)$ |
| $\mu_{\gamma_H,1}, \mu_{\gamma_H,2}, \mu_{\gamma_H,3} \mu_{\gamma_H,4},$ | Mean duration multipliers for non-ICU hospital compartments at different timepoints (see Section 4.3.2) | All | $\Gamma(1000, 1.001)$ |

Continued on next page

| Table S8 – continued from previous page | Description | Region | Prior distribution |
|---|---|---|---|
| $p_{NC}$ | Prevalence of non-COVID symptomatic illness that could lead to getting a PCR test | All | $B(1,1)$ |
| $p_{NC}^{weekend}$ | Prevalence of non-COVID symptomatic illness that could lead to getting a PCR test on a weekend | All | $B(1,1)$ |
| $\rho_{P2_{test}}$ | Overdispersion of PCR positivity | All | $B(1,1)$ |
| $\alpha_H$ | Overdispersion for hospital data streams | All | $B(1,1)$ |
| $\alpha_D$ | Overdispersion for death data streams | All | $B(1,1)$ |

#### 4.8 Running the model

The model is fitted to multiple data streams up to 13th September 2021, capturing the entirety of the SARS-CoV-2 epidemic up to (but excluding) the introduction of booster vaccines. The model is run under baseline assumptions reflected by the fixed parameters (Table S7) and VE (Table S3) tables.

### 5 Counterfactual Analysis

The previous sections provided methodological details of our model structure and how we fit to existing epidemiological data. This section outlines information and assumptions made for simulating counterfactual scenarios using information inferred from the model fits.

We assume that vaccine roll-out is exactly as that observed during the study period in terms of doses distributed per day. We also assume that the proportion of vaccine type (AZ or mRNA vaccines) administered by age group also remained the same as that observed. Transmissibility also remains the same as what is estimated from the model fits.

Simulated scenarios therefore depend on assumptions regarding: the dosing interval between first and second doses, the mean VE waning time, and the vaccine effectiveness which we discuss in sections 5.1 and 5.2 respectively.

#### 5.1 Vaccine dosing interval

We ran counterfactual analyses to explore the impact that different vaccine dosing intervals had on the epidemic in England. We maintained a 3-week gap throughout the study period and compared this with the reported/observed epidemic trajectory with a longer dosing interval due to the 12-week gap recommended by the JCVI.

#### 5.2 Sensitivity analyses

In addition to our main baseline counterfactual analysis, we run a number of sensitivity analyses to explore:

1. *Uncertainty in VE when a 3-week rather than 12-week interval is used.* While our baseline counterfactual assumes that VE remains identical between the 3-week and 12-week interval scenarios, Amirthalingam et al. [24] suggests that second dose VE could be slightly lower following a shorter dosing interval. We conduct sensitivity analyses for different changes to VE between the two scenarios.
2. *General uncertainty in VE against the Delta variant.* The model is refit and simulated for a range of VE values.
3. *Uncertainty in the level of waning vaccine-induced immunity.* The model is refit for a range of VE in the reduced protection after second dose ($V_3$) state.
4. *Uncertainty in the timing of vaccine-induced waning immunity.* The shorter (3-week) dosing interval may impact the mean waning duration post dose 2. Thus, a sensitivity analysis is conducted with a longer mean waning time.
5. *Uncertainty in waning of immunity following the first vaccine dose.* Because waning of VE post dose 1 is not explicitly modelled, the first dose VE values listed represent the mean VE afforded across the 12 weeks between doses. A shorter dosing interval could plausibly increase the mean VE post dose 1 due to less waning of post dose 1 VE occurring. We consider a sensitivity scenario whereby first dose VE is increased in the 3-week delay counterfactual.
6. *Immediate vaccine-induced protection after 1st dose.* Extreme assumption on time to effectiveness of vaccine-induced protection.

Table S9 summarises each of the sensitivity analyses explored and table S10 and table S11 the VE against Alpha and Delta for each analyses respectively.

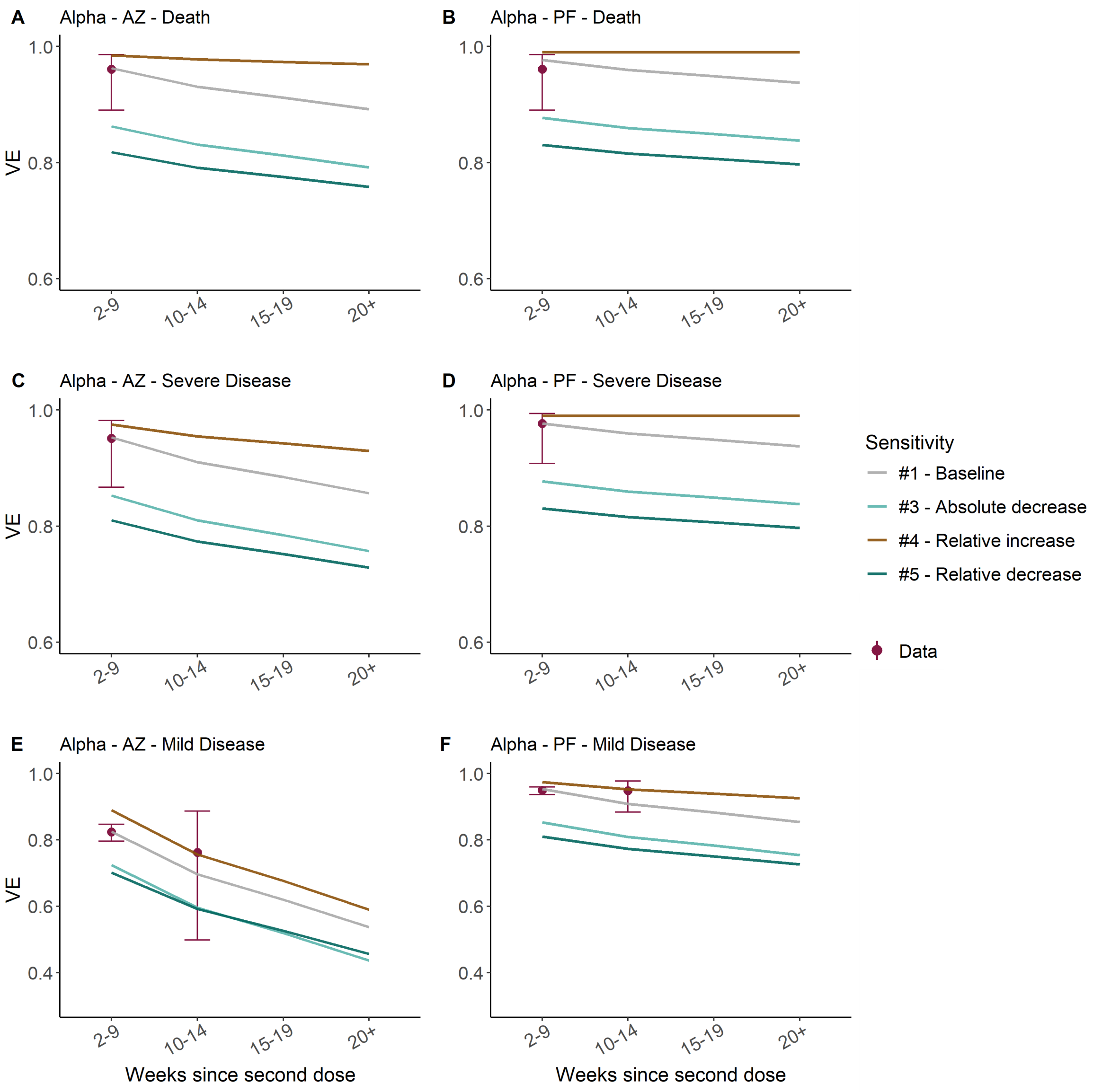

**Figure S9:** Vaccine effectiveness in weeks since second dose of AstraZeneca (AZ, left column) and Pfizer (PF, right column) vaccines against Alpha (see supplementary materials for values against Alpha) for death (top), severe disease, (middle) and mild disease/infection (bottom). We assume the same protection against infection and mild disease. Turquoise points show the VE estimates from Andrews et al, and the purple points our model assumptions. We assumed that the Moderna vaccine has the same VE as PF.

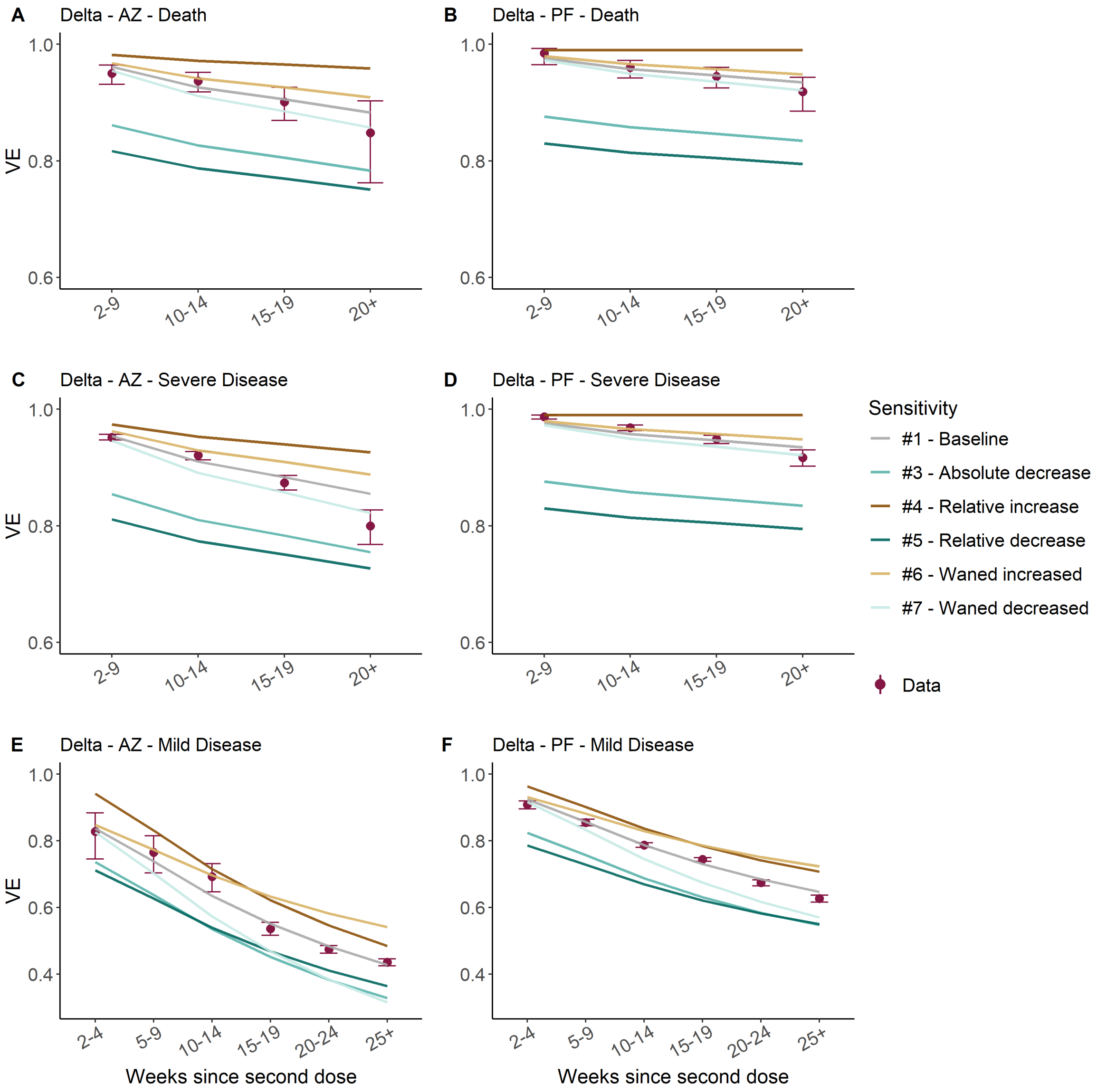

**Figure S10:** Vaccine effectiveness in weeks since second dose of AstraZeneca (AZ, left column) and Pfizer (PF, right column) vaccines against Delta (see supplementary materials for values against Alpha) for death (top), severe disease, (middle) and mild disease/infection (bottom). We assume the same protection against infection and mild disease. Turquoise points show the VE estimates from Andrews et al, and the purple points our model assumptions. We assumed that the Moderna vaccine has the same VE as PF.

| | Analysis | Strategy | Doses | Variant | Description and endpoints | Informed by |
|---|---|---|---|---|---|---|
| *1* | *Baseline* | - | | - | Equal VE for 12- and 3-week strategies | Conservative baseline |
| *2* | *3-week VE* | *3-week* | 2nd full protection and reduced protection | Alpha/Delta | Absolute 10% reduction in VE vs infection/mild disease for 2nd dose full protection and reduced protection VE | [24] |
| *3* | *3-week VE* | *3-week* | 2nd full protection and reduced protection | Alpha/Delta | Absolute 10% reduction vs all end points for 2nd dose full protection and reduced protection VE | Pessimistic scenario applying decrease seen in Amirthalingham et al to all end points |
| *4* | *All VE* | *12- and 3-week* | All | Alpha/Delta | 15% relative increase in VE vs all end points and doses | Arbitrarily higher VE |
| *5* | *All VE* | *12- and 3-week* | All | Alpha/Delta | 15% relative decrease in VE vs all end points and doses | Arbitrarily lower VE |
| *6* | *Waning vaccine immunity* | *12- and 3-week* | 2nd dose reduced protection | Delta | +1/4 VE between second dose full and reduced protection, all end points | Arbitrarily higher VE after 24 weeks |
| *7* | *Waning vaccine immunity* | *12- and 3-week* | 2nd dose reduced protection | Delta | -1/4 VE between second dose full and reduced protection, all end points | Arbitrarily lower VE after 24 weeks |
| *8* | *First dose waning* | *3-week* | First | Alpha/Delta | To account for first dose waning in the 12-week strategy, increase first dose 3-week VE across all end points by absolute 10% | Difference in dosing intervals |
| *9* | *Immediate protection* | - | 1st | Alpha/Delta | Immediate vaccine effectiveness upon first dose vaccination with 10% absolute reduction | Assumed |

**Table S9:** Summary of sensitivity analyses explored

**Table S10:** Vaccine effectiveness assumptions against the Alpha variant for three vaccines licensed for use in England. "Baseline" shows our main assumptions, and the other columns show sensitivity analyses assumptions. Cells with no value assumes the same VE as the baseline. Baseline values are the same as in Table S3. See Table S9 for explanation of sensitivity values.

| End point | Dose | Vaccine | *Baseline* | *3w (+10%)* | *3w all (-10%)* | All VE (+15%) | All VE (-15%) | *3w (1st dose)* | Immediate (1st dose -10%) |
|---|---|---|---|---|---|---|---|---|---|
| *Death* | 1 | *AZ* | 88% | | | 99% | 74% | 98% | 78% |
| | | *PF/Mod* | 89% | | | 99% | 76% | 99% | 79% |
| | 2 (full protection) | *AZ* | 99% | | 89% | | 84% | | |
| | | *PF/Mod* | 99% | | 89% | | 84% | | |
| | 2 (reduced protection) | *AZ* | 83% | | 73% | 96% | 71% | | |
| | | *PF/Mod* | 90% | | 80% | 99% | 77% | | |
| *Severe disease* | 1 | *AZ* | 81% | | | 93% | 69% | 91% | 71% |
| | | *PF/Mod* | 89% | | | 99% | 76% | 99% | 79% |
| | 2 (full protection) | *AZ* | 99% | | 89% | | 84% | | |
| | | *PF/Mod* | 99% | | 89% | | 84% | | |
| | 2 (reduced protection) | *AZ* | 77% | | 67% | 89% | 66% | | |
| | | *PF/Mod* | 90% | | 80% | 99% | 77% | | |
| *Mild disease/infection* | 1 | *AZ* | 64% | | | 73% | 54% | 74% | 54% |
| | | *PF/Mod* | 79% | | | 90% | 67% | 89% | 69% |
| | 2 (full protection) | *AZ* | 92% | 82% | 82% | 99% | 78% | | |
| | | *PF/Mod* | 99% | 89% | 89% | | 84% | | |
| | 2 (reduced protection) | *AZ* | 29% | 19% | 19% | 33% | 24% | | |
| | | *PF/Mod* | 77% | 67% | 67% | 88% | 65% | | |
| *Transmission* | 1 | *AZ* | 45% | | | 52% | 38% | 45% | 35% |
| | | *PF/Mod* | 45% | | | 52% | 38% | 45% | 35% |
| | 2 (full protection) | *AZ* | 45% | | 45% | 52% | 38% | | |
| | | *PF/Mod* | 45% | | 45% | 52% | 38% | | |
| | 2 (reduced protection) | *AZ* | 40% | | 40% | 46% | 34% | | |
| | | *PF/Mod* | 40% | | 40% | 46% | 34% | | |

**Table S11:** Vaccine effectiveness assumptions against the Delta variant for three vaccines licensed for use in England. "Baseline" shows our main assumptions, and the other columns show sensitivity analyses assumptions. Cells with no value assumes the same VE as the baseline. *References refer to baseline values. See Table S9 for explanation of sensitivity values.

| End point | Dose | Vaccine | *Baseline* | *3w (-10%)* | *3w all (-10%)* | *All VE (+15%)* | *All VE (-15%)* | *All Waning (x1.25)* | *All Waning (x0.75)* | *3w (1st dose)* | *Immediate* (1st dose -10%) |
|---|---|---|---|---|---|---|---|---|---|---|---|
| *Death* | 1 | *AZ* | 88% | | | 99% | 74% | | | 97% | 78% |
| | | *PF/Mod* | 89% | | | 99% | 76% | | | 99% | 79% |
| | 2 (full protection) | *AZ* | 99% | | 89% | | 84% | | | | |
| | | *PF/Mod* | 99% | | 89% | | 84% | | | | |
| | 2 (reduced protection) | *AZ* | 82% | | 72% | 94% | 70% | 86% | 78% | | |
| | | *PF/Mod* | 90% | | 80% | 99% | 77% | 93% | 88% | | |
| *Severe disease* | 1 | *AZ* | 81% | | | 93% | 69% | | | 91% | 71% |
| | | *PF/Mod* | 89% | | | 99% | 76% | | | 99% | 79% |
| | 2 (full protection) | *AZ* | 99% | | 89% | 99% | 84% | | | | |
| | | *PF/Mod* | 99% | | 89% | | 84% | | | | |
| | 2 (reduced protection) | *AZ* | 77% | | 67% | 89% | 66% | 83% | 72% | | |
| | | *PF/Mod* | 90% | | 80% | 99% | 77% | 93% | 88% | | |
| *Mild disease /infection* | 1 | *AZ* | 51% | | | 59% | 44% | | | 61% | 41% |
| | | *PF/Mod* | 51% | | | 59% | 44% | | | 61% | 41% |
| | 2 (full protection) | *AZ* | 87% | 77% | 77% | 99% | 74% | | | | |
| | | *PF/Mod* | 95% | 85% | 85% | 99% | 81% | | | | |
| | 2 (reduced protection) | *AZ* | 19% | 9% | 9% | 22% | 16% | 36% | 2% | | |
| | | *PF/Mod* | 49% | 39% | 39% | 56% | 41% | 60% | 37% | | |
| *Transmission* | 1 | *AZ* | 33% | | | 38% | 28% | | | 40% | 23% |
| | | *PF/Mod* | 33% | | | 38% | 28% | | | 40% | 23% |
| | 2 (full protection) | *AZ* | 40% | | 33% | 46% | 34% | | | | |
| | | *PF/Mod* | 40% | | 33% | 46% | 34% | | | | |
| | 2 (reduced protection) | *AZ* | 19% | | 9% | 22% | 16% | 24% | 14% | | |
| | | *PF/Mod* | 19% | | 9% | 22% | 16% | 24% | 14% | | |

### 6 Software and Implementation

Implementation of the model described above is fully described in FitzJohn et al. [55]. The primary interface to the model is coded in R [56] with functions written in packages `sircovid` and `spimalot`. The model is written in `odin` and run with `dust`, the pMCMC functions are written in `mcstate`. For this paper we used `sircovid` v0.13.14, `spimalot` v0.7.11 on branch `sarscov2-vaccine-delay`, `dust` v0.11.26, and `mcstate` v0.9.1. The above packages are publicly available in the *mrc-ide* GitHub organisation (https://github.com/mrc-ide/). The code and scripts used to create the results in this paper are available in https://github.com/mrc-ide/sarscov2-roadmap-england.

### 7 Supplementary Results

#### 7.1 Model fit to data

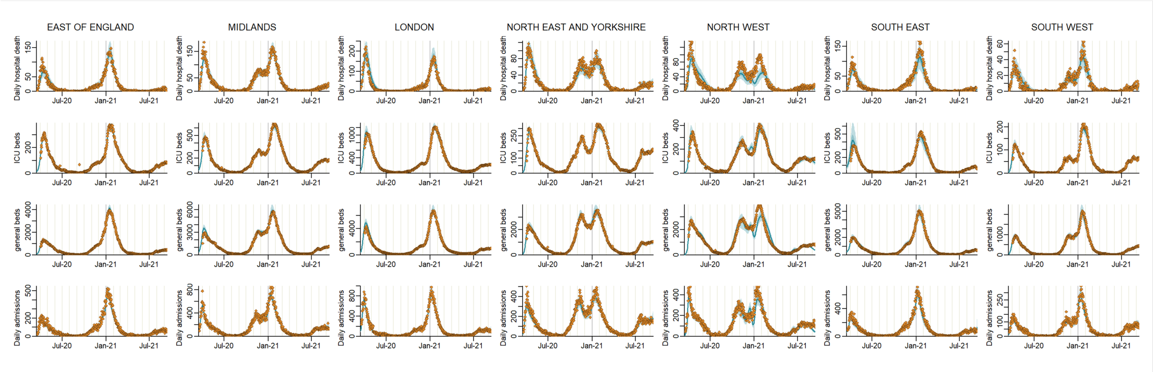

**Figure S11:** Model fits to NHS England Regions (columns): daily hospital deaths (top row), ICU beds occupancy (second row), general beds occupancy (third row) and all daily admissions (bottom row). The points show the data, the solid line the median model fit and the shaded area the 95% CrI.

The estimated beta values capture seasonal effects on transmissibility, the impact of school closures, and changing social mixing patterns. See Figure S12 and Figure S13. We use these fitted values for our counterfactual scenario.

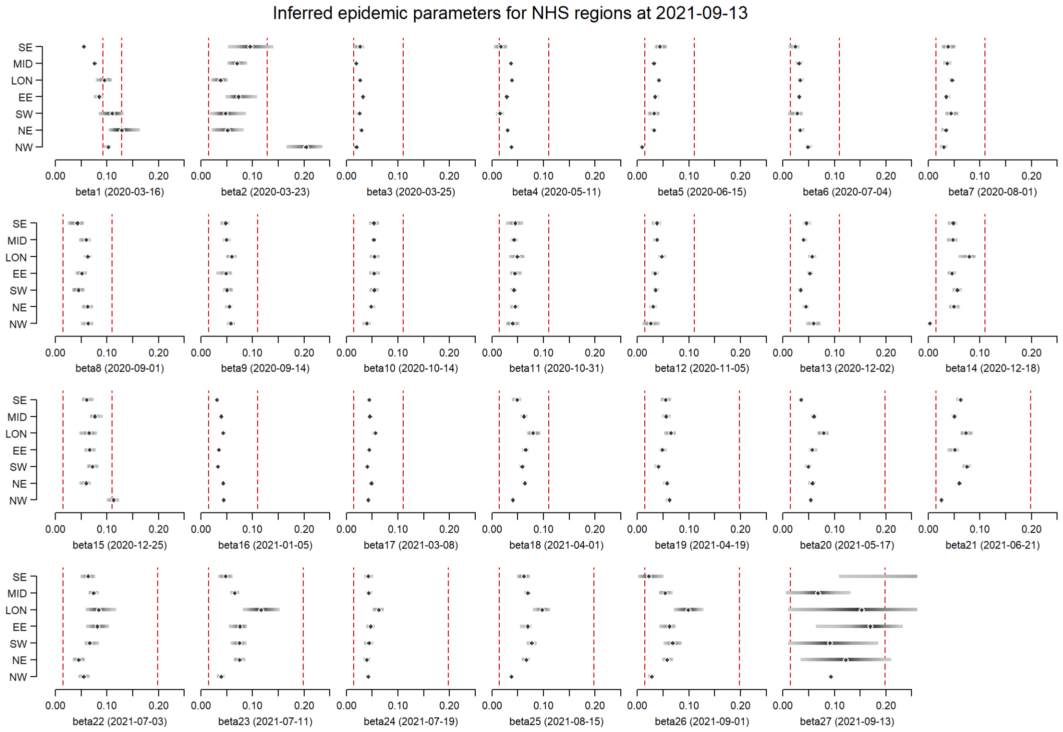

**Figure S12:** Estimated value of beta by region. The points show the median model fit and the shaded areas the 95% CrI. The vertical dashed red lines show the bounds of the prior distribution.

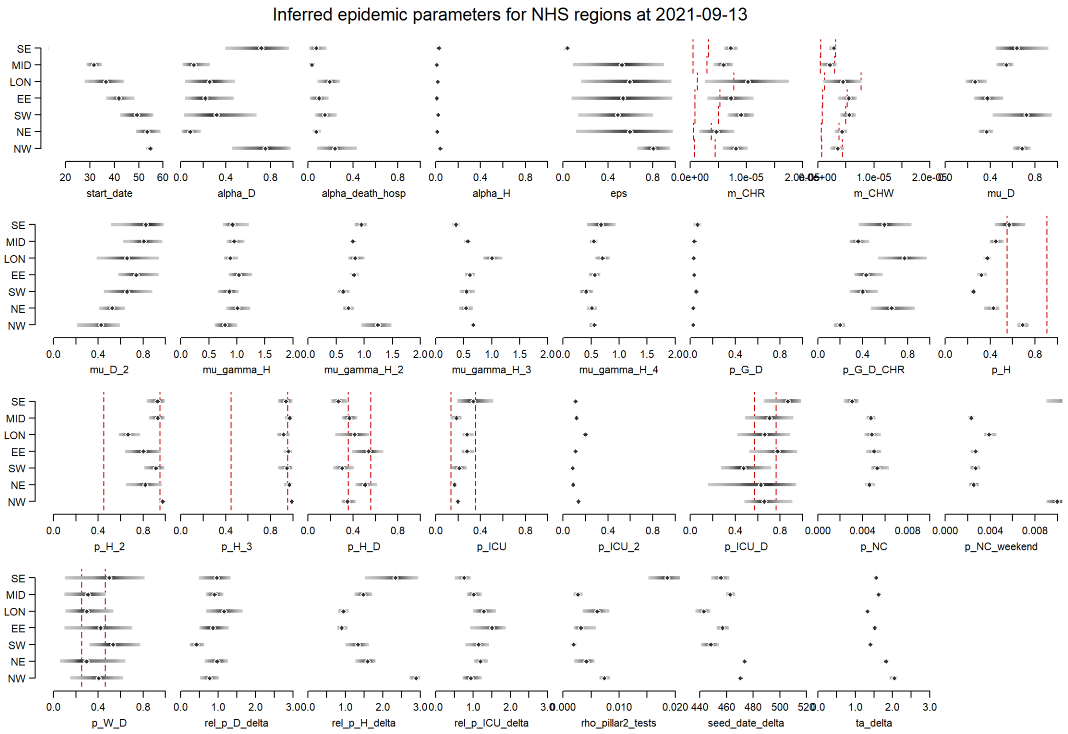

**Figure S13:** Model parameter posterior distributions. The points show the median model fit and the shaded areas the 95% CrI. The vertical dashed red lines show the bounds of the prior distribution.

#### 7.2 Hospital admissions by age

Although we do not fit explicitly to age-stratified data, our model is able to capture trends in hospitalisation by age well. Our model is fit to aggregated hospitalisations as reported on the Government COVID-19 dashboard [9]. However, due to missing information for some hospitalised cases, there are small discrepancies compared to the associated dataset with age-specific information. Our exploration of the secondary-uses services (SUS) linelist similarly has discrepancies.

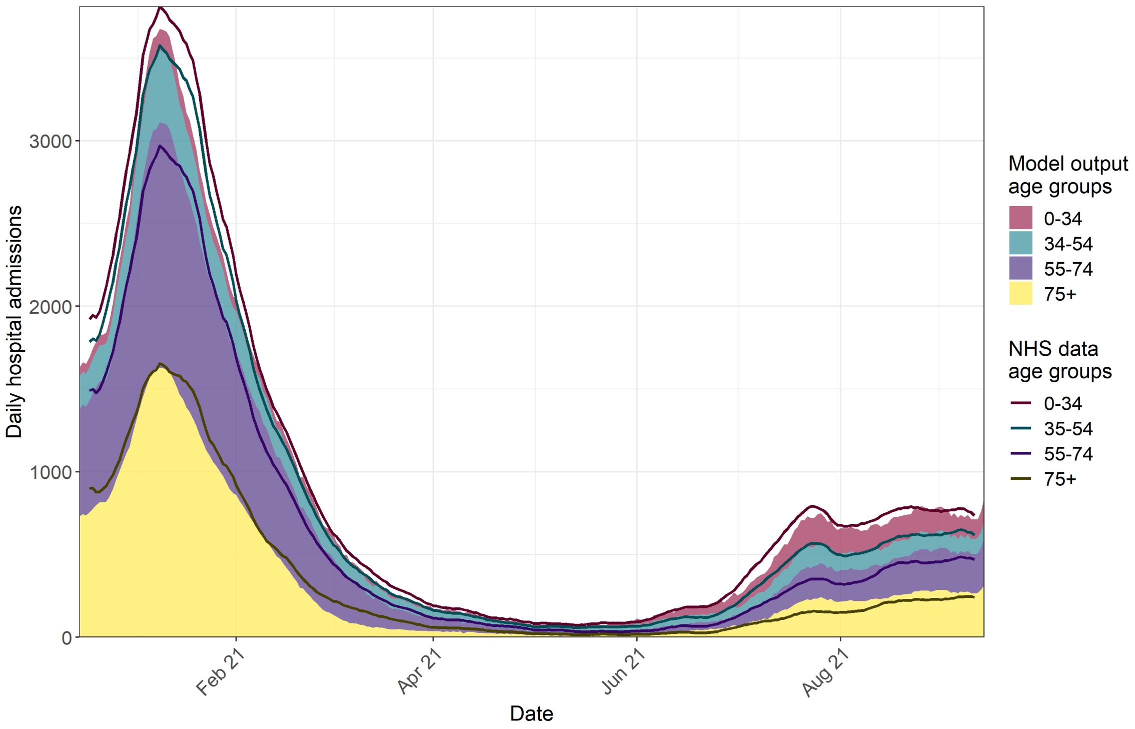

**Figure S14:** NHS reported daily COVID-19 hospital admissions by age group. Block colours show the modelled outputs by age group and the solid line shows the reported number of hospitalisations in the same age groups in the NHS data.

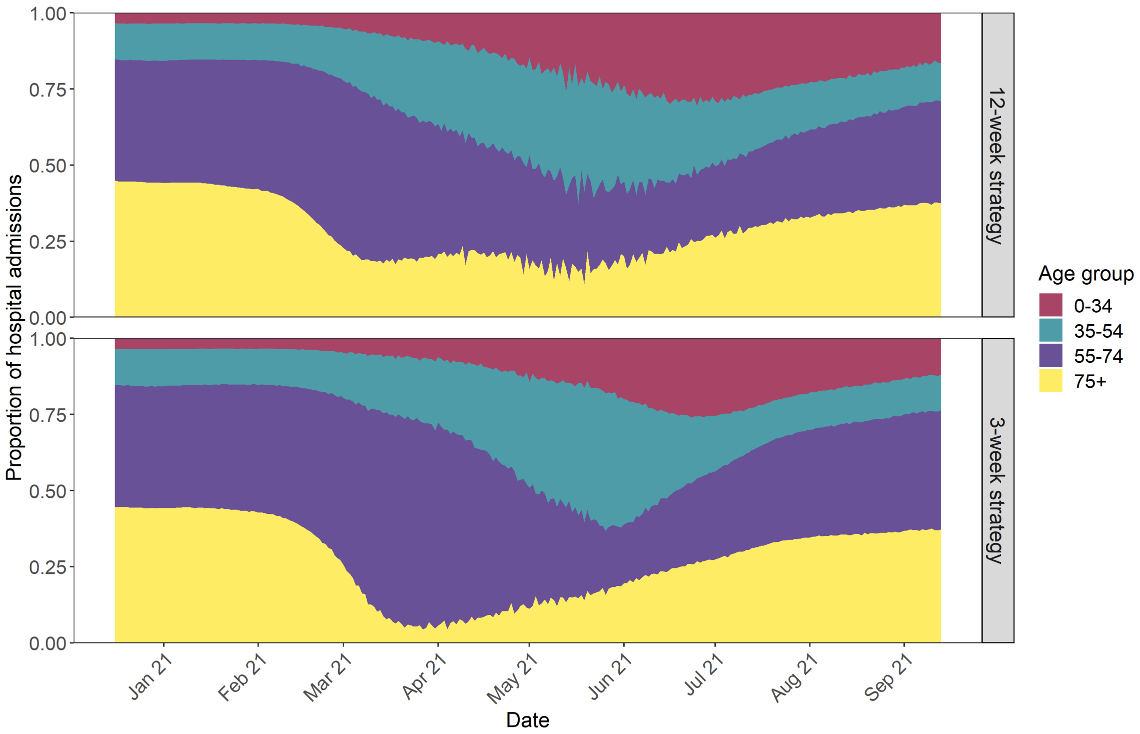

**Figure S15:** Proportion of COVID-19 hospital admissions over time by age group under the 12-week (top row) and 3-week (bottom row) strategies

#### 7.3 Results by vaccination status

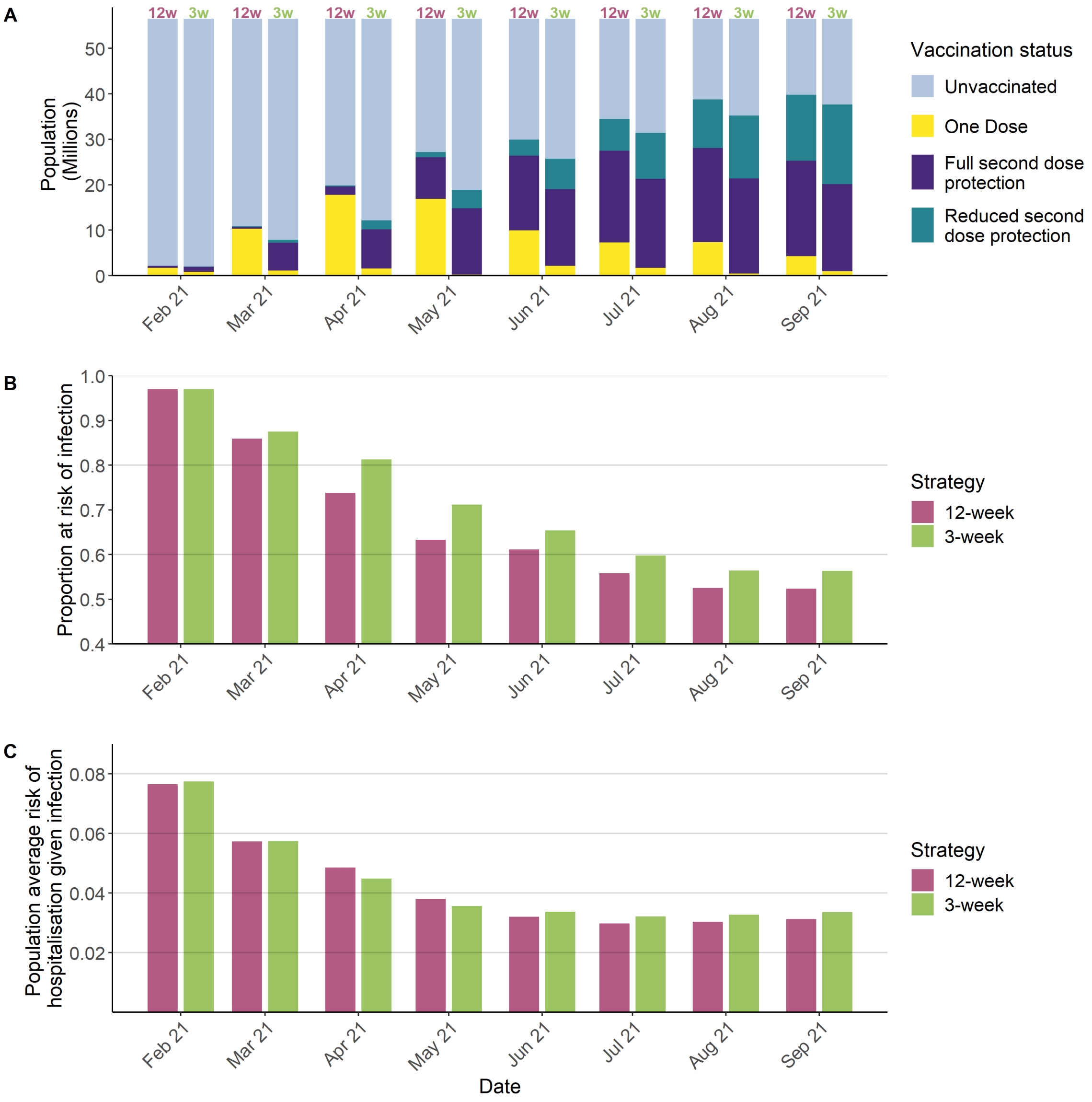

**Figure S16:** Vaccine protection against COVID-19. A) Vaccination status within the population (millions of people) for each month for the 12-week strategy (model fit, "12w") and 3-week strategy (counterfactual, "3w"). Colours show how the distribution of one-dose and two-dose vaccine-induced protection, and the unvaccinated population changes over time. B) Average vaccine-induced protection against infection with Alpha or Delta. C) Average probability of hospitalisation if infected, over time for the 12-week strategy (model fit, purple) and 3-week strategy (counterfactual, green).

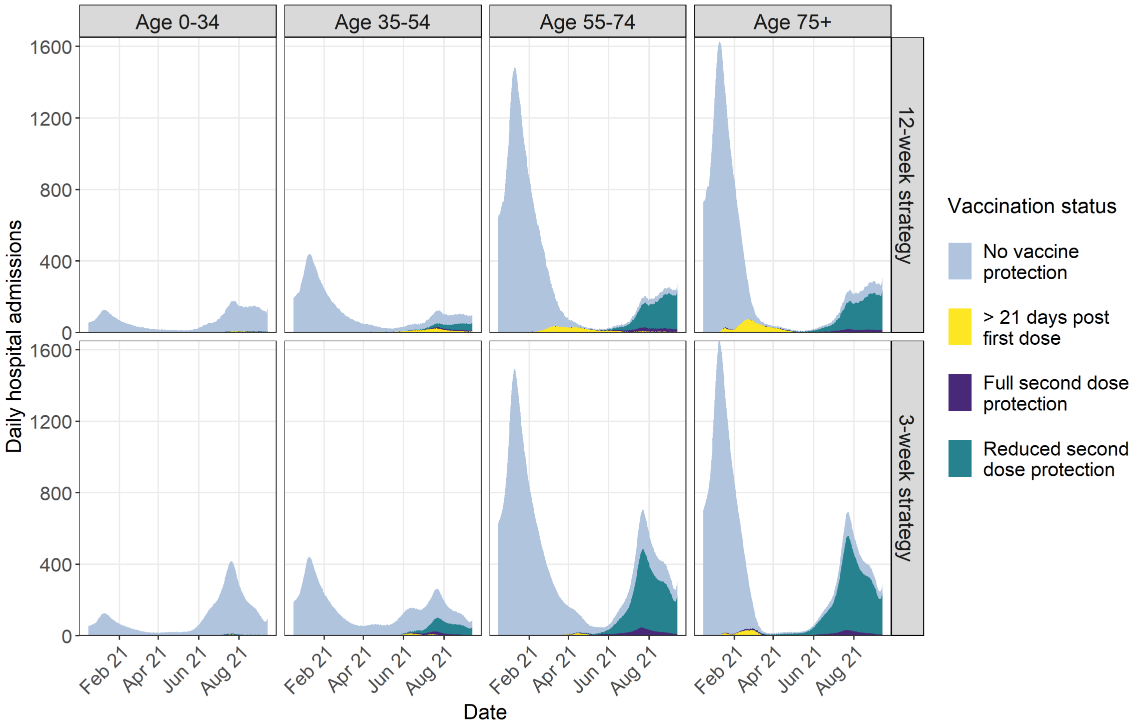

**Figure S17:** Modelled daily COVID-19 hospital admissions over time by age group (columns) for the 12-week (top row) and 3-week (bottom row) strategy. Colours show the vaccination status of hospitalised individuals.

### 7.4 Sensitivity Analyses Results

Figure S18 to Figure S25 show results of all sensitivity analyses which are described in Table S9 to Table S11.

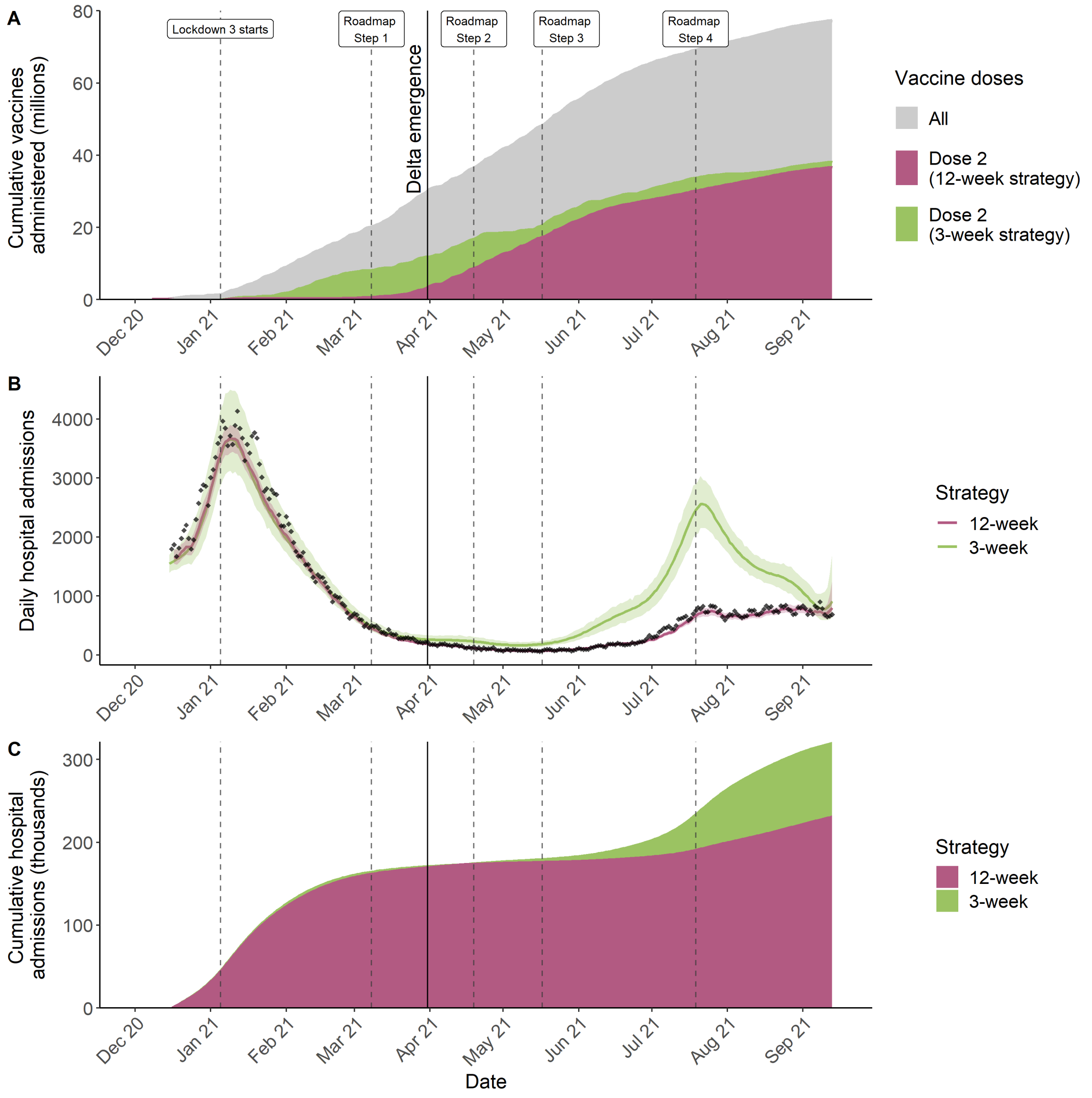

**Figure S18:** COVID-19 vaccines administered and trajectory of the COVID-19 epidemic in England. Figure is equivalent to main text Figure 2, but results are shown here for a 10% absolute reduction in VE against mild disease or infection against Alpha and Delta under the 3-week strategy (see Table S9 and Table S11). A) Cumulative vaccine doses administered between 8th December 2020 and 13th September 2021. Reported (purple) and counterfactual (green) second doses administered over time and all (first and second doses) are shown in grey. B) Observed (black points) daily hospital admissions. Purple line shows the median model fit and the shaded area the 95% CrI. The purple line and shaded area show the median/mean simulated daily hospital admission and the 95% CrI respectively under the counterfactual assuming a 3-week delay between vaccine doses was adhered to. C) Reported (purple) and counterfactual (green) cumulative hospital admissions over time. The vertical dashed lines show the roadmap out of lockdown steps and the vertical black line the time when the Delta variant emerged.

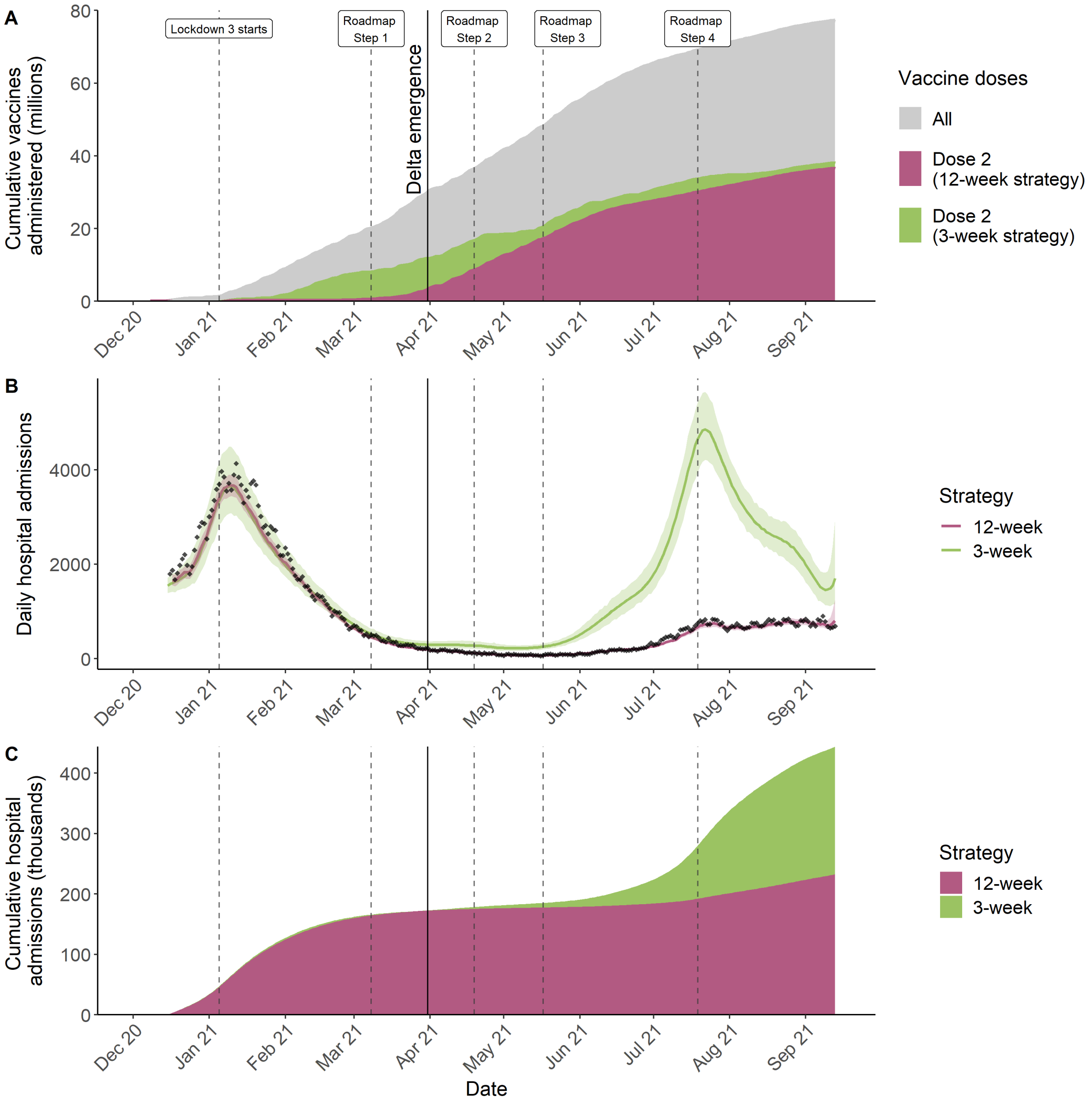

**Figure S19:** As Figure S18 but results are shown for a 10% absolute reduction in VE against all end points against Alpha and Delta under the 3-week strategy (see Table S9 and Table S11)

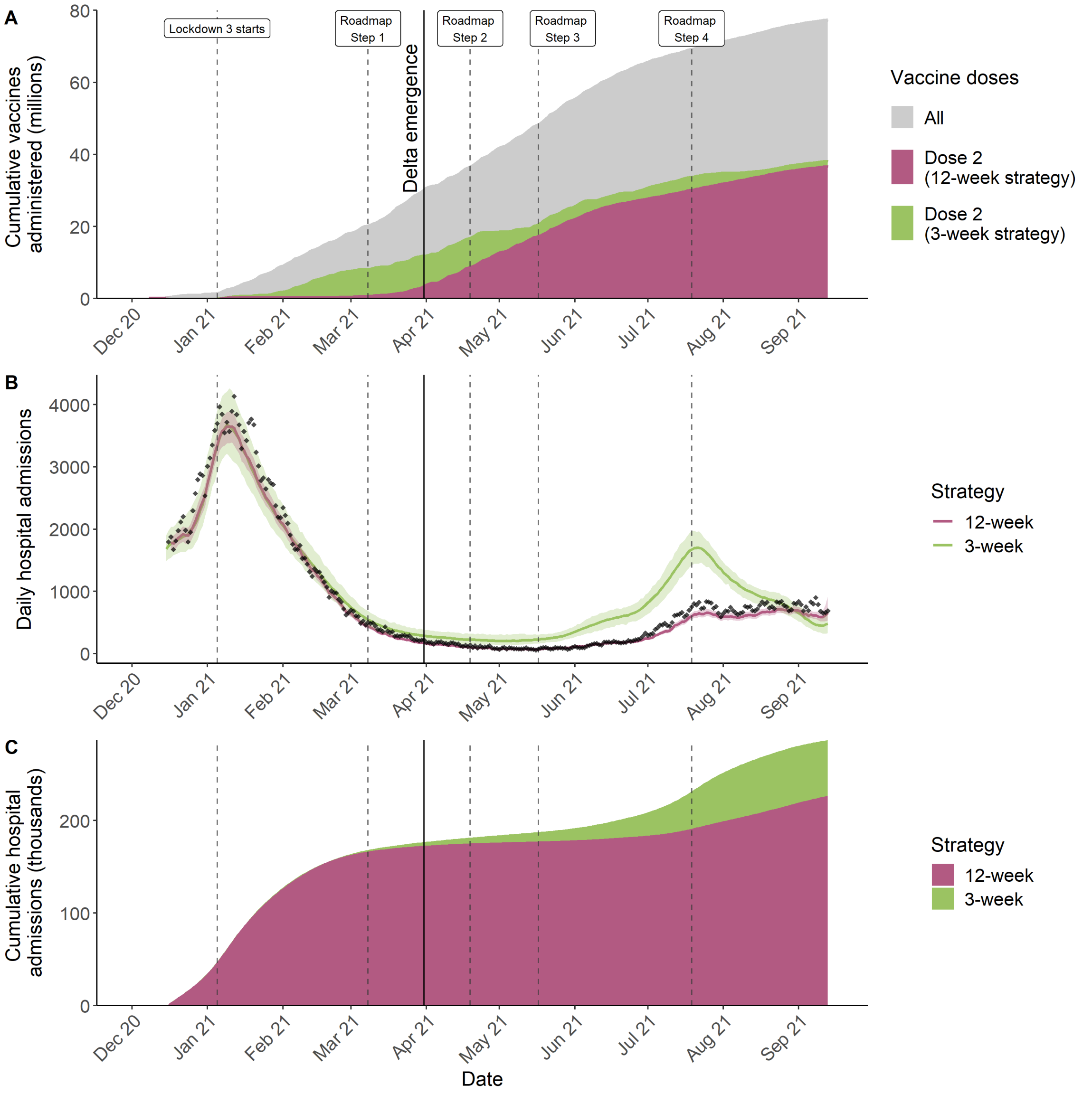

**Figure S20:** As Figure S18 but results are shown for a 15% relative increase in VE against all end points against Delta under both 12- and 3-week strategies (see Table S9 and Table S11).

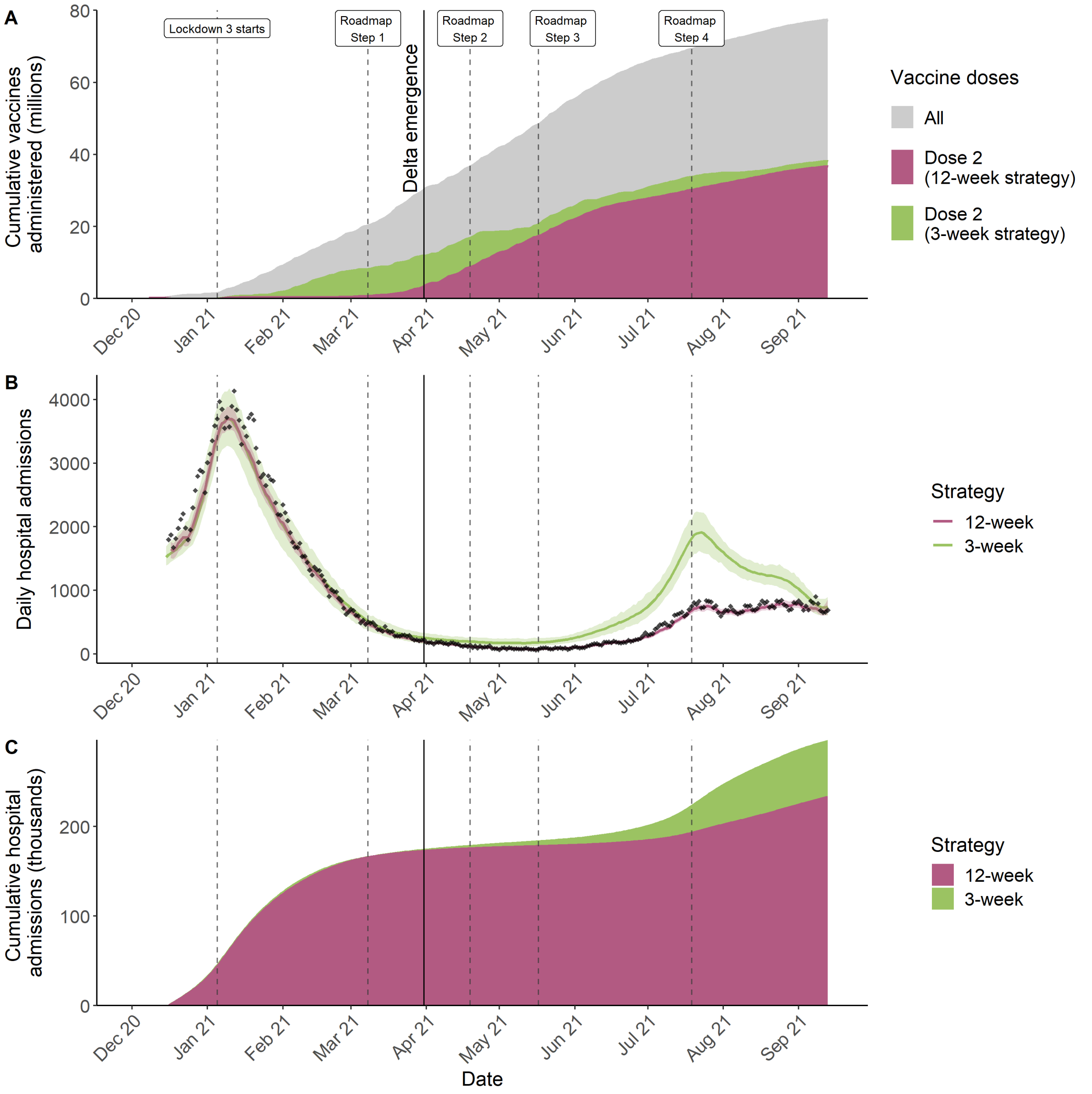

**Figure S21:** As Figure S18 but results are shown for a 15% relative decrease in VE against all end points against Delta under both 12- and 3-week strategies (see Table S9 and Table S11).

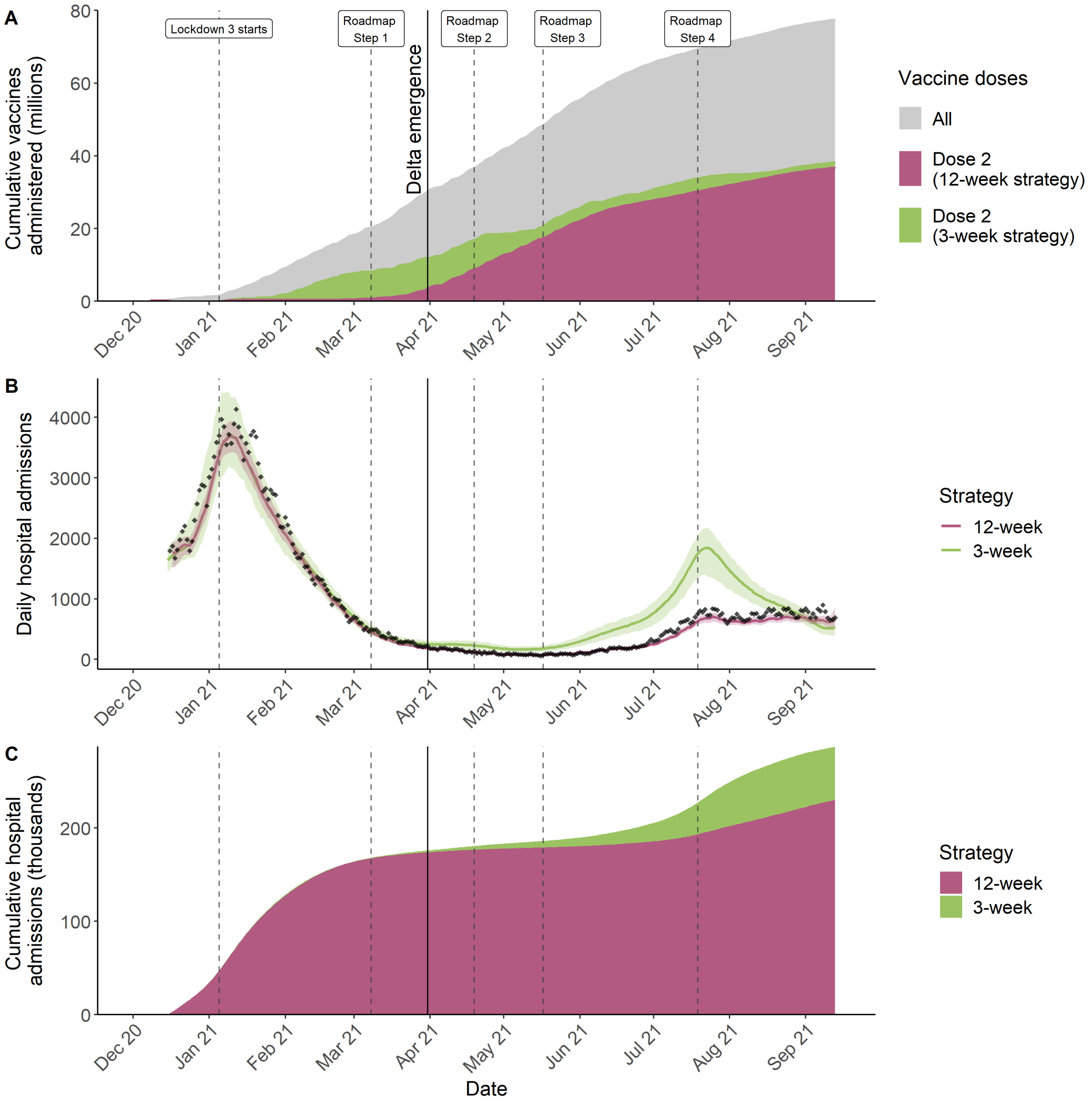

**Figure S22:** As Figure S18 but results are shown for a higher VE after 24-weeks after second dose against Delta under both 12- and 3-week strategies (see Table S9 and Table S11).

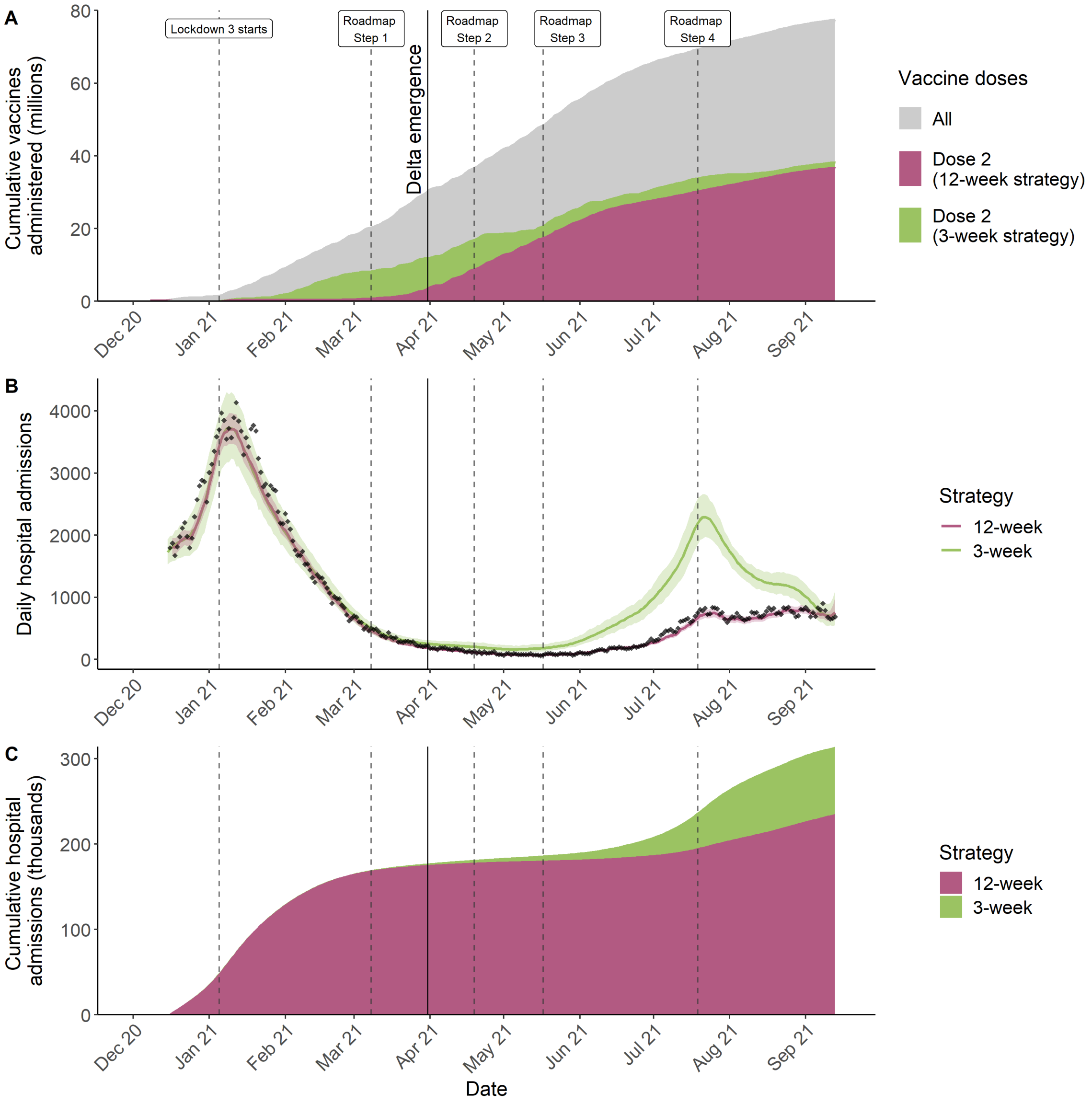

**Figure S23:** As Figure S18 but results are shown for a lower VE after 24-weeks after second dose Delta under both 12- and 3-week strategies (see Table S9 and Table S11).

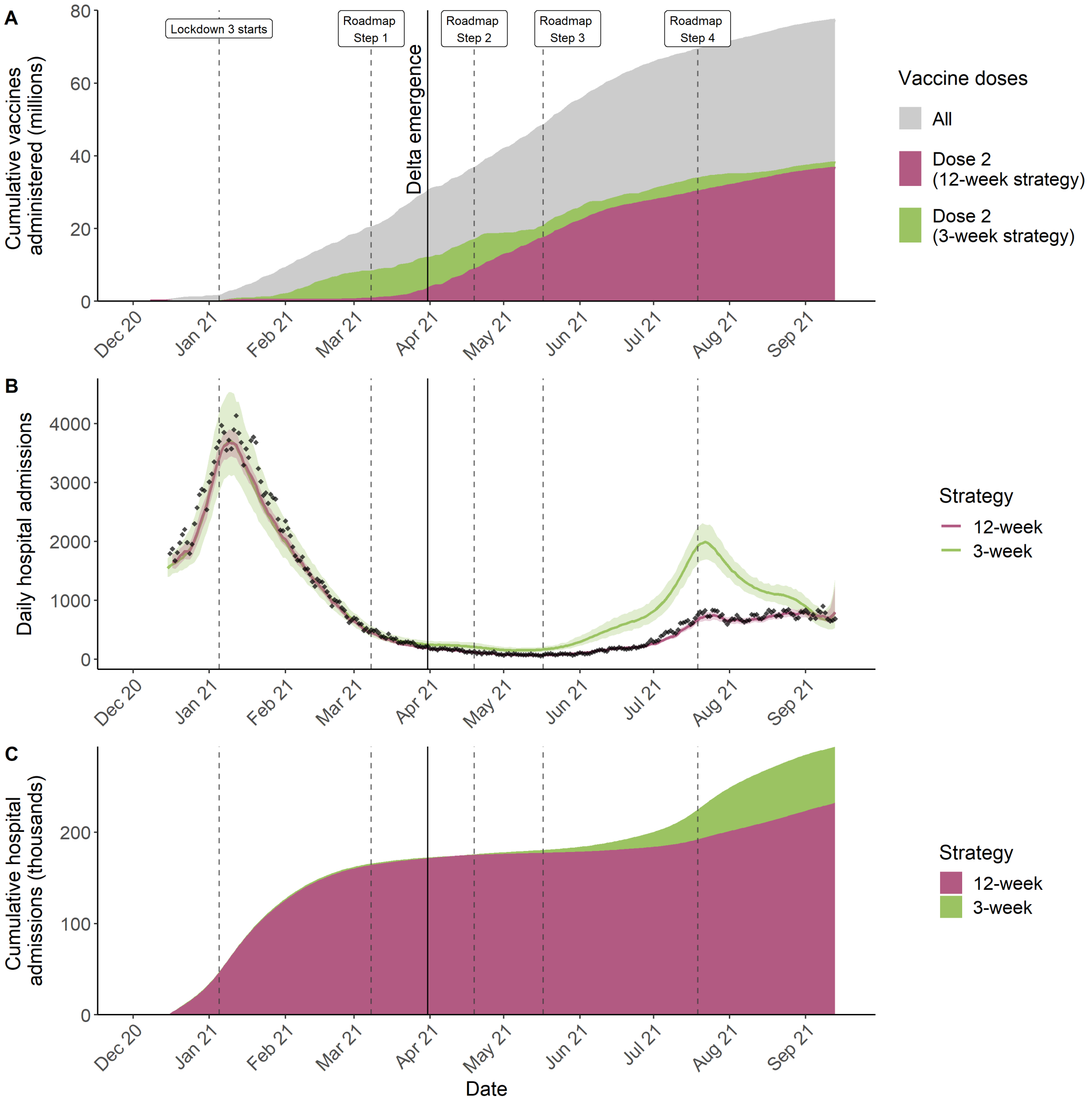

**Figure S24:** As Figure S18 but results are shown for a higher VE for the 3-week strategy to account for first dose waning under the 12-week strategy (see Table S9 and Table S11).

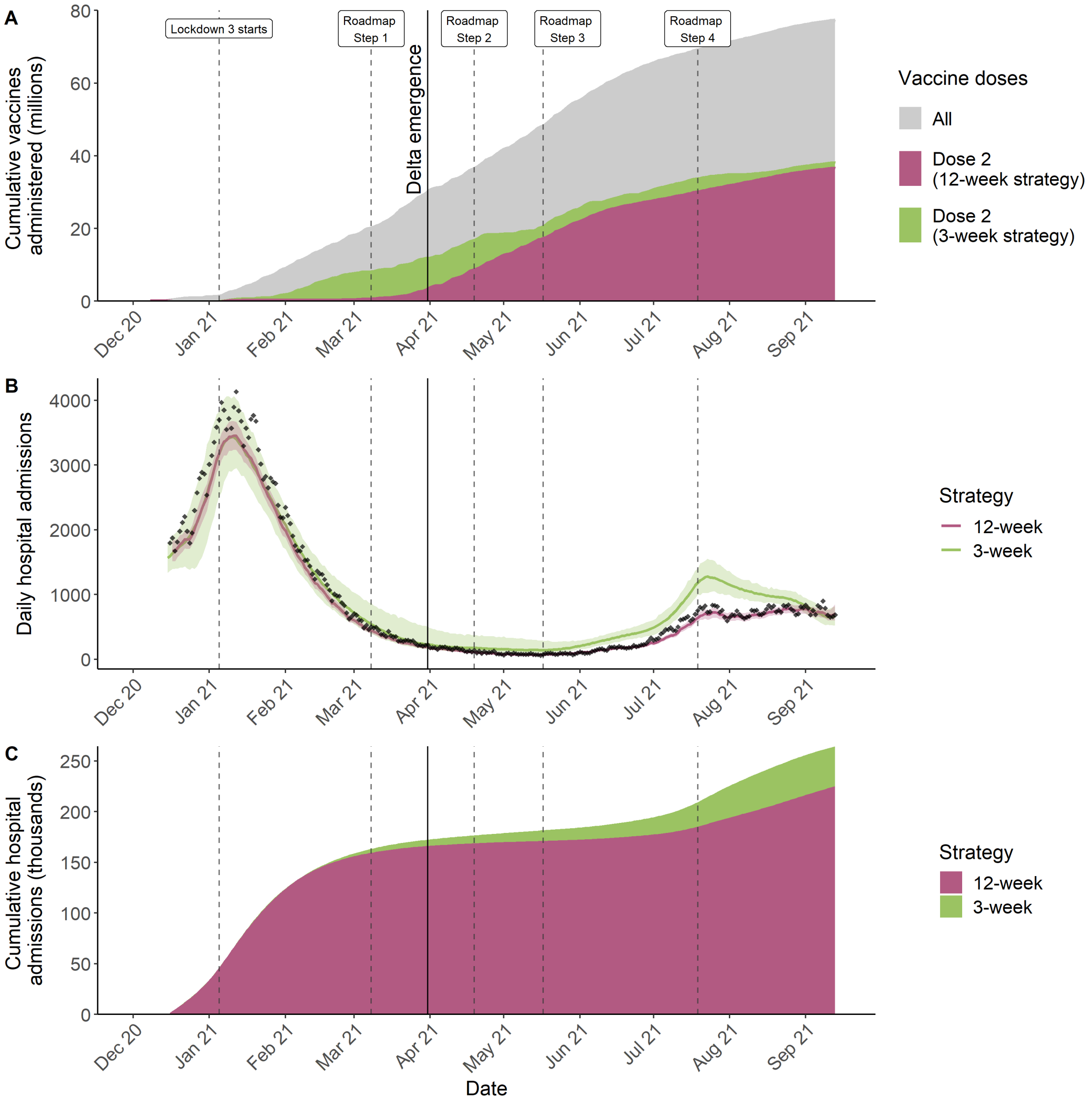

**Figure S25:** As Figure S18 but results are shown for immediate 1st dose vaccine-induced protection upon vaccination with a corresponding absolute 10% decrease in VE for both Alpha and Delta VE (see Table S9 and Table S11).

**Symbol Glossary**

| Symbol | Definition |
|---|---|
| | **Abbreviations** |
| CHW | Carehome workers |
| CHR | Carehome residents |
| ICU | Intensive care unit |
| VE | Vaccine effectiveness |
| AZ | AstraZeneca ChAdOx1 nCoV-19 (AZD1222) vaccine |
| PF | Pfizer-BioNTech COVID-19 Vaccine BNT162b2 |
| Mod | Moderna mRNA-1273 vaccine |
| | **Model Compartments** |
| $S^{ik}$ | Susceptible |
| $E^{i,j,k}$ | Exposed |
| $I_P^{i,j,k}$ | Infected pre-symptomatic |
| $I_A^{i,j,k}$ | Infected asymptomatic |
| $I_{C_1}^{i,j,k}$ | Symptomatic infected (infectious) |
| $I_{C_2}^{i,j,k}$ | Symptomatic infected (not infectious) |
| $G_D^{i,j,k}$ | Severe disease, not hospitalised |
| $D^{i,j,k}$ | Deceased (as a result of COVID-19) |
| $R^{i,j,k}$ | Recovered |
| $V_k$ | Vaccination strata |
| $ICU_{pre}^{i,j,k}$ | Awaiting admission to ICU |
| $ICU_{W_R}^{i,j,k}$ | Hospitalised in ICU, leading to recovery |
| $ICU_{W_D}^{i,j,k}$ | Hospitalised in ICU, leading to death following step-down from ICU |
| $ICU_D^{i,j,k}$ | Hospitalised in ICU, leading to death |
| $W_D^{i,j,k}$ | Step-down post-ICU period, leading to death |
| $W_R^{i,j,k}$ | Step-down post-ICU recovery period |
| $H_D^{i,j,k}$ | Hospitalised on general ward leading to death |
| $H_R^{i,j,k}$ | Hospitalised on general ward leading to recovery |
| | **Model Parameters** |
| $p_H^{i,j,k}(t)$ | Probability of hospitalisation given symptomatic |
| $p_{G_D}^{i,j,k}$ | Probability of dying in the community/care home given severe disease requiring hospitalisation |
| $p_{ICU}^{i,j}(t)$ | Probability of ICU admission given hospitalised |
| $p_{H_D}^{i,j,k}(t)$ | Probability of death given hospitalised and not in ICU |
| $p_{ICU_D}^{i,j,k}(t)$ | Probability of death given ICU |
| $p_{W_D}^{i,j,k}(t)$ | Probability of death after discharge |
| $\chi^{i,j,k}(t)$ | Susceptibility of an individual to variant $j$ given vaccine stratum $k$ |
| $\xi^{i,j,k}(t)$ | Infectivity of an individual infected with variant $j$ given vaccine stratum $k$ |
| $\lambda^{i,j,k}(t)$ | Variant-specific force of infection |
| $\Lambda^{i,k}(t)$ | Combined force of infection (both variants) |

Continued on next page

| **Table S12 – continued from previous page** | |
|---|---|
| Symbol | Definition |
| $\zeta^{i,k}(t)$ | Rate of progression from vaccine strata $k$ to $k+1$ |
| $\gamma_x$ | Rate of progression from compartment $x$ |
| $R^j(t)$ | Reproduction number for variant $j$ at time $t$ |
| $R_e^j(t)$ | Effective reproduction number for variant $j$ at time $t$ |
| $t_{Alpha}$ | Region specific outbreak start time |
| $t_{Delta}$ | Region specific Delta seeding time |
| $\nu_j$ | Duration of seeding period for variant $j$ |
| $\phi_j$ | Daily seeding rate for variant $j$ |
| $\delta^{i,j,k}(t)$ | Daily seeding rate of variant $j$ (stratified by age and vaccination strata) |
| $\sigma$ | Delta transmission advantage |
| $m_{i,i'}(t)$ | Person-to-person transmission rate |
| $c_{i,i'}$ | Person-to-person contact rate |
| $\beta(t)$ | Transmission rate |
| $\beta_i$ | Transmission rate at change-point $t_i$ |
| $\Theta_{i,j,k}(t)$ | Weighted number of infectious individuals |
| $\varepsilon$ | Relative reduction in contacts between CHR and general population |
| $\Delta_I^{i,k}$ | Mean duration of infectiousness weighted by infectivity |
| $\hat{\sigma}(t)$ | Ratio of effective reproduction number for Alpha and Delta ($R^{Alpha}(t)$ and $R^{Delta}(t)$). |
| | **Vaccine Effectiveness vs.** |
| $e_{inf}$ | Infection |
| $e_{sympt}$ | Symptoms |
| $e_{SD}$ | Severe disease |
| $e_{death}$ | Death |
| $e_{sympt\|inf}$ | Symptoms given infection |
| $e_{SD\|sympt}$ | Severe disease given symptoms |
| $e_{death\|SD}$ | Death given severe disease |
| $e_{ins}$ | Infectiousness |
| | **Fixed Parameters** |
| $p_C^{i,j,k}$ | Probability of being symptomatic given infected |
| $p^*(t)$ | Probability of COVID-19 diagnosis confirmed prior to hospital admission |
| $\gamma_U$ | Rate at which unconfirmed hospital patients are confirmed as infected |
| $\gamma_x$ | Rate at which individuals move out of compartment x |
| $p_{sero_{pos}}$ | Probability of seroconversion following infection |
| $p_{sero_{spec}}$ | Specificity of serology test |
| $p_{sero_{sens}}$ | Sensitivity of serology test |
| $1/\gamma_{sero_{pre}}$ | Mean time to seroconversion from onset of infectiousness |
| $1/\gamma_{sero^1_{pos}}$ | Mean duration of seropositivity (Euroimmun assay) |
| $1/\gamma_{sero^2_{pos}}$ | Mean duration of seropositivity (Roche N) |
| $\eta$ | Probability of cross-immunity to Delta following infection from Alpha |
| $\theta_{I_A}$ | Infectivity of an asymptomatic individual, relative to a symptomatic one |
